# Additional heterologous versus homologous booster vaccination in immunosuppressed patients without SARS-CoV-2 antibody seroconversion after primary mRNA vaccination: a randomized controlled trial

**DOI:** 10.1101/2021.09.05.21263125

**Authors:** Michael Bonelli, Daniel Mrak, Selma Tobudic, Daniela Sieghart, Maximilian Koblischke, Peter Mandl, Barbara Kornek, Elisabeth Simader, Helga Radner, Thomas Perkmann, Helmuth Haslacher, Margareta Mayer, Philipp Hofer, Kurt Redlich, Emma Husar-Memmer, Ruth Fritsch-Stork, Renate Thalhammer, Karin Stiasny, Stefan Winkler, Josef S. Smolen, Judith H. Aberle, Markus Zeitlinger, Leonhard X. Heinz, Daniel Aletaha

**Affiliations:** Division of Rheumatology, Department of Internal Medicine III, Medical University of Vienna, Austria; Division of Infectious Diseases and Tropical Medicine, Department of Internal Medicine I, Medical University of Vienna, Austria; Center for Virology, Medical University of Vienna, Austria; Department of Neurology, Medical University of Vienna, Austria; Department of Laboratory Medicine, Medical University of Vienna, Austria; Department of Pathology, Medical University of Vienna, Vienna, Austria; 2^nd^ Department of Medicine, Hietzing Hospital, Vienna, Austria; School of Medicine, Sigmund Freud University Vienna, Vienna, Austria; 1^st^ Medical Department, Hanusch Hospital of the Austrian Health Insurance Fund, Vienna, Austria; Ludwig Boltzmann Institute of Osteology, Hanusch Hospital and AUVA Trauma Center Meidling, Vienna, Austria; Departement of Clinical Pharmacology, Medical University of Vienna, Austria

**Keywords:** SARS-CoV-2, Rituximab, vaccination, immune response

## Abstract

Severe acute respiratory syndrome coronavirus-2 (SARS-CoV-2)-induced coronavirus disease 2019 (COVID-19) has led to exponentially rising mortality, particularly in immunosuppressed patients, who inadequately respond to conventional COVID-19 vaccination. In this blinded randomized clinical trial (EudraCT 2021-002348-57) we compare the efficacy and safety of an additional booster vaccination with a vector versus mRNA vaccine in non-seroconverted patients. We assigned 60 patients under rituximab treatment, who did not seroconvert after their primary mRNA vaccination with either BNT162b2 (Pfizer–BioNTech) or mRNA-1273 (Moderna), to receive a third dose, either using the same mRNA or the vector vaccine ChAdOx1 nCoV-19 (Oxford-AstraZeneca). Patients were stratified according to the presence of peripheral B-cells. The primary efficacy endpoint was the difference in the SARS-CoV-2 antibody seroconversion rate between vector (heterologous) and mRNA (homologous) vaccinated patients by week four. Key secondary endpoints included the overall seroconversion and cellular immune response; safety was assessed at weeks one and four.

Seroconversion rates at week four were comparable between vector (6/27 patients, 22%) and mRNA (9/28, 32%) vaccine (p=0.6). Overall, 27% of patients seroconverted; specific T-cell responses were observed in 20/20 (100%) vector versus 13/16 (81%) mRNA vaccinated patients. Newly induced humoral and/or cellular responses occurred in 9/11 (82%) patients. No serious adverse events, related to immunization, were observed. This enhanced humoral and/or cellular immune response supports an additional booster vaccination in non-seroconverted patients irrespective of a heterologous or homologous vaccination regimen.

---

The current pandemic caused by severe acute respiratory syndrome coronavirus-2 (SARS-CoV-2) has led to exponentially rising morbidity and mortality worldwide. Apart from aggressive quarantine and infection control hygiene measures, the most effective way to combat SARS-CoV-2 spread is a population-wide vaccination strategy, foremost in those at high risk to develop severe COVID-19^1,2^. Two types of vaccines have been currently approved by the European Medicines Agency: vector vaccines, such as ChAdOx1 nCoV-19 (Oxford-AstraZeneca) and Ad26.COV2-S (Johnson&Johnson), and mRNA vaccines, such as BNT162b2 (Pfizer–BioNTech) and mRNA-1273 (Moderna). However, immune responses to these vaccines vary between individuals and antibody levels wane over time^3,4^. Application of an additional booster dose is heavily investigated by ongoing clinical trials and first reports have been published^5–8^. Several countries already started a third vaccination, especially in patients at high risk. Most recently, the United States Food and Drug Administration authorized an additional vaccine dose for certain immunocompromised patients^9^.

Patients under immunosuppressive therapy with rituximab, a B-cell-depleting monoclonal antibody against the CD20 surface antigen, are at a high risk for severe COVID-19 requiring hospitalization and ICU admission^10,11^. At the same time, B-cell depletion reduces immune responses to vaccination^12,13^. This combination poses a dilemma and therefore a highly unmet clinical need for this group of patients. Those lacking B lymphocytes in the periphery at the time of vaccination and thus did not yet start reconstituting their B-cell pool often fail to seroconvert^14,15^. Although B-cell depleted patients can develop a T-cell response, to date, it is unclear to what extent cellular and humoral responses contribute to protection against SARS-CoV-2 infection.

The development of a humoral immune response currently constitutes a good surrogate of protection, and its absence is therefore often considered an alarm signal for an insufficient vaccination response. In order to stimulate the humoral immune response of rituximab-treated patients who do not respond to the conventional scheme of COVID-19 vaccination, an additional booster vaccination may be an obvious clinical strategy. Recent studies also evaluated the safety and efficacy of homologous versus heterologous schemes for primary and secondary vaccination in healthy individuals^16–18^. However, it is unknown whether a heterologous approach could benefit those who completely lack a humoral immune response after basic immunization. Furthermore, no data exist on responses to an additional booster vaccination in patients who had completely failed to mount a specific antibody response after the primary two-vaccination schedule.

In this blinded, randomized, controlled trial, we addressed this question and the general inducibility of a humoral or T-cell response in rituximab-treated autoimmune disease patients without anti-SARS-CoV-2 antibodies after their basic mRNA vaccination.

## RESULTS

Sixty-eight patients under rituximab treatment who had been immunized with two doses of mRNA vaccine were screened for eligibility. Eight patients were excluded due to presence of detectable SARS-CoV-2-specific antibodies. 60 non-seroconverted patients were randomized, of whom 30 were assigned to receive vector and 30 to receive mRNA vaccine as the third dose; Five patients withdrew consent between screening and baseline visit **(Figure 1)**. A total of 27/30 patients were vaccinated with a vector vaccine and 28/30 received an mRNA vaccine. All patients subsequently presented at follow-up visits and completed the trial at week four after vaccination. Patient characteristics were similar between the two randomized groups **(Table 1)**.

**Figure 1:**
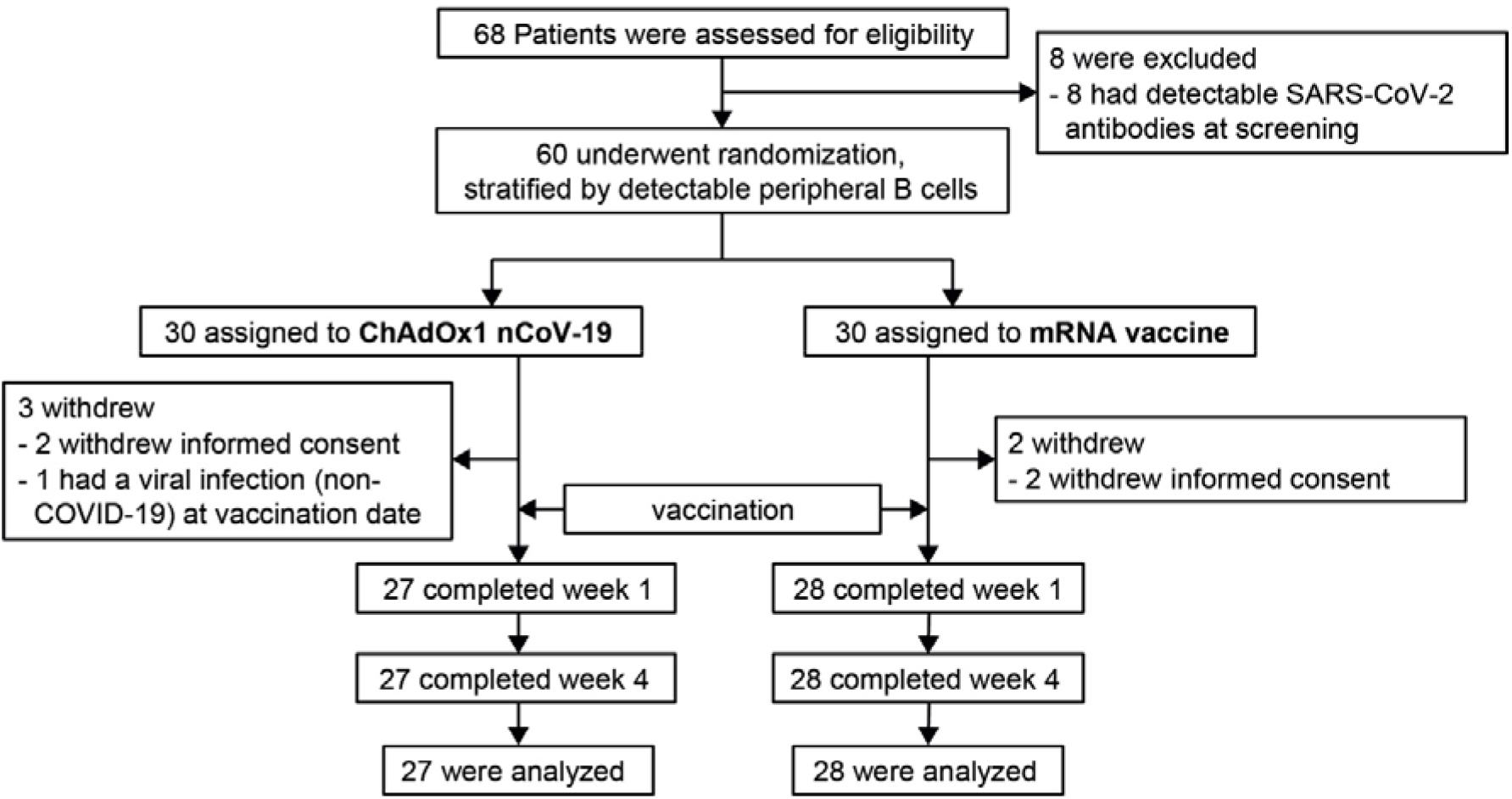
Screening, randomization and follow up of patients.

**Table 1.** Baseline characteristics of patients vaccinated with a third dose. Data are n (%) or mean (SD), csDMARD, conventional synthetic disease modifying antirheumatic drug, defined here as concomitant treatment with at least one of the following: methotrexate, mycophenolate mofetil, azathioprine, leflunomide, hydroxychloroquine.

|  | Vector | mRNA |
| --- | --- | --- |
| n | 27 | 28 |
| Age | 60.9 (15.0) | 58.9 (18.4) |
| Gender: female | 18 (66.7%) | 23 (82.1%) |
| **Diagnosis** |  |  |
| Arthritis | 11 (40.7%) | 10 (35.7%) |
| Connective tissue diseases | 7 (25.9%) | 9 (32.1%) |
| Vasculitis | 4 (14.8%) | 4 (14.3%) |
| Multiple sclerosis | 3 (11.1%) | 3 (10.7%) |
| IgG4-related disease | 2 (7.4%) | 2 (7.1%) |
| Months between RTX and screening | 7.0 (6.2) | 6.0 (3.6) |
| Weeks between 2<sup>nd</sup> vaccination and screening | 8.2 (3.7) | 6.6 (2.3) |
| Patients with detectable B-cells | 8 (29.6%) | 10 (35.7%) |
| **Concomitant medication** |  |  |
| Any csDMARD | 10 (37.0%) | 16 (57.1%) |
| Methotrexate | 3 (11.1%) | 7 (25.0%) |
| Mycophenolate mofetil | 2 (7.4%) | 4 (14.3%) |
| Azathioprine | 2 (7.4%) | 3 (10.7%) |
| Leflunomide | 3 (11.1%) | 1 (3.6%) |
| Hydroxychloroquine | 0 (0.0%) | 4 (14.3%) |
| Immunoglobulin therapy | 1 (3.7%) | 1 (3.6%) |
| Prednisone | 7 (25.9%) | 8 (28.6%) |
| Primary vaccination with BNT162b2 | 21 (78%) | 21 (75%) |
| Primary vaccination with mRNA-1273 | 6 (22%) | 7 (25%) |

Seroconversion rates at week four were numerically lower in the vector than in the mRNA group (6/27, 22% compared to 9/28, 32% of patients) **(Figure 2A)**. Despite the numerical difference in favor of the homologous vaccination group, disadvantage of the heterologous group cannot be supported statistically (p=0.6).

**Figure 2.**
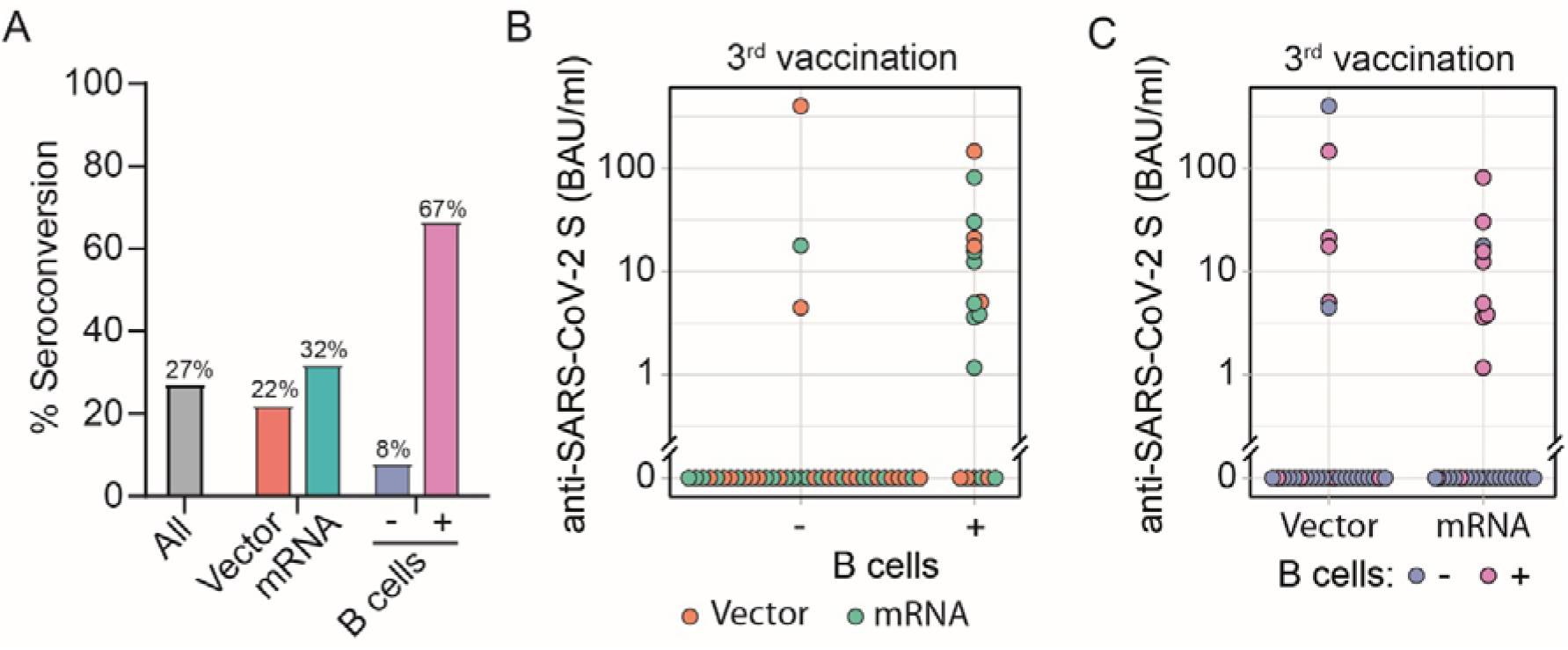
Antibody seroconversion rate four weeks after vector vs. mRNA booster vaccination. Antibodies to the receptor-binding domain (RBD) of the viral spike (S) protein were determined using an anti-SARS-CoV-2 immunoassay. **A**. Seroconversion rate was calculated based on the presence of anti-RBD antibodies in patients stratified by booster vaccination with vector or mRNA vaccine, in all patients and in patients with and without detectable peripheral B-cells **B**. Anti-RBD antibody levels in patients with (n=18) and without (n=37) peripheral B-cells, with color of the circles indicating the type of vaccine. **C**. Anti-RBD antibody levels in patients four weeks after booster vaccination with vector (n=27) or mRNA vaccine (n=28), with color of the circles indicating the presence or absence of detectable peripheral CD19^+^ B-cells.

Even though the primary endpoint was not met, 27% of all vaccinated patients seroconverted independent of the vaccine used with a median SARS-CoV-2 S antibody level of 15.7 [IQR: 4.7, 25.8] BAU/mL. Seroconversion rate was higher in patients with detectable peripheral CD19^+^ B-cells versus those without **(Figure 2A**). Among patients with no detectable peripheral B-cells (37/55, 67%) antibodies to the receptor-binding domain of the viral spike (S) protein (anti-RBD antibodies) were detectable in 3/37 (8%) patients; in patients with detectable peripheral B-cells, seroconversion rate was 67% (12/18) at week four **(Figure 2A to C)**. Median levels of anti-RBD antibodies were 19.4 [IQR: 8.2, 114.8] and 12.4 [IQR: 3.8, 17.8], respectively, in seroconverted vector and mRNA vaccinated patients **(Figure 2B; Supplementary Table 1)**.

SARS-CoV-2-specific T-cell responses were determined by ELISpot assay in all patients before and after booster vaccination. Matched samples before and after the third vaccination were available from 36 patients. Patient characteristics for this group stratified by third vaccination are presented as **Supplementary Table 2**. At screening, 15/20 (75%) of patients assigned to the vector group and 10/16 (63%) assigned to the mRNA group had detectable spike-specific T-cells. Administration of a third vaccine dose led to an increase to 20/20 (100%) in the vector and 13/16 (81%) in the mRNA group **(Figure 3A-B)**. The number of spot forming cells (SFC) to the spike peptide pools (S1/S2) was slightly higher after boost with vector vaccine (median: 459, IQR [133, 722] as compared to mRNA vaccine (median: 305, IQR [171, 416]) **(Figure 3C)**.

**Figure 3.**
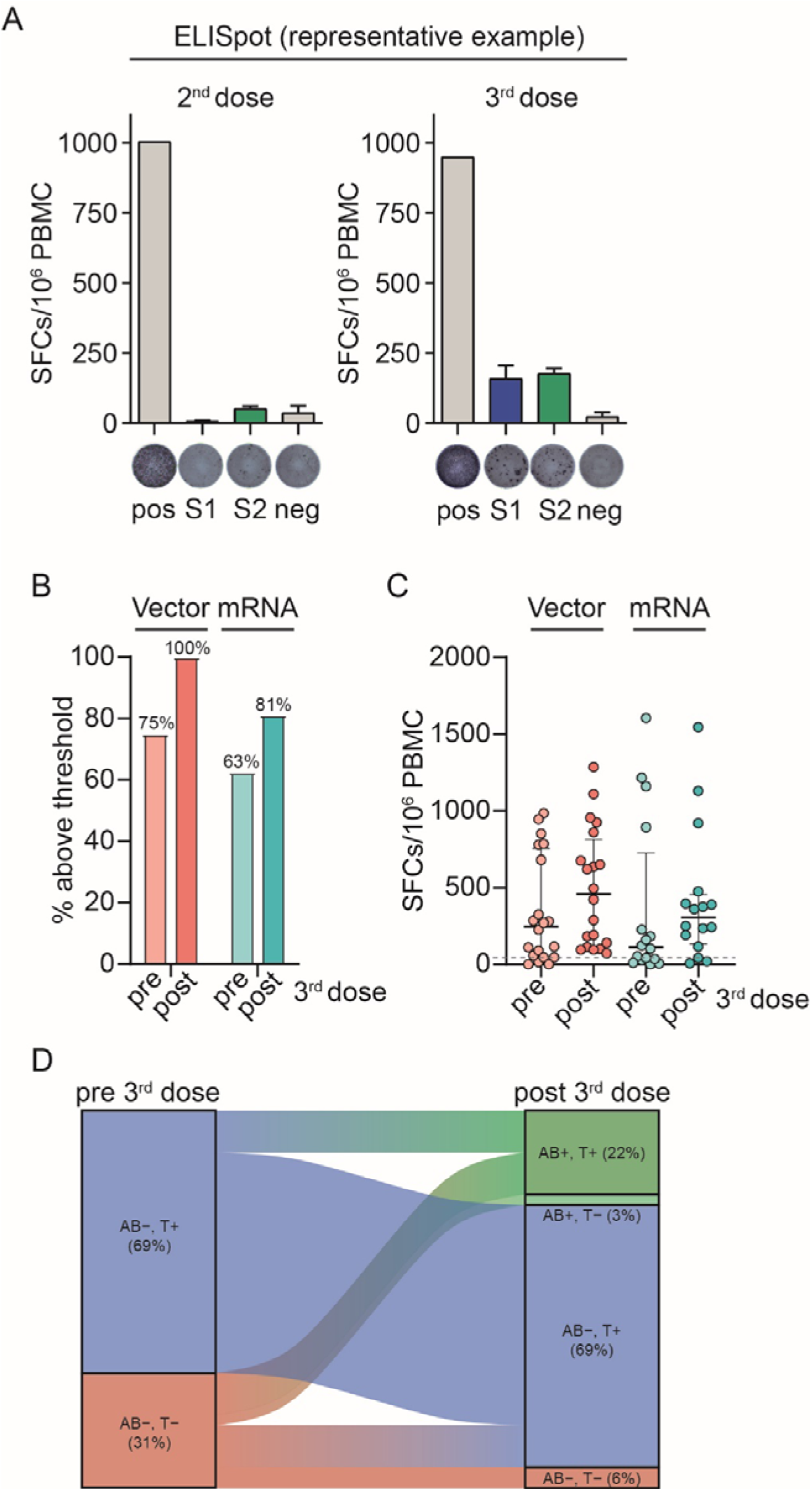
SARS-CoV-2-specific T-cell responses after additional vector or mRNA booster vaccination in Rituximab-treated patients. **A**. One representative *ex vivo* IFN-γ ELISpot result from peripheral blood mononuclear cells (PBMCs) stimulated with spike subunit S1 and S2 peptide pools shown for one patient before and after booster vaccination. Y-axis indicates the number of spot-forming cells (SFCs) per 10^6^ PBMCs. **B**. Percent of patients without T-cell response before and after third vaccination with vector and mRNA vaccine. **C**. Composite ELISpot results from 36 patients before and after the third vaccination with mRNA (n=16) and vector vaccine (n=20). Circles show sum of total response from S1 and S2 peptide pools. Vertical line shows median, whiskers interquartile range. Dotted lines represent the cut-off as defined by the mean SFC count plus three times the standard deviation from pre-pandemic controls. **D**. Humoral and cellular immune responses before and after the third vaccination. AB: Antibody, T: T-cell response, (-): negative, (+): positive

Integrative analysis of humoral and T-cell responses for 36 patients with matched samples before and after the third vaccination was performed: before third vaccination 11/36 (31%) had neither anti-RBD antibodies nor T-cell response (AB-,T-), and 25/36 patients (69%) did not have a humoral but exhibited a cellular immune response (AB-,T+). After the third vaccination 8/36 (22%) showed a humoral and T-cell response (AB+,T+), 1/36 (3%) had a humoral but no detectable cellular immune response (AB+,T-), in 25/36 (69%) a cellular but no humoral immune response (AB-,T+) was observed; 2/36 (6%) developed neither a humoral nor a cellular immune response. Overall a cellular and/or humoral immune response could be achieved through an additional booster vaccination in 9 out of 11 (82%) of those patients who did not respond to conventional vaccination strategy with two doses of mRNA vaccine **(Figure 3D, Supplementary Table 2 and 3)**.

Exploratory post-hoc univariate logistic regression models revealed that detectable peripheral B-cells strongly favored the likelihood of seroconversion (OR 22.67, 95%CI 5.46 to 125.10), while co-medication with any conventional synthetic disease modifying antirheumatic drug (csDMARD) favored non-seroconversion. Compared to mRNA booster vaccination, the vector vaccine showed a lower likelihood of inducing humoral response though not statistically significant. With respect to T-cell response, no association with age, use of prednisone or presence of peripheral B-cells could be observed **(Figure 4)**. All patients vaccinated with the vector regimen developed a T-cell response, while all patients without T-cell response were co-treated with a csDMARD, resulting in non-convergence of the respective regression models **(Supplementary Table 4)**.

**Figure 4:**
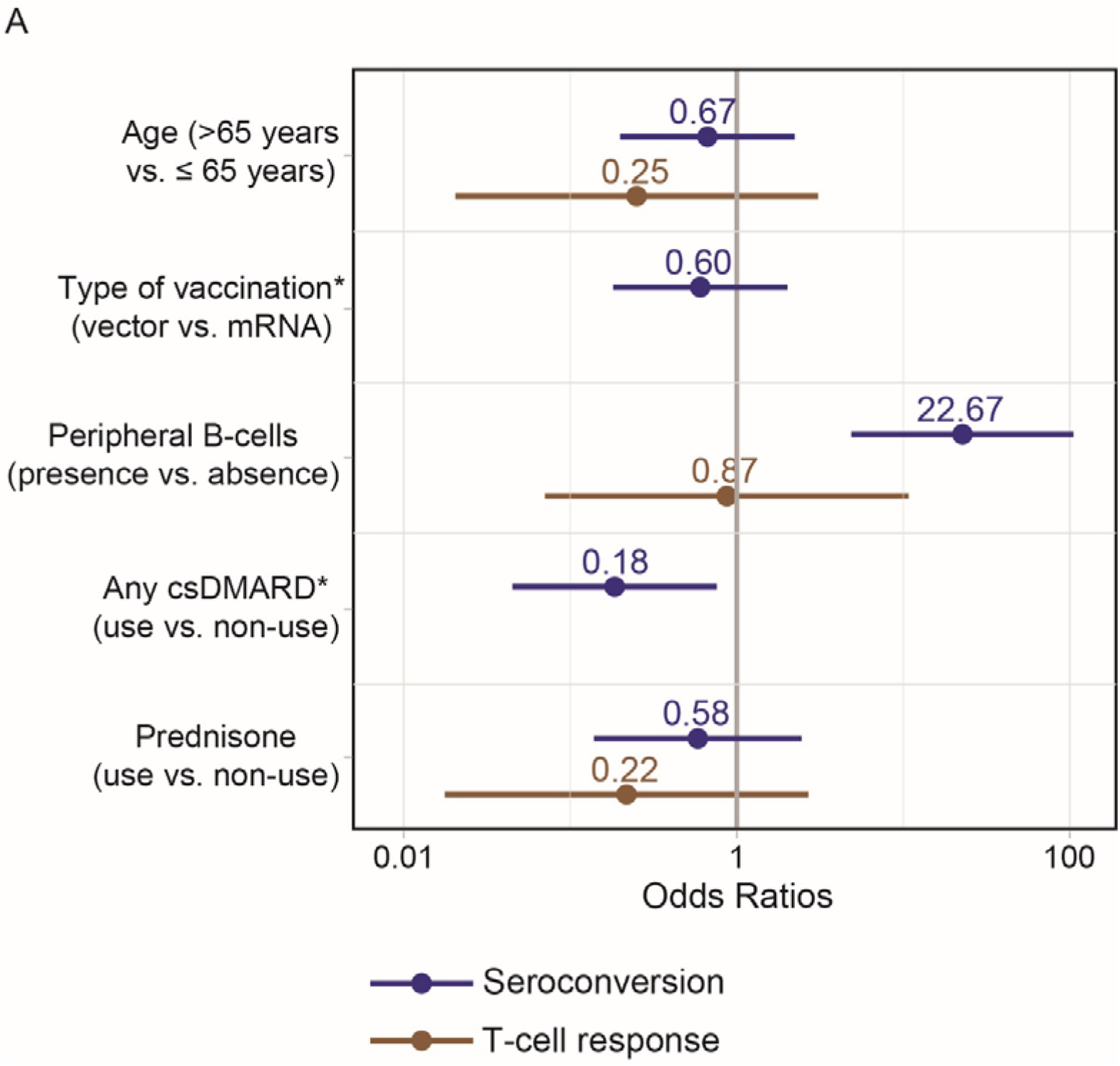
Odds ratios (OR) of logistic regression assessing humoral and cellular immune responses. ^*^All patients treated with vector vaccine developed a T-cell response and all patients without T-cell response were co-treated with csDMARDs, so consequently no OR could be calculated due to non-convergence of the respective models.

Systemic reactogenicity was evaluated by the patients using a paper-based diary daily during the first week after vaccination. Adverse events, in general, were monitored until 28 days after vaccination. One serious adverse event was reported after the screening visit prior to vaccination. Most side effects were similar between vector and mRNA booster vaccine groups. Numerically, a higher prevalence of systemic reactogenicity after the booster dose was reported by patients in the heterologous vaccine group compared to homologous vaccine schemes for fatigue, arthralgia and myalgias. 13/27 (48%) of vector-vaccinated patients developed arthralgias as compared to 8/28 (29%) patients with mRNA booster vaccination. Myalgia was reported in 15/27 (56%) vector-vaccinated patients compared to 9/28 (32%) mRNA-vaccinated patients. Fatigue was present in 21/27 (78%) vector-vaccinated patients, while only 13/28 (46%) mRNA-boosted patients experienced fatigue. Local pain at the injection site was more frequent during the first two days in mRNA-vaccinated (16/28, 57%) than vector-vaccinated patients (8/27, 30%). The local and systemic reactogenicity for the first week after vaccination is displayed in **Figure 5**.

**Figure 5:**
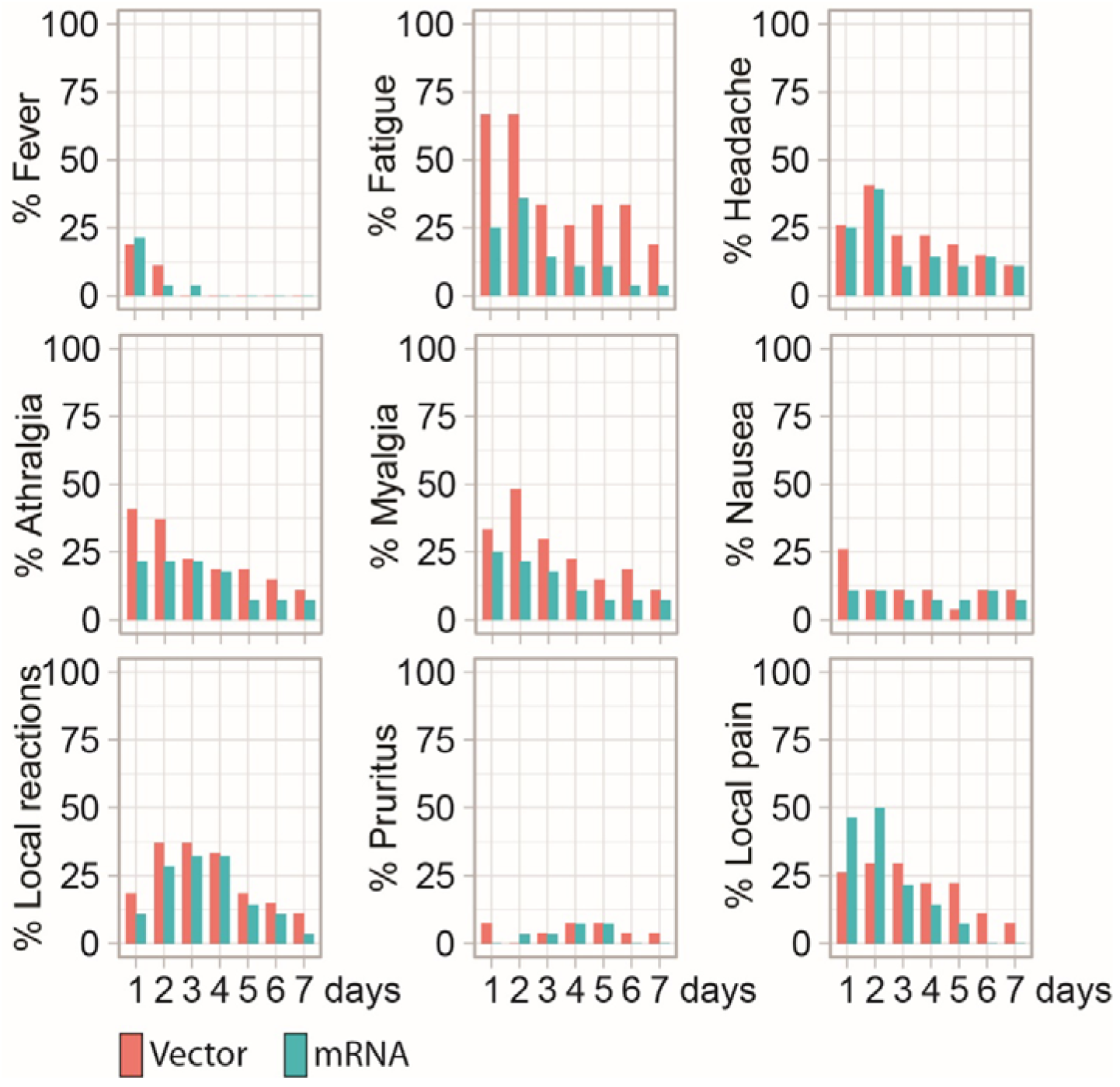
Safety. Systemic reactogenicity was evaluated daily during the first week after vaccination.

No thrombocytopenia or antibodies against platelet factor 4 (PF4) were observed after additional booster vaccination. None of the patients experienced any anaphylactoid reaction or neurological complication. Seven patients (13%) reported an alteration or worsening in their underlying disease one week after vaccination, but no disease flare that required glucocorticoid treatment or change in immunosuppressive medication was reported within the study period.

## DISCUSSION

In this randomized, controlled clinical trial, we enrolled patients treated with rituximab for an underlying autoimmune disease, who had not seroconverted upon vaccination with two doses of an mRNA vaccine, and thus continued to be at high risk for a severe disease course of SARS-CoV-2 infection. The additional SARS-CoV-2 booster vaccination evaluated in this trial resulted in the development of a humoral immune response in 27% of this initially vaccination-refractory patient population. Moreover, the additional booster vaccination reduced the proportion of patients lacking both a humoral and cellular immune response to primary vaccination from 31% to 6%.

Currently approved vector and mRNA vaccination strategies against SARS-CoV-2 consider only homologous vaccination. However, recent studies indicate a better humoral and cellular immune response after heterologous prime-boost vaccination in healthy individuals^16,18–21^. In our study, no significant advantage for either the homologous or heterologous vaccination strategy was found: the primary outcome showed a 10% higher seroconversion rate for mRNA (homologous) versus vector (heterologous) vaccination. Conversely, the inducibility of a T-cell response was numerically higher for the vector vaccine. However, while unlikely, these findings statistically cannot rule out a higher efficacy of an additional heterologous versus homologous booster vaccination.

To date, limited data exist that report on the efficacy and safety of a third vaccine in immunosuppressed patients to guide the vaccination strategy on non-seroconverted patients, particularly those at high risk for severe Covid-19 infections. Data published so far report on increased immunogenicity of a third vaccine in patients with solid organ transplantations^6–8^. However, most of the patients included in these trials had already shown some humoral response, as evidenced by the inclusion criteria, which allowed for the presence of low antibody levels against SARS-CoV-2 after two vaccinations. In contrast, none of the patients in our study had detectable anti-SARS-CoV-2 antibodies at baseline.

Detectable peripheral B-cells serve as a key factor for seroconversion in rituximab-treated patients^14^ and randomization was therefore stratified by presence or absence of peripheral B-cells. As described after conventional vaccination with two mRNA vaccine doses, presence of detectable peripheral B-cells was the strongest determinant for seroconversion also in patients receiving an additional booster vaccination. These data support the critical consideration of the timing of rituximab treatment, potentially suggesting postponing its application until after vaccination, or that vaccination should be timed after peripheral B-cells have repopulated. Which strategy may be preferable will be guided by the perceived severity of underlying disease as well as the risk from a severe COVID-19 infection.

The concern with such booster vaccination, also among candidate patients, may mostly relate to the risk of adverse reactions. Although no serious adverse events after booster vaccination were reported in both groups, our data show a numerically higher incidence of adverse events in patients boosted with the heterologous vector than with the homologous mRNA vaccine. These data are in line with recently published reports, which describe an increase in systemic reactogenicity in participants receiving heterologous schedules as compared to homologous schedules^17^. Reactogenicity was similar upon third vaccination as reported previously^16^. In our trial, typical general systemic reactions (like fever, myalgias, and similar) were observed, that were within the scope of the reports from the large approval studies ^22–24^, with some numerical differences seen between the two treatment groups.

One limitation of the trial is the absence of a placebo control, which was considered unethical in this high-risk population. While the small sample size precluded delivering ultimate statistical evidence concerning the clinical question of differences in immune responses to booster vaccination with homologous versus heterologous products, the important result of our study is that a third booster vaccination is effective in inducing an immune response in these refractory patients. Since we cannot generalize these data to the wider population of non-responders to COVID-19 vaccines, i.e. beyond immunosuppressed patients, broader population-based programs are needed to evaluate the impact of an additional booster vaccination in non-responding healthy individuals. It is important to note, that it still needs to be determined how humoral and cellular immune responses (or their absence) relate to protection against clinical infection with SARS-CoV-2.

Our data show that a cellular and/or humoral immune response can be achieved upon a third COVID-19 vaccination in most of the patients who initially developed neither a humoral nor a cellular immune response. The efficacy data together with the safety data seen in our trial provide a favorable risk/benefit ratio and support the implementation of a third vaccination for non-seroconverted high-risk autoimmune disease patients treated with B-cell-depleting agents. This might be a viable way to protect this group of patients from more dire consequences of an acquired SARS-CoV-2 infection.

## METHODS

### TRIAL DESIGN & POPULATION

In this prospective patient and efficacy (laboratory) blinded randomized controlled trial (EudraCT: 2021-002348-57), adults (≥ 18 years) with chronic-inflammatory rheumatic or neurologic diseases under current rituximab therapy and without detectable SARS-CoV-2 spike protein antibodies at least four weeks after their second standard vaccination with an mRNA vaccine (BNT162b2 or mRNA-1273) were included. Key exclusion criteria were previous infection with SARS-CoV-2 or known allergies to study compounds. The detailed inclusion/exclusion criteria can be found in the trial protocol **(Supplementary information)**.

Patients were block-randomized in a 1:1 ratio based on the presence or absence of peripheral B lymphocytes by a computerized randomization algorithm to receive either a third dose of an mRNA vaccine (BNT162b2 or mRNA-1273, respective of their initial vaccination compound) or a third vaccination with a vector COVID-19 vaccine (ChAdOx1 nCoV-19).

During the screening visit (visit one), data on demographics, concomitant medication, possible hypersensitivity reactions to the previous SARS-CoV-2 vaccination and medical history regarding SARS-CoV-2 infections were collected. The absence of detectable SARS-CoV-2 antibodies against nucleocapsid and spike-protein was verified before enrollment and the level of peripheral B lymphocytes was assessed. The vaccination was applied during a baseline visit (visit two, within 28 days after screening) followed by visits three and four (one and four weeks after vaccination, respectively) to determine the efficacy and safety of the third COVID-19 vaccination. Serum samples obtained during visits one, three and four were stored below -70°C at the Biobank of the Medical University of Vienna, a centralized facility for the preparation and storage of biomaterial with certified quality management (International Organization for Standardization (ISO) 9001:2015)^25^. Peripheral blood mononuclear cells (PBMCs) were isolated at screening and week one by density gradient centrifugation and stored in the vapor phase of liquid nitrogen until further use.

All patients were blinded throughout visit four, mainly to allow objectivity in safety assessment of the two strategies; blinding of vaccines was ensured by using pre-arranged dose aliquots in syringes without reference to the type used. The City of Vienna provided the vaccines for this study free of charge through the Central Pharmacy of the Vienna General Hospital. The study was conducted in following Good Clinical Practice guidelines and the Declaration of Helsinki. The study protocol and all relevant documents were approved by the competent authorities and the local ethical committee (vote number 1481/2021) of the Medical University of Vienna. All patients provided their written informed consent. The first patient was included on May 25, 2021 and the last patient finalized the 4-week follow-up on August 5, 2021.

### ASSESSMENTS

#### Quantification of CD19^+^ peripheral B-cells

Immunological phenotyping was performed by flow cytometry (FACSCanto II, Becton Dickinson, San Jose, California, USA) using the whole blood first stain and then lyse and wash method (Becton Dickinson). Lymphocyte subsets were characterized with a combination of the following monoclonal antibodies (all provided by Becton Dickinson): fluorescein isothiocyanate (FITC)-labelled anti-CD3, phycoerythrin (PE)-labelled anti-CD16^+^56^+^, peridinin-chlorophyll-protein (PerCP)-cy5.5-labelled anti-CD4, PE-Cy7-labelled anti-CD19, allophycocyanin (APC)-Cy7-labelled anti-CD8, V450-labelled anti-human leukocyte antigen (HLA)-DR, V500-labelled anti-CD45 and APC-labelled anti-CD14. Results were expressed as percentage of CD19^+^ B-cells among total lymphocytes.

#### Anti-SARS-CoV-2 antibody testing

The Elecsys Anti-SARS-CoV-2 S immunoassay was used for the quantitative determination of antibodies to the receptor-binding domain (RBD) of the viral spike (S) protein.^26,27^ The quantitation range is between 0.4 and 2500.0□BAU/mL (binding antibody units per milliliter). A value greater than 0.8 BAU/mL was considered as positive. Tests were performed on a Cobas e801 analyser (Roche Diagnostics, Rotkreuz, Switzerland) at the Department of Laboratory Medicine, Medical University of Vienna (certified acc. to ISO 9001:2015 and accredited acc. to ISO 15189:2012).

#### T-cell responses

For T-cell stimulation (see below), PepMix SARS-CoV-2 peptide pools were purchased from JPT (Berlin, Germany). The pools cover the entire sequences of the SARS-CoV-2 S protein and comprise 15-mer peptides overlapping by 11 amino acids (aa). The S peptides are split into two subpools S1 (aa 1–643) and S2 (aa 633– 1273). Peptides were dissolved in dimethyl sulfoxide and diluted in AIM-V medium for use in enzyme-linked immunosorbent spot (ELISpot) assays.

For ex vivo *T-cell IFN-γ ELISpot assay*, PBMCs from patients before and after the third vaccination were thawed and processed on the same day. A total of 1–2×10^5^ cells per well were incubated with SARS-CoV-2 peptides (2□µg/mL; duplicates), AIM-V medium (negative control; 3–4 wells) or phytohemagglutinin (PHA) (L4144, Sigma; 0,5□µg/mL; positive control) in 96-well plates coated with 1.5□µg anti-IFN-γ (1-D1K, Mabtech) for 24□hours. After washing, spots were developed with 0.1□µg biotin-conjugated anti-IFN-γ (7-B6-1, Mabtech), streptavidin-coupled alkaline phosphatase (Mabtech, 1:1000) and 5-bromo-4-chloro-3-indolyl phosphate/nitro blue tetrazolium (Sigma). Spots were counted using a Bio-Sys Bioreader 5000 Pro-S/BR177 and Bioreader software generation 10. Data were calculated as spot-forming cells (SFCs) per 10^6^ PBMCs after subtracting of the spots from the negative control (mean spot numbers from three to four unstimulated wells). T-cell responses were considered positive if spot counts were greater than the mean SFCs plus three times the standard deviation from pre-pandemic controls, as defined previously^14^.

### ENDPOINTS AND SAMPLE SIZE

The primary study endpoint was defined as difference in antibody seroconversion rates between the vector and mRNA vaccinated groups.

Secondary endpoints included seroconversion rate and SARS-CoV-2 antibody levels at week 4 overall and stratified for patients with and without detectable peripheral B-cells as well as cellular immune response defined by T lymphocyte restimulation potential before and one week after vaccination. Safety was reported and evaluated for incidence and severity of adverse events as well as potential effects on the underlying disease activity over a period of 28 days. Additionally, a paper-based patient diary was used. The study sample size was pragmatically targeted at 60 individuals, based on the number of rituximab-treated patients potentially eligible during the tight recruitment period, including estimates of non-responders to a standard protocol of mRNA vaccination, and expected participation rates. Based on a Chi-squared test comparing vector versus mRNA vaccine, this number of patients would allow to achieve at least 80% power at a minimal detectable difference of 28% (5% of responders in one group versus 33% in the other).

### STATISTICAL ANALYSIS

All subjects vaccinated with a third dose were included in the analysis. Primary outcome was assessed using Chi-squared test. Secondary outcomes and safety data are presented in a descriptive manner. Post hoc exploratory analyses were performed to evaluate factors associated with seroconversion rates by univariate logistic regression analyses. Variable selection was based on previous data in rituximab patients, and included age, concomitant medication, type of booster vaccination and presence or absence of detectable peripheral B-cells^14^. GraphPad Prism (version 9.1.0) was used for the graphical presentation of the data. “R” version 4.0.3 (R Development Core Team. Vienna, Austria) was used for the entire statistical analysis. Following packages were utilized: “ggplot2”, “ggbeeswarm” and “sjPlot” for creating plots and “tableone” to create baseline tables.

## Data Availability

Anonymous participant data is available under specific conditions. Proposals will be reviewed and approved by the sponsor, scientific committee, and staff on the basis of scientific merit and absence of competing interests. Once the proposal has been approved, data can be transferred through a secure online platform after the signing of a data access agreement and a confidentiality agreement.

## Acknowledgments

We thank all the patients who participated. We thank Martina Durechova, Daffodil Dioso and Michael Zauner for their support. We thank Brigitte Meyer, Birgit Niederreiter, Carl-Walter Steiner, Ursula Sinzinger, Amelie Popovitsch, Jutta Hutecek, Sebastian Weiss, Patrick Mucher, Astrid Radakovics, and Manuela Repl for their technical assistance. We thank Sylvia Taxer, Zoltan Vass, Eva Rath, Nikolaus Hommer, Lisa Göschl and Jochen Zwerina for their support. We thank Franz X. Heinz for critically reading the manuscript.

## Author contributions

All authors contributed to manuscript preparation. M.B., D.A., D.S., D.M., M.Z., ST and S.W. contributed to the study design. D.M., H.R., L.H. and D.S. contributed to data analysis. T.P., H.H. and K.S. performed antibody measurements. J.A., M.K., M.M. and P.H. contributed to cellular assays, M.B., D.A. J.S., D.M., D.S., L.H. and M.Z. contributed to manuscript preparation. S.T, D.M., M.B, P.M., B.K., E.S., R.F and E.H. contributed to patient recruitment, R.T. determined leucocyte subsets.

