## Supplementary material for "Additional heterologous versus homologous booster vaccination in immunosuppressed patients without SARS-CoV-2 antibody seroconversion after primary mRNA vaccination: a randomized controlled trial"

***Table of contents***

| ***Title*** | ***Page(s)*** |
| --- | --- |
| **Supplementary Table 1:** Patient characteristics stratified by seroconversion | 3 |
| **Supplementary Table 2:** Patient characteristics with assessed T-cell response | 4 |
| **Supplementary Table 3:** Integrative analysis of humoral and/or T-cell responses stratified by vector or mRNA vaccine. | 5 |
| **Supplementary Table 4:** Patient characteristics stratified by T-cell response | 6 |
| **Trial protocol v1** | 7 |
| **Trial protocol v2** | 37 |
| **Trial protocol v3** | 68 |
| **Trial protocol v4** | 100 |

**Supplementary Table 1:** Patient characteristics stratified by seroconversion

| Seroconversion | no | yes |
| --- | --- | --- |
| n | 40 | 15 |
| Age (mean (SD)) | 60.6 (16.7) | 58.0 (17.0) |
| Gender: female n (%) | 30 (75.0) | 11 (73.3) |
| **Diagnosis n (%)** |  |  |
| Arthritis | 18 (45.0) | 3 (20.0) |
| Connective tissue diseases | 12 (30.0) | 4 (26.7) |
| IgG4-related disease | 2 (5.0) | 2 (3.3) |
| Multiple sclerosis | 3 (7.5) | 3 (20.0) |
| Vasculitis | 5 (12.5) | 3 (20.0) |
| Months between RTX and screening (mean (SD)) | 6.04 (5.78) | 7.69 (1.41) |
| Weeks between 2^nd^ vaccination and screening (mean (SD)) | 7.80 (3.47) | 6.43 (1.62) |
| Patients with detectable B-cells: n (%) | 6 (15.0) | 12 (80.0) |
| Peripheral B-cells, percentage (median [IQR]) | 0.0 [0.0, 0.0] | 0.8 [0.4, 1.5] |
| **Concomitant medication, n (%)** |  |  |
| Any csDMARD | 23 (57.5) | 3 (20.0) |
| Methotrexate | 9 (22.5) | 1 (6.7) |
| Mycophenolate mofetil | 5 (12.5) | 1 (6.7) |
| Azathioprine | 4 (10.0) | 1 (6.7) |
| Leflunomide | 4 (10.0) | 0 (0.0) |
| Hydroxychloroquine | 4 (10.0) | 0 (0.0) |
| Immunoglobulin therapy | 1 (2.5) | 1 (6.7) |
| Prednisone | 12 (30.0) | 3 (20.0) |
| **Booster vaccination n (%)** |  |  |
| ChAdOx1 nCoV-19 | 21 (52.5) | 6 (40.0) |
| BNT162b2 | 15 (37.5) | 6 (40.0) |
| mRNA-1273 | 4 (10.0) | 3 (20.0) |
| SARS-Cov-2 Spike antibodies BAU/mL  (median [IQR]) | 0.0 [0.0, 0.0] | 15.7 [4.7, 25.8] |

csDMARD, conventional synthetic disease modifying antirheumatic drug, defined here as concomitant treatment with at least one of the following: methotrexate, mycophenolate mofetil, azathioprine, leflunomide, hydroxychloroquine.

**Supplementary Table 2:** Patient characteristics stratified by T-cell response

|  | Vector | mRNA |
| --- | --- | --- |
| n | 20 | 16 |
| Age (mean (SD)) | 56.6 (13.8) | 56.3 (20.0) |
| Gender: female n (%) | 15 (75.0) | 14 (87.5) |
| **Diagnosis n (%)** |  |  |
| Arthritis | 7 (35.0) | 6 (37.5) |
| Connective tissue diseases | 6 (30.0) | 5 (31.2) |
| IgG4-related disease | 1 (5.0) | 2 (12.5) |
| Multiple sclerosis | 3 (15.0) | 1 (6.2) |
| Vasculitis | 3 (15.0) | 2 (12.5) |
| Months between RTX and screening  (mean (SD)) | 6.6 (5.3) | 5.4 (4.0) |
| Weeks between 2^nd^ vaccination and screening (mean (SD)) | 7.2 (2.8) | 6.2 (1.9) |
| Detectable B-cells: n (%) | 6 (30.0) | 5 (31.2) |
| **Concomitant medication, n (%)** |  |  |
| Any csDMARD | 6 (30.0) | 10 (62.5) |
| Mycophenolate mofetil | 1 (5.0) | 2 (12.5) |
| Leflunomide | 3 (15.0) | 1 (6.2) |
| Hydroxychloroquine | 0 (0.0) | 3 (18.8) |
| Methotrexate | 2 (10.0) | 5 (31.2) |
| Azathioprine | 0 (0.0) | 1 (6.2) |
| Immunoglobulin therapy | 1 (5.0) | 1 (6.2) |
| Prednisone | 6 (30.0) | 6 (37.5) |
| Primary vaccination with mRNA-1273 n (%) | 3 (15.0) | 1 (6.2) |
| Seroconversion: n (%) | 5 (25.0) | 4 (25.0) |
| T-cell response at screening (median [IQR]) | 248 [54, 705] | 113 [33, 394] |
| T-cell response at week one (median [IQR]) | 459 [133, 722] | 305 [171, 416] |

csDMARD, conventional synthetic disease modifying antirheumatic drug, defined here as concomitant treatment with at least one of the following: methotrexate, mycophenolate mofetil, azathioprine, leflunomide, hydroxychloroquine.

**Supplementary Table 3:** Integrative analysis of humoral and/or T-cell responses stratified by vector or mRNA vaccine.

| pre Group | post Group | n (Vector) | n (mRNA) |
| --- | --- | --- | --- |
| AB (-), T (-) | AB (+), T (+) | 2 | 2 |
| AB (-), T (+) | AB (+), T (+) | 3 | 1 |
| AB (-), T (-) | AB (+), T (-) | 0 | 1 |
| AB (-), T (+) | AB (-), T (+) | 12 | 9 |
| AB (-), T (-) | AB (-), T (+) | 3 | 1 |
| AB (-), T (-) | AB (-), T (-) | 0 | 2 |

Pre-Group: Before third dose, Post-Group: after third dose, AB: Antibody, T: T-cell response, +: positive,-: negative, n (Vector): Number of patients in the vector group, n (mRNA): number of patients in the mRNA group.

**Supplementary Table 4:** Patient characteristics stratified by T-cell response

| T-cell response | no | yes |
| --- | --- | --- |
| n | 3 | 33 |
| Age (mean (SD)) | 59.7 (19.1) | 56.2 (16.7) |
| Gender: female n (%) | 3 (100) | 26 (79) |
| **Diagnosis n (%)** |  |  |
| Arthritis | 2 (66.7) | 11 (33.3) |
| Connective tissue diseases | 1 (33.3) | 10 (30.3) |
| Vasculitis | 0 (0.0) | 5 (15.2) |
| Multiple sclerosis | 0 (0.0) | 4 (12.1) |
| IgG4-related disease | 0 (0.0) | 3 (9.1) |
| Months between RTX and screening (mean (SD)) | 8.5 (2.1) | 5.8 (4.9) |
| Weeks between 2^nd^ vaccination and screening  (mean (SD)) | 7.24 (2.5) | 6.7 (2.5) |
| Patients with detectable B-cells: n (%) | 1 (33.3) | 10 (30.3) |
| **Concomitant medication, n (%)** |  |  |
| Any csDMARD | 3 (100.0) | 13 (39.4) |
| Methotrexate | 2 (66.7) | 5 (15.2) |
| Mycophenolate mofetil | 0 (0.0) | 3 (9.1) |
| Leflunomide | 0 (0.0) | 4 (12.1) |
| Hydroxychloroquine | 0 (0.0) | 3 (9.1) |
| Azathioprine | 1 (33.3) | 0 (0.0) |
| Immunoglobulin therapy | 0 (0.0) | 2 (6.1) |
| Prednisone | 2 (66.7) | 10 (30.3) |
| **Booster vaccination n (%)** |  |  |
| ChAdOx1 nCoV-19 | 0 (0.0) | 20 (60.6) |
| BNT162b2 | 2 (66.7) | 13 (39.4) |
| mRNA-1273 | 1 (33.3) | 0 (0.0) |
| Seroconversion: n (%) | 1 (33.3) | 8 (24.2) |
| T-cell response at week one (median [IQR]) | 20 [14, 31] | 389 [190, 675] |

csDMARD, conventional synthetic disease modifying antirheumatic drug, defined here as concomitant treatment with at least one of the following: methotrexate, mycophenolate mofetil, azathioprine, leflunomide, hydroxychloroquine.

### SARS-CoV-2 VAC3 study protocol Version 1.0, April 28^th^ 2021

### Clinical Study Protocol

A Randomized, Parallel Group, Single-Blind, Phase 2 Study to Evaluate the immune response of two classes of SARS-Cov-2 Vaccines employed as Third Vaccination in Patients under current Rituximab Therapy and no humoral response after standard mRNA vaccination

**Daniela Sieghart^1^, Selma Tobudic^2^, Martina Durechova^1^, Daniel Mrak^1^, Michael Bonelli^1^, Daniel Aletaha^1^**

**1** Department of Internal Medicine III, Division of Rheumatology, Medical University of Vienna, Austria

**2** Department of Medicine I, Division of Infectious Diseases and Tropical Medicine, Medical University of Vienna, Austria

**Confidentiality Statement**

The information contained in this document, especially unpublished data, is the property of the sponsor of this study. It is therefore provided to you in confidence as an Investigator, potential Investigator, or consultant, for review by you, your staff, and an Independent Ethics Committee or Institutional Review Board. It is understood that this information will not be disclosed to others without written authorization from the Sponsor except to the extent necessary to obtain informed consent from those persons to whom the study drug may be administered.

### 1. SPONSOR, INVESTIGATOR, MONITOR AND SIGNATURES

**Sponsor/or representative (OEL) (AMG §§ 2a, 31, 32)**

**Univ.-Prof. Dr.med.univ.** **Alexandra Kautzky-Willer,**

Department of Internal medicine III, Medical University of Vienna, Austria

Signature (OEL)	Date

**Coordinating Investigator (AMG §§ 2a, 35, 36**)

**Univ.-Prof. Dr.med.univ.** **Daniel Aletaha,**

Department of Internal Medicine III, Division of Rheumatology, Medical University of Vienna, Austria

Signature	Date

#### 2. **Study summary**

**Background.** Treatment with rituximab (RTX), a monoclonal antibody targeting CD20, constitutes an important therapeutic strategy for patients with several inflammatory rheumatic diseases. Some recent reports have already highlighted the risk of serious consequences upon severe acute respiratory syndrome coronavirus 2 (SARS-CoV-2) infection in patients treated with RTX. Besides the risk of a more severe disease course during B cell depleting therapy, a major concern is the immunogenicity of vaccination during immunomodulatory therapies, especially with RTX. Indeed, RTX has been shown to impair humoral responses to various vaccines including SARS-CoV-2 vaccination. Studies from other vaccines have shown that an additional vaccination or a change of the vaccine can lead to development of a humoral immune response.

**Objective.** To investigate if a third vaccination with a vector based vaccine is superior to a further dose of the vaccine used for basis immunization in previous non responders receiving rituximab.

**Methods.** We will perform a prospective single blind randomized controlled study. A total of 60 patients under rituximab treatment will be enrolled in this clinical trial who received two vaccinations with an mRNA vaccine. Four study visits per patient will be planned. During the screening visit antibodies to the receptor-binding domain will be determined. All patients without detectable humoral immunity against SARS-Cov2 will be invited for a second boost vaccination within 4 weeks after the screening visit. During the baseline visit patients will be randomized to receive a third vaccination with either an mRNA-SARS-CoV-2 vaccine (Biontech/Pfizer or Moderna – according to the vaccine used for their first two vaccinations) or a vector SARS-CoV-2 (AstraZeneca) vaccination as a second boost. Additional study visits are scheduled at weeks 1 and 4 for assessment of humoral and cellular immunity, as well as clinical signs of adverse effects of disease activity reactivation. Patients without humoral response at week 4 will be offered participation in an extended follow-up for additional 8 weeks (i.e. at week 8 and 12 after baseline)

**Expected Results.** We expect at least 30% of patients receiving a third SARS-CoV-2 vaccination to develop humoral response after 4 weeks. The change in mode of action of the vaccine might be beneficial to induce response in patients with rituximab.

##### **3. BACKGROUND**

The current pandemic caused by severe acute respiratory syndrome coronavirus‐2 (SARS‐CoV‐2) has led to exponentially rising morbidity and mortality worldwide. https://coronavirus.jhu.edu/map.html. Apart from aggressive quarantine and hygiene control measures, the most effective way to inhibit SARS‐CoV‐2 spread is a population‐wide vaccination campaign. The SARS-CoV-2 pandemic has required rapid action and the development of vaccines in an unprecedented timeframe. Data from the preclinical development of vaccine candidates for SARS-CoV and MERS-CoV enabled the initial step of experimental vaccine design to be essentially omitted, saving a considerable amount of time. In many cases, production processes were adapted from existing vaccines or vaccine candidates, and in some instances, preclinical and toxicology data from related vaccines could be used. As a result, the first clinical trial of a vaccine candidate for SARS-CoV-2 began in March 2020. Trials were designed such that clinical phases are overlapping and trial starts are staggered, with initial phase I/II trials followed by rapid progression to phase III trials after an interim analysis of the phase I/II data.

More than 180 vaccine candidates, based on several different platforms, are currently in development against SARS-CoV-2.^1^ The World Health Organization (WHO) maintains a working document that includes most of the vaccines in development and is available at https://www.who.int/publications/m/item/draft-landscape-of-covid-19-candidate-vaccines.

RNA vaccines are a relatively recent development. The genetic information for the antigen is delivered instead of the antigen itself. The antigen is then expressed in the vaccinated individual cells. ^2^ Either mRNA (with modifications) or a self-replicating RNA can be used. Higher doses are required for mRNA than for self-replicating RNA, and the RNA is usually delivered via lipid nanoparticles (LNPs).^2-5^

Two mRNA vaccines, BNT162b2, developed by BioNTech/Fosun Pharma/Pfizer, and mRNA-1273 developed by Moderna/NIAID, are currently in phase III trials. ^6^^,^ ^7^

In December 2020, BNT162b2 was under evaluation for emergency use authorization (EUA) for widespread use by several medical regulators globally. On 21 December, the European Medicines Agency (EMA) has given official approval for the BNT162b2 vaccine developed by BioNTech and Pfizer.

On 18 December 2020, mRNA-1273 developed by Moderna/NIAID was issued an emergency use authorization by the United States Food and Drug Administration. On 06 January, the EMA has given official approval for the mRNA-1273, developed by Moderna/NIAID.

Preliminary results from Phase I–II clinical trials on BNT162b2, published in October 2020, indicated the potential for its efficacy and safety. ^8^ ^9^ Findings from studies conducted in the United States and Germany among healthy men and women showed that two 30-μg doses of BNT162b2 elicited high SARS-CoV-2 neutralizing antibody titers and robust antigen-specific CD8+ and Th1-type CD4+ T-cell response. ^10^ Besides, the reactogenicity profile of BNT162b2 represented mainly short-term local (i.e., injection site) and systemic responses. These findings supported the progression of BNT162b2 into the next phase. In phase III, a total of 43548 participants underwent randomization, of whom 43448 received injections: 21720 with BNT162b2 and 21728 with placebo. There were 8 cases of COVID-19 with onset at least 7 days after the second dose among participants assigned to receive BNT162b2 and 162 cases among those assigned to placebo. BNT162b2 was 95% effective in preventing COVID-19 (95% credible interval, 90.3 to 97.6). Similar vaccine efficacy (generally 90 to 100%) was observed across subgroups defined by age, sex, race, ethnicity, baseline body-mass index, and the presence of coexisting conditions.

In July 2020, preliminary results of the Phase I dose-escalation clinical trial of mRNA-1273 were published, showing dose-dependent induction of neutralizing antibodies against SARS-CoV-2 as early as 15 days post-injection. 6 Mild to moderate adverse reactions, such as fever, fatigue, headache, myalgia, and pain at the injection site, were observed in all dose groups but were common with increased dosage. The vaccine in low doses was deemed safe and effective to advance a Phase III clinical trial using two 100-μg doses administered 29 days apart.

In a Phase III trial of mRNA-1273 conducted in the United States between July and October, enrolled and assigned 30 000 volunteers to two groups — one group receiving two 100-μg doses of mRNA-1273 vaccine and the other receiving a placebo of 0.9% sodium chloride. Preliminary data from Phase III clinical trial, published in November 2020, indicating 94% efficacy in preventing COVID-19 infection. Side effects included flu-like symptoms, such as pain at the injection site, fatigue, muscle pain, and headache. The Moderna results were not final – as the trial is not scheduled to conclude until late-2022– and were not peer-reviewed or published in a medical journal.

The efficacy of ChAdOx1 nCoV-19 vaccine was reported from phase III trials in the United Kingdom and Brazil.^11^ The interpretation of the results of these trials is complicated by a dosing error (in which some participants unintentionally received a half-dose for their first of two doses), a small number of participants, and differences in efficacy in the two countries. The overall efficacy in preventing symptomatic infection more than 14 days after the second dose was 70.4% (95%CI: 54.8%-80.6%), with efficacy of 62.1% (95%CI: 41.0%-75.7%) in those who received standard doses, and 90.0% (95%CI 67.4% to 97.0%) in those who received a half-dose followed by a standard dose. Notably, the lower efficacy was based on results from the study in Brazil, and higher efficacy was reported for the study in the United Kingdom. Hospitalizations and severe COVID-19 occurred rarely but exclusively in the placebo arm of these trials. Due to varying intervals between the first and second doses, vaccine efficacy after a single standard dose from day 22 to day 90 was modeled and estimated to be 76% (95%CI: 59%-86%), with maintenance of antibody levels up to day 90. Furthermore, vaccine efficacy appeared to be 82.4% (95%CI: 62.7%-91.7%) when the interval between doses was more than 12 weeks compared to 54.9% (95%CI: 32.7%-69.7%) when the interval was less than 6 weeks. Similarly, geometric mean antibody levels were higher with a longer prime-boost interval in those age 18-55 years.^12^ A larger phase III trial using two standard doses 28 days apart with a majority of the participants in the US recently completed enrollment. Preliminary results of this trial showed vaccine efficacy of 76% (95%CI: 68-82%) at preventing symptomatic infection and 100% efficacy at preventing severe or critical disease and hospitalization. Vaccine efficacy was consistent across ethnicity and age. TheChAdOx1nCoV-19 vaccine has received EUA from the United Kingdom and the European Union. In the United Kingdom, a single-blind multi-center randomized phase II/III trial of the ChAdOx1 nCOV-19 vaccine asked participants to provide a weekly self-administered nose and throat swab starting one week after administration of the first vaccine (or placebo). This study revealed that among those who were infected, vaccinated persons had lower peak viral load and shorter duration of RT-PCR-positive results for SARS-CoV-2 compared to controls, suggesting that the vaccine is effective in reducing transmission.^13^

Patients with immunosuppression with COVID-19 may be at higher risk of hospitalization and ICU admission than matched comparators.^14^ An mRNA vaccine vaccination should be performed as primary immunization with two vaccine-doses 21 or 28 days apart. There are no data about the efficacy and safety of mRNA vaccine against SARS-CoV-2 in patients with a chronic immune-mediated disease such as rheumatoid arthritis, certain kinds of vasculitis, and lupus. However, after vaccination, immune responses might vary among immunosuppressed patients, dependent upon underlying clinical conditions and the level of immunosuppression. Moreover, it is not clear whether mRNA vaccination against SARS-CoV-2 in the immunosuppressed has a protective effect at all. Specifically, B-cell depleting therapies like CD20 antagonists rituximab have a high risk of vaccination failure.^15^ As for other vaccines, the efficacy of a COVID-19 vaccine may be reduced in patients treated with rituximab. For patients treated with rituximab, it is preferable to administer the vaccine at least 6 months after the last infusion^16^ to have the highest possible number of restored B-cells. The current treatment scheme for rituximab is 500mg or 1000mg intravenously every 6 months.

However, the vaccine is not contraindicated in this population and the treating physician should discuss with the patient the appropriate timing of vaccination taking into consideration the context of pandemic, the risk of poor COVID-19 disease prognosis, and a possibly reduced humoral response to vaccination under rituximab treatment.

**4. STUDY RATIONALE**

Patients who had received rituximab are at high risk of non-response to SARS-CoV-2 vaccination. It is unclear whether patients who did not develop humoral immunity after a standard protocol application with a mRNA vaccine would benefit from a second boost of mRNA SARS-CoV-2 or from a single additional shot of vector SARS-CoV-2 vaccine.

**5. STUDY OBJECTIVES**

The study aims to investigate the humoral and cellular immune responses after a second boost vaccination against SARS-CoV-2 in adult patients treated with rituximab (anti B cell therapy) who did not show response to the first two vaccinations with an mRNA vaccine.

**5.1. Primary Objective (Hypothesis)**

To assess the immunogenicity to a third vaccination mRNA-SARS-CoV-2 vaccine (Biontech/Pfizer or Moderna) compared to a vector SARS-CoV-2 (AstraZeneca) vaccination as a second boost in patients with rituximab by measuring quantitative antibody levels by enzyme-linked immunosorbent assay test (ELISA) and neutralization test (NT) or pseudo viral neutralization assay.

**Null and alternative hypotheses:**

H0: There is no statistical difference in the seroconversion rate between patients receiving a third mRNA vaccination and the patients receiving a second boost with AstraZeneca.

H1: There is statistical difference in the seroconversion rate between patients receiving a third mRNA vaccination and the patients receiving a second boost with AstraZeneca.

**Secondary Objectives**

- To compare cellular immunogenicity of the third mRNA SARS-Cov-2 vaccination will be compared to patients receiving AstraZeneca vaccination as second boost in immunosuppressed patients. T cell proliferation will be assessed, and T-cell cytokine expression will be measured using flow-cytometry following *in vitro* stimulation of peripheral blood mononuclear cells (PBMCs) with SARS-Cov-2 specific antigens.
- To assess safety of a second boost vaccination.
- To evaluate the influence of vaccination on underlying disease activity.

**6. STUDY DESIGN**

A prospective single blind randomized controlled study will be performed. A total of 60 patients with B-cell depleting therapy (rituximab) will be enrolled in this clinical trial. Four study visits per patient will be planned.

After inclusion of the patients (after receiving their written informed consent) serum samples for determination of antibody levels will be obtained at least 4 weeks after the second mRNA vaccination (screening visit). All patients without detectable humoral immunity against SARS-Cov2 will be invited for a second boost vaccination within 4 weeks after the screening visit. Additional study visits are scheduled at baseline (vaccination), and at weeks 1 and 4 for assessment of humoral and cellular immunity, as well as clinical signs of adverse effects of disease activity reactivation. Patients without humoral response at week 4 will be offered participation in an extended follow-up for additional 8 weeks (i.e. at week 8 and 12 after baseline)

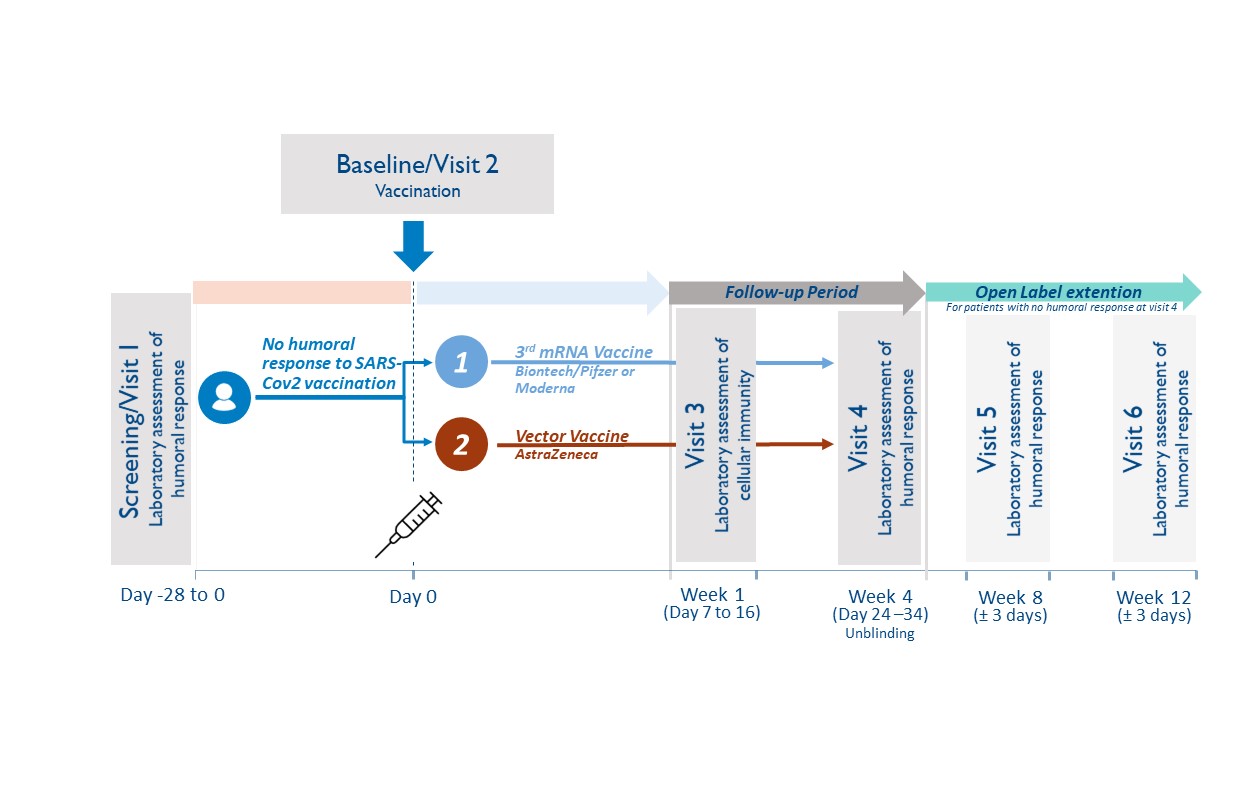

**6.1. Study population**

Adult Patient (≥ 18 years) under current rituximab therapy who did not develop humoral response against SARS-CoV-2 after their standard vaccination with Biontech/Pfizer or Moderna will be recruited at the Division of Rheumatology, Medical University of Vienna.

Study entry is defined as the date of signature of the study participant (subject) on the informed consent form. All subjects enrolled will be assigned a subject code, consisting of a consecutive subject number (2 digits).

**6.2. Inclusion criteria**

Male and female subjects will be eligible for participation in this study if they:

1. Are ≥18 years on the day of screening
2. Have a rheumatic condition and have been treated with a B-cell depleting therapy (rituximab) within the last 12 months
3. Received two doses of mRNA SARS-CoV-2 (Biontech/Pfizer or Moderna) vaccine according to recommendations in the label and/or national guidelines.
4. Did not develop humoral immunity 4 weeks after second mRNA vaccination to SARS-CoV-2 (analyzed during the study “Characterization of immune responsiveness after mRNA SARS-CoV-2 Vaccination in patients with immunodeficiency or immunosuppressive therapy”, EK-Nr. 1073/2021, EudraCT Nr. 2021-000291-11)
5. A maximum of 6 months after second vaccination
6. Have an understanding of the study, agree to its provisions, and give written informed consent before study entry
7. If female and capable of bearing children – have a negative urine pregnancy test result at study entry and agree to employ adequate birth control measures for the duration of the study

**6.3.	Exclusion criteria**

Subjects will be excluded from participation in this study if they:

1. Have shown humoral response to the SARS-CoV-2 vaccination
2. Had grade 3 adverse effects from the mRNA vaccination reported
3. Pregnancy and breast feeding
4. Signs of SARS-CoV-2 infection (including previous positive PCR testing)
5. Any other contraindication to any of the study compounds
6. Urgent need for next rituximab application

**6.4.	Study duration**

For the individual study participant, the active blinded study phase will be 4 weeks, with possibility to enroll in an open label extension for additional 8 weeks.

**6.5.	Randomization Procedure**

The web-based computerized randomization algorithm “Randomizer for Clinical Trials” by the Medical University of Vienna will be used for randomization. Patients will be randomized in a 1:1 ratio between the third dose mRNA SARS-CoV-2 boost (Biontech/Pfizer or Moderna, respective of their initial vaccination compound) and a single dose boost vector SARS-CoV-2 vaccine (AstraZeneca). Randomization will be stratified by presence or absence of B-cell depletion (determined at screening visit).

**6.6.1.	Blinding**

All patients will be blinded. Blinding of initial treatment allocation will be maintained throughout the 4 week study period. Blinding is ensured during the routine vaccination protocol at the General Hospital Vienna, where patients will see the pre-arranged dose aliquots in syringes without reference to the type of vaccine. To facilitate the process of vaccination of full vials, patients randomized to the same vaccine will be scheduled on the same day. Blinding will be mainly performed to prevent selective drop-outs due to knowledge of treatment allocation.

**6.6.2.	Unblinding Process**

The blinding can be lifted at the request of an investigator, when knowledge of the treatment is essential for appropriate patient management (e.g. after trial termination). In case of an emergency, the principal investigator has the sole responsibility for determining if unblinding of a patient´s treatment assignment is warranted.

If any serious adverse event arises during the study and unblinding appears necessary, the subinvestigator will notify the principal Investigator within two days in maximum. In the following the matter will be discussed and may lead to unblinding.

**6.7.1	Withdrawal and replacement of subjects**

Criteria for withdrawal:

Subjects may prematurely discontinue from the study at any time. The study's premature discontinuation is to be understood when the subject did not undergo complete the last visit (study visit 4) or all pivotal assessments during the study.

Subjects must be withdrawn under the following circumstances:

•	at their own request

•	if the investigator feels it would not be in the best interest of the subject to continue

•	if the subject violates conditions laid out in the consent form/information sheet or disregards instructions by the study personal

In all cases, the reason why subjects are withdrawn must be recorded in detail in the CRF and the subject's medical records. Should the study be discontinued prematurely, all study materials (complete, partially completed, and empty CRFs) will be retained.

Follow-up of patients withdrawn from the study:

In case of premature discontinuation after the start of vaccination, no further investigations concerning the study will be performed. Furthermore, participants may request that no more data will be recorded from the time point of withdrawal and that all biological samples collected in the course of the study will be destroyed.

**6.7.2	Premature termination of the study**

The sponsor has the right to close this study at any time. The IEC and the competent regulatory authority must be informed within 15 days of early termination.

The trial or single-dose steps will be terminated prematurely in the following cases:

- If the number of drop-outs is so high that proper completion of the trial cannot realistically be expected.
- Recruitment is not reaching the critical number

#### 6.8. **Adverse events and reporting**

##### a. Definition of adverse events

An adverse event is any untoward medical occurrence in a patient or clinical investigation subject administered a pharmaceutical product and which does not necessarily have a causal relationship with this treatment. An adverse event (AE) can therefore be any unfavourable and unintended sign (including an abnormal laboratory finding), symptom, or disease temporally associated with the use of a medicinal (investigational) product, whether or not related to the medicinal (investigational) product (see the ICH Guideline for Clinical Safety Data Management: Definitions and Standards for Expedited Reporting)

Adverse events include:

- Exacerbation of a pre-existing disease.
- Increase in frequency or intensity of a pre-existing episodic disease or medical condition.
- Disease or medical condition detected or diagnosed after study drug administration even though it may have been present prior to the start of the study.
- Continuous persistent disease or symptoms present at baseline that worsen following the start of the study.
- Lack of efficacy in the acute treatment of a life-threatening disease.
- Events considered by the Investigator to be related to study-mandated procedures.
- Abnormal assessments, e.g., ECG and physical examination findings, must be reported as AEs if they represent a clinically significant finding that was not present at baseline or worsened during the course of the study.
- Laboratory test abnormalities must be reported as AEs if they represent a clinically significant finding, symptomatic or not, which was not present at baseline or worsened during the course of the study or led to dose reduction, interruption or permanent discontinuation of study drug.

Adverse events do not include:

- Pre-planned interventions or occurrence of endpoints specified in the study protocol are not considered AE´s, if not defined otherwise (eg.as a result of overdose)
- Medical or surgical procedure, e.g., surgery, endoscopy, tooth extraction, transfusion. However, the event leading to the procedure is an AE. If this event is serious, the procedure must be described in the SAE narrative.
- Pre-existing disease or medical condition that does not worsen.
- Situations in which an adverse change did not occur, e.g., hospitalisations for cosmetic elective surgery or for social and/or convenience reasons.

Overdose of either study drug or concomitant medication without any signs or symptoms. However, overdose must be mentioned in the Study Drug Log.

##### b. Definition of serious adverse events (SAEs)

A Serious Adverse Event (SAE) is defined by the International Conference on Harmonization (ICH) guidelines as any AE fulfilling at least one of the following criteria:

- Fatal (including fetal death).
- Life-threatening – defined as an event in which the subject was, in the judgment of the investigator, at risk of death at the time of the event; it does not refer to an event that hypothetically might have caused death had it been more severe.
- Requiring subject's hospitalization or prolongation of existing hospitalization – inpatient hospitalization refers to any inpatient admission, regardless of length of stay.
- Resulting in persistent or significant disability or incapacity (i.e., a substantial disruption of a person’s ability to conduct normal life functions).
- Congenital anomaly or birth defect.
- Is medically significant or requires intervention to prevent at least one of the outcomes listed above.

In case of any SAE, the investigator has to use all means to ensure patient´s safety. Every SAE has to be reported as outlined in section 16.8.

##### c. Definition of suspected or unexpected serious adverse reactions (SUSARs)

SUSARs are all suspected adverse reactions related to the study drug that are both unexpected (not previously described in the SmPC or Investigator’s brochure) and serious.

##### d. Pregnancy

Maternal pregnancies must be reported to the principal investigator/sponsor. To ensure subject safety, each pregnancy must be reported to the PI/sponsor within 2 weeks of learning of its occurrence.

Pregnancies, should be followed up and reported to the PI/sponsor until the outcome of the pregnancy (including premature termination) and status of mother and child is known.

##### e. Severity of adverse events

The severity of clinical AEs is graded on a three-point scale: mild, moderate, severe, and reported on specific AE pages of the CRF.

If the severity of an AE worsens during study drug administration, only the worst intensity should be reported on the AE page. If the AE lessens in intensity, no change in the severity is required:

- *Mild* - Event may be noticeable to subject; does not influence daily activities; the AE resolves spontaneously or may require minimal therapeutic intervention;
- *Moderate* - Event may make subject uncomfortable; performance of daily activities may be influenced; intervention may be needed; the AE produces no sequelae.
- *Severe* - Event may cause noticeable discomfort; usually interferes with daily activities; subject may not be able to continue in the study; the AE produces sequelae, which require prolonged therapeutic intervention.

A mild, moderate or severe AE may or may not be serious.

##### f. Relationship of adverse events to study drug

For all AEs, the investigator will assess the causal relationship between the study drug and the AE using his/her clinical expertise and judgment according to the following algorithm that best fits the circumstances of the AE:

**Not related**

- May or may not follow a temporal sequence from administration of the study product
- Is biologically implausible and does not follow known response pattern to the suspect study drug (if response pattern is previously known).
- Can be explained by the known characteristics of the subject’s clinical state or other modes of therapy administered to the subject.

**Unlikely**

- There is a reasonable temporal relation between the AE and the intake of the study medication, but there is a plausible other explanation for the occurrence of the AE.

**Possibly**

- The AE has a reasonable temporal relationship with drug administration.
- The AE may equally be explained by the study subject’s clinically state, environmental or toxic factors, or concomitant therapy administered to the study subject.
- The relationship between study drug and AE may also be pharmacologically or clinically plausible.

**Probably**

- There is a reasonable temporal relation between the AE and the intake of the study medication, and plausible reasons point to a causal relation with the study medication.

**Related**

- Reasonable temporal relation between the AE and the intake of the study medication and
- There is no other explanation for the AE and
- Subsidence or disappearance of the AE on withdrawal of the study medication and
- Recurrence of the symptoms on restart at previous dose (only applies for re-institution of mediation).

**Not assessable**

The causal relationship between the study drug and the AE cannot be judged.

##### g. Adverse events reporting procedures

A special section is designated to adverse events in the case report form. For each subject, adverse events occurring after signing the informed consent must be recorded on the applicable adverse events page(s) in the case report form. Recording should be done in a concise manner using standard, acceptable medical terms. The adverse event recorded should not be a procedure or a clinical measurement (i.e., a laboratory value or vital sign) but should reflect the reason for the procedure or the diagnosis based on the abnormal measurement. If, in the investigator’s judgment, a clinically significant worsening from baseline is observed in laboratory or other parameters, physical exam finding, or vital sign, a corresponding clinical adverse event should be recorded on the adverse event page(s) of the CRF. If a specific medical diagnosis has been made that diagnosis should be recorded on the adverse event page(s) of the CRF.

SAEs and any newly identified pregnancy (maternal or paternal exposure), malignancy, overdose, opportunistic infection or case of active TB occurring after first administration of study agent in subjects participating in this clinical trial requires submission of a Safety Report Form to the Sponsor (only if other sites are reporting SAE) within 24 hours of notification or observation.

*The following details must thereby be entered:*

- Type of adverse event
- Start (date and time)
- End (date and time)
- Severity (mild, moderate, severe)
- Serious (no / yes)
- Unexpected (no / yes)
- Outcome (resolved, ongoing, ongoing – improved, ongoing – worsening)
- Relation to study drug (unrelated, possibly related, definitely related)

Adverse events are to be documented in the case report form in accordance with the above mentioned criteria.

##### h. Reporting procedures for SAEs

In the event of serious, the investigator has to use all supportive measures for best patient treatment. A serious adverse event must be reported if it occurs during a subject’s participation in the study (whether receiving study agent or not) or within six (6) months of receiving the last dose of study agent.

A Safety Report Form must be completed and faxed to the Sponsor (+43 (0)1 40400 43060) 24 hours of observation or notification of the event. The sponsor will be responsible for potential SUSAR assessment (see below) and reporting SAEs including potential SUSAR to the manufacturing company. A written report is also to be prepared and made available to the clinical investigator within five days.

The following details should at least be available:

- Patient initials and number
- Patient: date of birth, sex, ethical origin
- The suspected investigational medical product (IMP)
- The adverse event assessed as serious
- Short description of the event and outcome

Any serious adverse event that is ongoing when a subject completes his/her participation in the trial must be followed until any of the following occurs:

- The event resolves or stabilizes
- The event returns to baseline condition or value (if a baseline value is available)
- The event can be attributed to agent(s) other than the study agent, or to factors unrelated to study conduct

##### i. Reporting procedures for SUSARs

All SAEs will be evaluated regarding a possible classification as SUSAR by the sponsor, who will then perform all necessary reports to the manufacturing/distributing pharmaceutical company and forward the SUSAR to the CRO. The CRO will be responsible for reporting SUSARs to the regulatory authorities. In addition, due to their possible safety concern for the study participants, all SUSARs need to be reported to the Institutional Review Board / Independent Ethics Committee (IRB / IEC).

These reports are time critical and should be done within a maximum of 7 days (fatal or life threatening outcome) or 15 days (non fatal, not life threatening). The representative of the Sponsor Investigator shall inform all investigators concerned of relevant information about Serious Unexpected Suspected Adverse Reactions that could adversely affect the safety of subjects.

Such reports shall be made by the study management and the following details should be at least available:

- Patient inclusion number
- Patient: year of birth, sex, ethical origin
- Name of investigator and investigating site
- Period of administration
- The suspected investigational medical product (IMP)
- The adverse event assessed as serious and unexpected, and for which there is a reasonable suspected causal relationship to the IMP
- Concomitant disease and medication
- Short description of the event:
  - Description
  - Onset and if applicable, end
  - Therapeutic intervention
  - Causal relationship
  - Hospitalization of prolongation of hospitalization

7. **METHODOLOGY**

**7.1 Study design and blood samples**

A randomized controlled single-blind phase II trial will be performed. A total of 60 patients with current rituximab therapy who did not develop humoral immune response to SARS-CoV-2 after vaccination with mRNA SARS-CoV-2 vaccine (Biontech/Pfizer or Moderna). Four study visits per patient will be planned during the course of the trial: screening visit (-0-14 days), baseline, week 1, week 4. Patients will be unblinded at week 4 and a follow-up visit 12 weeks after vaccination in an open label approach. In addition, patients whio did not show humoral response at week 4 will be invited for an additional study visit at week 8. Vaccination will take place at baseline visit.

Blood samples for the determination of SARS-CoV-2 antibodies will be obtained at screening visit, at week 1, and week 4 and week 12.

**7.2	Study procedures**

**7.2.1	General rules for trial procedures**

•	All study measures like blood sampling and measurements have to be documented with the date (dd:mm:yyyy).

•	In case several study procedures are scheduled simultaneously, there is no specific sequence that should be followed.

•	The dates of all procedures should be according to the protocol. The time margins mentioned in the study flow chart are admissible. If for any reason, a study procedure is not performed within scheduled margins, a protocol deviation should be noted, and the procedure should be performed as soon as possible or as adequate.

•	If it is necessary for organizational reasons, it is admissible to perform procedures scheduled for one visit at two different time points. Allowed time margins should thereby not be exceeded.

**7.2.2	Study visits**

**7.2.2.1 Screening visit (visit 1, -1-28 days, at least 4 weeks after the second immunization with mRNA SARS-CoV-2 vaccine)**

The investigator will inform the subject about the procedures, risks, and benefits of the study. Participants will be informed about future visits, which will be synchronized with the appointment for the third vaccination.

Fully informed, written consent must be obtained from each subject before any assessment is performed. The subject must be allowed sufficient time to consider his/her participation in the study.

The following assessments will be performed:

- Inclusion and exclusion criteria
- Demographic data, including sex, age, weight, and height
- Medical history and concomitant medication
- Assessment of adverse reactions after the mRNA SARS-CoV-2 vaccination
- Assessment of recent COVID-19 disease
- Blood will be drawn:
  - 16 ml of blood will be drawn for PBMC isolation and determination of cellular immunity
  - 8 ml of blood will be drawn for SARS-Cov-2 antibody level detection (prior vaccination).
  - 8 ml of blood will be drawn for determination of leukocyte subpopulations
  - 8 ml of blood will be drawn for biobanking of serum
  - 20 ml of blood will be drawn for routine laboratory testing: blood count, chemistry, coagulation factors
- Pregnancy test in women with childbearing potential
- Randomization to one of the three vaccines: mRNA SARS-CoV-2 (Biontech/Pfizer or Moderna) or vector SARS-CoV-2 vaccination (AstraZeneca)
- SARS-CoV-2 vaccine appointment

**7.2.2.2. Baseline visit (visit 2, week 0, max. 28 days after visit 1)**

The following activities will be performed:

- The investigator will ask all subjects about any adverse experiences occurring since Visit 1 (screening). All adverse experiences will be documented in the CRF.
- Patients will receive a patients diary, fever thermometer and spacer
- Application of mRNA SARS-CoV-2 (Biontech/Pfizer or Moderna) or vector SARS-CoV-2 vaccine (AstraZeneca)

**7.2.2.3. Visit 3 (1 week after the third SARS-CoV-2 vaccination)**

The following activities will be performed:

- The investigator will ask all subjects about any adverse experiences occurring since Visit 2. All adverse experiences will be documented in the CRF.
- Patient´s diaries will be discussed
- Blood will be drawn:
  - 16 ml of blood will be drawn for PBMC isolation and determination of cellular immunity
  - 8 ml of blood will be drawn for SARS-Cov-2 antibody level detection
  - 6 ml of blood will be drawn for determination of leukocyte subpopulations
  - 8 ml of blood will be drawn for biobanking of serum
  - 20 ml of blood will be drawn for routine laboratory testing: blood account, chemistry, coagulation factors
  - 8 ml blood will be drawn for detection of anti-PDE4D antibody

**7.2.2. 4. Visit 4 (4 weeks after the third SARS-CoV-2 vaccination)**

The following activities will be performed:

•	The investigator will ask all subjects about any adverse experiences occurring since Visit 3. All adverse experiences will be documented in the CRF.

- Patient´s diary will be returned to investigator

•	Blood draw:

- 8 ml of blood will be drawn for SARS-Cov-2 antibody level detection
- 6 ml of blood will be drawn for determination of leukocyte subpopulations
- 8 ml of blood will be drawn for biobanking of serum
- 20 ml of blood will be drawn for routine laboratory testing: blood account, chemistry, coagulation factors
- 8 ml blood will be drawn for detection of anti-PDE4D antibody

**7.2.2.5. Visits 5 (week 8) and 6 (week 12) (only for patients enrolling in open label extension)**

The following activities will be performed:

•	The investigator will ask all subjects about any adverse experiences occurring since Visit 3. All adverse experiences will be documented in the CRF.

- Blood draw:
  - 8 ml of blood will be drawn for SARS-Cov-2 antibody level detection
  - 8 ml of blood will be drawn for biobanking of serum

**7.2.3	Determination of the humoral and cellular immunity**

**7.2.3.1 Determination of serum antibodies against vaccination antigens**

Analysis will be performed at the Department of Laboratory Medicine, Medical University of Vienna. All procedures will be carried out according to standard operating procedures in an ISO 9001:2015 certified environment (ref: 10.1089/bio.2018.0032).

Previous SARS-CoV-2 infection will be detected using nucleocapsid-based chemiluminescence assays (e.g., Roche SARS-CoV-2 NC total antibody ECLIA, Abbott SARS-CoV-2 NC IgG CMIA). Vaccination response will be assessed by spike-protein-based assays (e.g., Technozym RBD ELISA, Siemens RBD immunoassay, Roche SARS-CoV-2 RBD ECLIA).

All analyses will be carried out at the Department of Laboratory Medicine, Medical University of Vienna.

Neutralization assays will be performed as described earlier.^13^

**7.2.3.2 Determination of cellular immune response**

To investigate cellular immunity following SARS-CoV-2 vaccination Peripheral blood mononuclear cells (PBMCs) will be isolated from heparinized venous blood by density gradient centrifugation at 400 *g* of heparinized blood over LSM 1077 Lymphocyte Separation Medium, cryopreserved and stored in liquid nitrogen for later use.

**7.3	Study endpoints**

**7.3.1	Primary study endpoint**

Difference in SARS-CoV-2 antibody seroconversion rate by week 4 after vaccination boost at baseline between 3^rd^ mRNA SARS-CoV-2 (Biontech/Pfizer or Moderna) and vector SARS-CoV-2 vaccine (AstraZeneca).

**7.3.2	Secondary study endpoints**

The secondary endpoints of this study are:

- Overall SARS-Cov-2 antibody seroconversion rate by week 4 after vaccination boost at baseline
- Difference in overall SARS-Cov-2 antibody seroconversion rate by week 4 after vaccination boost at baseline between patient with and without B-cell repopulation.
- Antibody concentrations of SARS-CoV-2 ELISA 4 weeks after vaccination boost at baseline.
- Effect of immunosuppressive comedication on SARS-Cov-2 antibody seroconversion rate by week 4 after vaccination boost at baseline
- Evaluation of cellular immunity before and one week after the vaccination.
- Safety of vaccination boost.

#### 8. **ETHICAL CONSIDERATIONS**

##### a.  Ethical Review

All relevant documents must be reviewed and approved by the Ethics Committee and additionally submitted to the relevant authorities in accordance with the guidance of submission and conduct of clinical trials.

The clinical trial shall be performed in full compliance with the legal regulations according to the Drug Law (AMG - Arzneimittelgesetz) of the Republic of Austria.

An application must also be submitted to the Austrian Competent Authorities (Bundesamt für Sicherheit im Gesundheitswesen (BASG) represented by the Agency for Health and Food Safety (AGES PharmMed), and registered to the European Clinical Trial Database (EudraCT) using the required forms. The timelines for (silent) approval set by national law must be followed before starting the study.

##### b. Consent Procedures

After a detailed information about the study procedures and study medication, as well as the potentially related risks and benefits, the written informed consent will be obtained from each participant by the principal investigator or a designee. At each site, informed consent will be prepared according to the institutional requirements for informed consents. Consent will be collected by the investigator before patient inclusion in trial and before participating in any study procedure.

One copy of each consent form, signed by the participant and by the investigator, will be given to the patient and the original will be kept by the investigator.

##### c. Privacy of patients

All records will be kept confidential. The patient´s name will not be released at any time and data sets for each subject will be identified only by the patient enrollment number.

##### d. Amendments

Proposed amendments must be submitted to the appropriate Competent Authorities (CA) and Ethics Committee (EC) and approved before the change is implemented. These changes are usually presented in the form of an amendment. Amendments that are intended to eliminate an apparent immediate hazard to subjects may be implemented prior to receiving CA/EC approval. However, in this case, approval must be obtained as soon as possible after implementation.

##### e. Insurance

Patients will be covered by a clinical trial insurance. The patients will be insured as defined during their participation in the clinical trial by legal requirements. The investigator of the clinical trial will receive a copy of the insurance conditions of the ´patients insurance´. The sponsor is providing insurance in order to indemnify (legal and financial coverage) the investigator/center against claims arising from the study, except for claims that arise from malpractice and/or negligence. The compensation of the subject in the event of study-related injuries will comply with the applicable regulations. Details on the existing patients insurance are given in the patient information sheet.

Patients will be insured with a national insurance partner. The name and contact details for the insurer will be provided on the informed consent form.

##### f. Regulatory Requirements and GCP

The investigators at all sites are responsible for, and should warrant that the conduct of the study shall be compliant to ICH-GCP, local regulatory requirements with the EC-approved research protocol.

The investigators will ensure, that this study is conducted in full conformance with the principles of the “Declaration of Helsinki” (as amended at the 64th WMA General Assembly, Fortaleza, Brazil, October 2013) and with the national laws and regulations of the country in which the clinical trial is conducted.

g. Research Ethics

Patients who will participate on this clinical trial will receive the third SARS-CoV-2 vaccination in a controlled way and will be informed about their antibody results immediately. Knowledge gained from this study may be helpful to determine efficacy of additional SARS-CoV-2 vaccination. Additional risk equates to the risk of blood draw in general. Therefore pain, local irritation, small bruises, local damage of nerves but also dizziness and infections can occur. Assessed data will be stored from the principal investigator. Only authorized team members have access to the data. Processing of the data is only performed on password protected computers at the General Hospital Vienna.

#### Study Organization

##### h. Data Collection and Case Report From

For each subject enrolled, regardless of the study drug initiation, an CRF must be completed by the investigator or a designated sub-investigator. This also applies to those subjects, who fail to complete the study. If a subject withdraws from the study, the reason must be noted on the CRF. Case report forms are to be completed on an ongoing basis. Entries and corrections will only be performed by study site staff, who have been authorized by the investigator. Entry errors have to be corrected according the ICH-GCP Guidelines.

The entries will be checked by trained personnel (monitor) and any errors or inconsistencies will be checked and corrected immediately.

Data, collected at all visits, are entered into an Excel sheet. The CRFs constitute source documents established before study onset as detailed in the monitoring plan. The Maintenance of the study database will be performed by the Division of Rheumatology of the Medical University of Vienna.

The investigator shall maintain the records of drug disposition and the final CRF´s for a minimum of 15 years after the study closure.

##### i. Monitoring

The principal investigator will ensure, that the trial will be appropriately monitored by ensuring that all the rights of the subject are adequately protected, that the trial data are accurate, complete and verifiable from source documents and that the conduct of the trial is in compliance with the protocol and its subsequent amendments, with GCP and with applicable regulatory requirements. Monitoring will be performed by a contract research organization.

The sponsor investigator will ensure, that monitoring activities occur following a pre-defined monitoring plan.

The investigators will verify, that for all patients a written informed consent is obtained before each subject´s participation in the trial. The investigators will also ensure, that all patients enrolled will be eligible according to the in- and exclusion criteria as defined in the protocol.

The on-site monitors will be responsible for verifying, that the appropriate assurances and certification for training in the protection of human subjects are in place at the site prior to the initiation of the protocol. They will provide education to all site staff regarding the conduct of the study according to good clinical practices (GCP´s). The sponsor will be responsible for ensuring, that informed consent has been obtained for each patient. These records will be verified during the monitoring/auditing visits that will be performed by the designated monitor.

Monitoring visits are planned at the beginning, during and at the end of the study according to the monitoring plan.

##### j. Audit and inspections

All investigators agree to accept audits and inspections by the competent authorities during and after completion of the study. All data and documents may be subject to audits and regulatory inspection.

##### k. Relevant protocol deviations

All protocol deviations will be listed in the study report and assessed as to their influence on the quality of the study analysis. No deviations from the protocol and of any type will be made without complying with all IRB/EC established procedures in accordance with applicable regulations.

##### l. Provision and handling for study medication

Vaccine for the study will be provided by the general Vaccination Board of the County of Vienna. Patients will be vaccinated utilizing the Vienna General Hospital Vaccination infrastructure (“Impfstrasse”, 4Süd). Study vaccine will be stored as recommended by the manufacturer’s instructions.

##### m. Accountability for study medication

The investigator or his/her representative will verify, that study drug supplies are received intact and in the correct amounts. This will be documented with sign and date, as well as the arrival of drugs. All sheets will be kept in the site files as a record of what was received. Additionally, an investigational product accountability log will be documented, including, but not limited to, date received, lot number, kit number, date dispensed, subject number and the identification of the person dispensing the drug.

**9. STATISTICAL ANALYSIS**

**9.1 Sample size considerations**

According to the available number of patients at our Department, including estimates of non-responders to a standard protocol of mRNA vaccination, and expected participation rates, we will attempt to include 60 patients into this trial. Based on a Chi² test comparing 3^rd^ mRNA versus vector vaccine, this number of patients will allow to achieve at least 80% power as shown in the table:

| Expected response rate to 3^rd^ dose mRNA vaccine | Minimum detectable difference allowing for 80% power |
| --- | --- |
| 5% | 28% |
| 10% | 31% |
| 20% | 35% |
| 30% | 36% |

For differences smaller than the minimum detectable difference statistical significance will not be possible to claim.

**9.2 Relevant protocol deviations**

All protocol deviations will be listed in the study report. Major deviations regarding subjects' safety will lead to withdrawal.

**9.3 Endpoints analysis**

The primary endpoint (difference in SARS-CoV-2 antibody seroconversion rate by week 4 after vaccination boost at baseline between third mRNA SARS-CoV-2 (Biontech/Pfizer or Moderna) and vector SARS-CoV-2 vaccine (AstraZeneca)) will be analyzed using Chi-squared Test. For the seroconversion rates, 95% confidence intervals will be calculated. All subjects with available antibody responses fulfilling the eligibility criteria will be included in the analysis.

The secondary endpoints (see section 7.3.2) will be analyzed using descriptive statistics.

**9.4 Missing, unused and spurious data**

Only subjects for whom data are available will be included in the statistical analysis. Missing values will neither be replaced nor estimated.

**9.5 Interim analysis**

No interim analysis will be performed.

**9.6 Software program(s)**

Statistical analysis will be performed using R studios software or SPSS Statistics (Version 17.0 or higher).

#### 10. **DISCLOSURE OF DATA**

The investigator will publish the results of the study as an original scientific report in a peer reviewed scientific journal. In addition, the findings will be presented as oral and poster presentations at international scientific meetings. All obtained data and information will be regarded confidentially.

No data, results or any information of this study may be used for publication without prior agreement of the principal investigator, who is mentioned on the title page of this protocol. The authors of a publication, manuscript, article, abstract or oral presentation, are those, who contribute to the results of the study and/or the writing of the paper. The principal investigator will write the first draft of the paper and – if applicable – delivers the related presentations. He will also be the first author of the main efficacy manuscript. The correspondence address mentioned in each publication is the Sponsor-investigator´s address, as specified on the front-page. Additional co-authors will be determined based on their contributions during the course of the trial.

### SARS-CoV-2 VAC3 study protocol Version 2.0, May 18^th^ 2021

### Clinical Study Protocol

A Randomized, Parallel Group, Single-Blind, Phase 2 Study to Evaluate the immune response of two classes of SARS-CoV-2 Vaccines employed as Third Vaccination in Patients under current Rituximab Therapy and no humoral response after standard mRNA vaccination

**Daniela Sieghart^1^, Selma Tobudic^2^, Martina Durechova^1^, Daniel Mrak^1^, Michael Bonelli^1^, Daniel Aletaha^1^**

**1** Department of Internal Medicine III, Division of Rheumatology, Medical University of Vienna, Austria

**2** Department of Medicine I, Division of Infectious Diseases and Tropical Medicine, Medical University of Vienna, Austria

**Confidentiality Statement**

The information contained in this document, especially unpublished data, is the property of the sponsor of this study. It is therefore provided to you in confidence as an Investigator, potential Investigator, or consultant, for review by you, your staff, and an Independent Ethics Committee or Institutional Review Board. It is understood that this information will not be disclosed to others without written authorization from the Sponsor except to the extent necessary to obtain informed consent from those persons to whom the study drug may be administered.

### 3. SPONSOR, INVESTIGATOR, MONITOR AND SIGNATURES

**Sponsor/or representative (OEL) (AMG §§ 2a, 31, 32)**

**Univ.-Prof. Dr.med.univ.** **Alexandra Kautzky-Willer,**

Department of Internal medicine III, Medical University of Vienna, Austria

Signature (OEL)	Date

**Coordinating Investigator (AMG §§ 2a, 35, 36**)

**Univ.-Prof. Dr.med.univ.** **Daniel Aletaha,**

Department of Internal Medicine III, Division of Rheumatology, Medical University of Vienna, Austria

Signature	Date

#### 4. **Study summary**

**Background.** Treatment with rituximab (RTX), a monoclonal antibody targeting CD20, constitutes an important therapeutic strategy for patients with several inflammatory rheumatic diseases. Some recent reports have already highlighted the risk of serious consequences upon severe acute respiratory syndrome coronavirus 2 (SARS-CoV-2) infection in patients treated with RTX. Besides the risk of a more severe disease course during B cell depleting therapy, a major concern is the immunogenicity of vaccination during immunomodulatory therapies, especially with RTX. Indeed, RTX has been shown to impair humoral responses to various vaccines including SARS-CoV-2 vaccination. Studies from other vaccines have shown that an additional vaccination or a change of the vaccine can lead to development of a humoral immune response.

**Objective.** To investigate if a third vaccination with a vector based vaccine is superior to a further dose of the vaccine used for basis immunization in previous non responders receiving rituximab.

**Methods.** We will perform a prospective single blind randomized controlled study. A total of 60 patients under rituximab treatment will be enrolled in this clinical trial who received two vaccinations with an mRNA vaccine. Four study visits per patient will be planned. During the screening visit antibodies to the receptor-binding domain will be determined. All patients without detectable humoral immunity against SARS-Cov2 will be invited for a second boost vaccination within 4 weeks after the screening visit. During the baseline visit patients will be randomized to receive a third vaccination with either an mRNA-SARS-CoV-2 vaccine (Biontech/Pfizer or Moderna – according to the vaccine used for their first two vaccinations) or a vector SARS-CoV-2 (AstraZeneca) vaccination as a second boost. Additional study visits are scheduled at weeks 1 and 4 for assessment of humoral and cellular immunity, as well as clinical signs of adverse effects of disease activity reactivation. Patients without humoral response at week 4 will be offered participation in an extended follow-up for additional 8 weeks (i.e. at week 8 and 12 after baseline)

**Expected Results.** We expect at least 30% of patients receiving a third SARS-CoV-2 vaccination to develop humoral response after 4 weeks. The change in mode of action of the vaccine might be beneficial to induce response in patients with rituximab.

##### **3. BACKGROUND**

The current pandemic caused by severe acute respiratory syndrome coronavirus‐2 (SARS‐CoV‐2) has led to exponentially rising morbidity and mortality worldwide. https://coronavirus.jhu.edu/map.html. Apart from aggressive quarantine and hygiene control measures, the most effective way to inhibit SARS‐CoV‐2 spread is a population‐wide vaccination campaign. The SARS-CoV-2 pandemic has required rapid action and the development of vaccines in an unprecedented timeframe. Data from the preclinical development of vaccine candidates for SARS-CoV and MERS-CoV enabled the initial step of experimental vaccine design to be essentially omitted, saving a considerable amount of time. In many cases, production processes were adapted from existing vaccines or vaccine candidates, and in some instances, preclinical and toxicology data from related vaccines could be used. As a result, the first clinical trial of a vaccine candidate for SARS-CoV-2 began in March 2020. Trials were designed such that clinical phases are overlapping and trial starts are staggered, with initial phase I/II trials followed by rapid progression to phase III trials after an interim analysis of the phase I/II data.

More than 180 vaccine candidates, based on several different platforms, are currently in development against SARS-CoV-2.^1^ The World Health Organization (WHO) maintains a working document that includes most of the vaccines in development and is available at https://www.who.int/publications/m/item/draft-landscape-of-covid-19-candidate-vaccines.

RNA vaccines are a relatively recent development. The genetic information for the antigen is delivered instead of the antigen itself. The antigen is then expressed in the vaccinated individual cells. ^2^ Either mRNA (with modifications) or a self-replicating RNA can be used. Higher doses are required for mRNA than for self-replicating RNA, and the RNA is usually delivered via lipid nanoparticles (LNPs).^2-5^

Two mRNA vaccines, BNT162b2, developed by BioNTech/Fosun Pharma/Pfizer, and mRNA-1273 developed by Moderna/NIAID, are currently in phase III trials. ^6^^,^ ^7^

In December 2020, BNT162b2 was under evaluation for emergency use authorization (EUA) for widespread use by several medical regulators globally. On 21 December, the European Medicines Agency (EMA) has given official approval for the BNT162b2 vaccine developed by BioNTech and Pfizer.

On 18 December 2020, mRNA-1273 developed by Moderna/NIAID was issued an emergency use authorization by the United States Food and Drug Administration. On 06 January, the EMA has given official approval for the mRNA-1273, developed by Moderna/NIAID.

Preliminary results from Phase I–II clinical trials on BNT162b2, published in October 2020, indicated the potential for its efficacy and safety. ^8^ ^9^ Findings from studies conducted in the United States and Germany among healthy men and women showed that two 30-μg doses of BNT162b2 elicited high SARS-CoV-2 neutralizing antibody titers and robust antigen-specific CD8+ and Th1-type CD4+ T-cell response. ^10^ Besides, the reactogenicity profile of BNT162b2 represented mainly short-term local (i.e., injection site) and systemic responses. These findings supported the progression of BNT162b2 into the next phase. In phase III, a total of 43548 participants underwent randomization, of whom 43448 received injections: 21720 with BNT162b2 and 21728 with placebo. There were 8 cases of COVID-19 with onset at least 7 days after the second dose among participants assigned to receive BNT162b2 and 162 cases among those assigned to placebo. BNT162b2 was 95% effective in preventing COVID-19 (95% credible interval, 90.3 to 97.6). Similar vaccine efficacy (generally 90 to 100%) was observed across subgroups defined by age, sex, race, ethnicity, baseline body-mass index, and the presence of coexisting conditions.

In July 2020, preliminary results of the Phase I dose-escalation clinical trial of mRNA-1273 were published, showing dose-dependent induction of neutralizing antibodies against SARS-CoV-2 as early as 15 days post-injection. 6 Mild to moderate adverse reactions, such as fever, fatigue, headache, myalgia, and pain at the injection site, were observed in all dose groups but were common with increased dosage. The vaccine in low doses was deemed safe and effective to advance a Phase III clinical trial using two 100-μg doses administered 29 days apart.

In a Phase III trial of mRNA-1273 conducted in the United States between July and October, enrolled and assigned 30 000 volunteers to two groups — one group receiving two 100-μg doses of mRNA-1273 vaccine and the other receiving a placebo of 0.9% sodium chloride. Preliminary data from Phase III clinical trial, published in November 2020, indicating 94% efficacy in preventing COVID-19 infection. Side effects included flu-like symptoms, such as pain at the injection site, fatigue, muscle pain, and headache. The Moderna results were not final – as the trial is not scheduled to conclude until late-2022– and were not peer-reviewed or published in a medical journal.

The efficacy of ChAdOx1 nCoV-19 vaccine was reported from phase III trials in the United Kingdom and Brazil.^11^ The interpretation of the results of these trials is complicated by a dosing error (in which some participants unintentionally received a half-dose for their first of two doses), a small number of participants, and differences in efficacy in the two countries. The overall efficacy in preventing symptomatic infection more than 14 days after the second dose was 70.4% (95%CI: 54.8%-80.6%), with efficacy of 62.1% (95%CI: 41.0%-75.7%) in those who received standard doses, and 90.0% (95%CI 67.4% to 97.0%) in those who received a half-dose followed by a standard dose. Notably, the lower efficacy was based on results from the study in Brazil, and higher efficacy was reported for the study in the United Kingdom. Hospitalizations and severe COVID-19 occurred rarely but exclusively in the placebo arm of these trials. Due to varying intervals between the first and second doses, vaccine efficacy after a single standard dose from day 22 to day 90 was modeled and estimated to be 76% (95%CI: 59%-86%), with maintenance of antibody levels up to day 90. Furthermore, vaccine efficacy appeared to be 82.4% (95%CI: 62.7%-91.7%) when the interval between doses was more than 12 weeks compared to 54.9% (95%CI: 32.7%-69.7%) when the interval was less than 6 weeks. Similarly, geometric mean antibody levels were higher with a longer prime-boost interval in those age 18-55 years.^12^ A larger phase III trial using two standard doses 28 days apart with a majority of the participants in the US recently completed enrollment. Preliminary results of this trial showed vaccine efficacy of 76% (95%CI: 68-82%) at preventing symptomatic infection and 100% efficacy at preventing severe or critical disease and hospitalization. Vaccine efficacy was consistent across ethnicity and age. TheChAdOx1nCoV-19 vaccine has received EUA from the United Kingdom and the European Union. In the United Kingdom, a single-blind multi-center randomized phase II/III trial of the ChAdOx1 nCOV-19 vaccine asked participants to provide a weekly self-administered nose and throat swab starting one week after administration of the first vaccine (or placebo). This study revealed that among those who were infected, vaccinated persons had lower peak viral load and shorter duration of RT-PCR-positive results for SARS-CoV-2 compared to controls, suggesting that the vaccine is effective in reducing transmission.^13^

Patients with immunosuppression with COVID-19 may be at higher risk of hospitalization and ICU admission than matched comparators.^14^ An mRNA vaccine vaccination should be performed as primary immunization with two vaccine-doses 21 or 28 days apart. There are no data about the efficacy and safety of mRNA vaccine against SARS-CoV-2 in patients with a chronic immune-mediated disease such as rheumatoid arthritis, certain kinds of vasculitis, and lupus. However, after vaccination, immune responses might vary among immunosuppressed patients, dependent upon underlying clinical conditions and the level of immunosuppression. Moreover, it is not clear whether mRNA vaccination against SARS-CoV-2 in the immunosuppressed has a protective effect at all. Specifically, B-cell depleting therapies like CD20 antagonists rituximab have a high risk of vaccination failure.^15^ As for other vaccines, the efficacy of a COVID-19 vaccine may be reduced in patients treated with rituximab. For patients treated with rituximab, it is preferable to administer the vaccine at least 6 months after the last infusion^16^ to have the highest possible number of restored B-cells. The current treatment scheme for rituximab is 500mg or 1000mg intravenously every 6 months.

However, the vaccine is not contraindicated in this population and the treating physician should discuss with the patient the appropriate timing of vaccination taking into consideration the context of pandemic, the risk of poor COVID-19 disease prognosis, and a possibly reduced humoral response to vaccination under rituximab treatment.

**4. STUDY RATIONALE**

Patients who had received rituximab are at high risk of non-response to SARS-CoV-2 vaccination. It is unclear whether patients who did not develop humoral immunity after a standard protocol application with a mRNA vaccine would benefit from a second boost of mRNA SARS-CoV-2 or from a single additional shot of vector SARS-CoV-2 vaccine.

**5. STUDY OBJECTIVES**

The study aims to investigate the humoral and cellular immune responses after a second boost vaccination against SARS-CoV-2 in adult patients treated with rituximab (anti B cell therapy) who did not show response to the first two vaccinations with an mRNA vaccine.

**5.1. Primary Objective (Hypothesis)**

To assess the immunogenicity to a third vaccination mRNA-SARS-CoV-2 vaccine (Biontech/Pfizer or Moderna) compared to a vector SARS-CoV-2 (AstraZeneca) vaccination as a second boost in patients with rituximab by measuring quantitative antibody levels by enzyme-linked immunosorbent assay test (ELISA) and neutralization test (NT) or pseudo viral neutralization assay.

**Null and alternative hypotheses:**

H0: There is no statistical difference in the seroconversion rate between patients receiving a third mRNA vaccination and the patients receiving a second boost with AstraZeneca.

H1: There is statistical difference in the seroconversion rate between patients receiving a third mRNA vaccination and the patients receiving a second boost with AstraZeneca.

**Secondary Objectives**

- To compare cellular immunogenicity of the third mRNA SARS-Cov-2 vaccination will be compared to patients receiving AstraZeneca vaccination as second boost in immunosuppressed patients. T cell proliferation will be assessed, and T-cell cytokine expression will be measured using flow-cytometry following *in vitro* stimulation of peripheral blood mononuclear cells (PBMCs) with SARS-Cov-2 specific antigens.
- To assess safety of a second boost vaccination.
- To evaluate the influence of vaccination on underlying disease activity.

**6. STUDY DESIGN**

A prospective single blind randomized controlled study will be performed. A total of 60 patients with B-cell depleting therapy (rituximab) will be enrolled in this clinical trial. Four study visits per patient will be planned.

After inclusion of the patients (after receiving their written informed consent) serum samples for determination of antibody levels will be obtained at least 4 weeks after the second mRNA vaccination (screening visit). All patients without detectable humoral immunity against SARS-Cov2 will be invited for a second boost vaccination within 4 weeks after the screening visit. Additional study visits are scheduled at baseline (vaccination), and at weeks 1 and 4 for assessment of humoral and cellular immunity, as well as clinical signs of adverse effects of disease activity reactivation. Patients without humoral response at week 4 will be offered participation in an extended follow-up for additional 8 weeks (i.e. at week 8 and 12 after baseline)

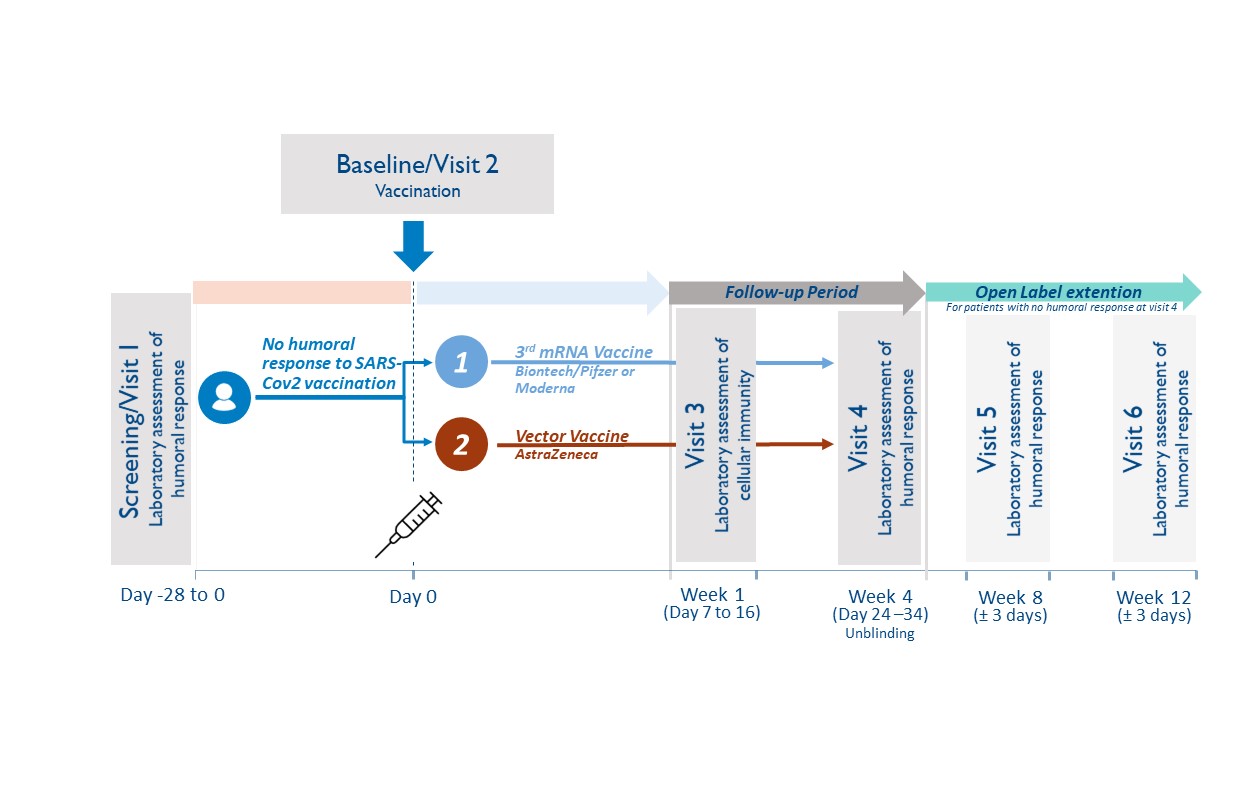

**6.1. Study population**

Adult Patient (≥ 18 years) under current rituximab therapy who did not develop humoral response against SARS-CoV-2 after their standard vaccination with Biontech/Pfizer or Moderna will be recruited at the Division of Rheumatology, Medical University of Vienna.

Study entry is defined as the date of signature of the study participant (subject) on the informed consent form. All subjects enrolled will be assigned a subject code, consisting of a consecutive subject number (2 digits).

**6.2. Inclusion criteria**

Male and female subjects will be eligible for participation in this study if they:

8. Are ≥18 years on the day of screening
9. Have a rheumatic condition and have been treated with a B-cell depleting therapy (rituximab) within the last 12 months
10. Received two doses of mRNA SARS-CoV-2 (Biontech/Pfizer or Moderna) vaccine according to recommendations in the label and/or national guidelines.
11. Did not develop humoral immunity 4 weeks after second mRNA vaccination to SARS-CoV-2 (analyzed during the study “Characterization of immune responsiveness after mRNA SARS-CoV-2 Vaccination in patients with immunodeficiency or immunosuppressive therapy”, EK-Nr. 1073/2021, EudraCT Nr. 2021-000291-11)
12. A maximum of 6 months after second vaccination
13. Have an understanding of the study, agree to its provisions, and give written informed consent before study entry
14. If female and capable of bearing children – have a negative urine pregnancy test result at study entry and agree to employ adequate birth control measures for the duration of the study

**6.3.	Exclusion criteria**

Subjects will be excluded from participation in this study if they:

7. Have shown humoral response to the SARS-CoV-2 vaccination
8. Had grade 3 adverse effects from the mRNA vaccination reported
9. Pregnancy and breast feeding
10. Signs of SARS-CoV-2 infection (including previous positive PCR testing)
11. Any other contraindication to any of the study compounds
12. Urgent need for next rituximab application

**6.4.	Study duration**

For the individual study participant, the active blinded study phase will be 4 weeks, with possibility to enroll in an open label extension for additional 8 weeks.

**6.5.	Randomization Procedure**

The web-based computerized randomization algorithm “Randomizer for Clinical Trials” by the Medical University of Vienna will be used for randomization. Patients will be randomized in a 1:1 ratio between the third dose mRNA SARS-CoV-2 boost (Biontech/Pfizer or Moderna, respective of their initial vaccination compound) and a single dose boost vector SARS-CoV-2 vaccine (AstraZeneca). Randomization will be stratified by presence or absence of B-cell depletion (determined at screening visit).

**6.6.1.	Blinding**

All patients will be blinded. Blinding of initial treatment allocation will be maintained throughout the 4 week study period. Blinding is ensured during the routine vaccination protocol at the General Hospital Vienna, where patients will see the pre-arranged dose aliquots in syringes without reference to the type of vaccine. To facilitate the process of vaccination of full vials, patients randomized to the same vaccine will be scheduled on the same day. Blinding will be mainly performed to prevent selective drop-outs due to knowledge of treatment allocation.

**6.6.2.	Unblinding Process**

The blinding can be lifted at the request of an investigator, when knowledge of the treatment is essential for appropriate patient management (e.g. after trial termination). In case of an emergency, the principal investigator has the sole responsibility for determining if unblinding of a patient´s treatment assignment is warranted.

If any serious adverse event arises during the study and unblinding appears necessary, the subinvestigator will notify the principal Investigator within two days in maximum. In the following the matter will be discussed and may lead to unblinding.

**6.7.1	Withdrawal and replacement of subjects**

Criteria for withdrawal:

Subjects may prematurely discontinue from the study at any time. The study's premature discontinuation is to be understood when the subject did not undergo complete the last visit (study visit 4) or all pivotal assessments during the study.

Subjects must be withdrawn under the following circumstances:

•	at their own request

•	if the investigator feels it would not be in the best interest of the subject to continue

•	if the subject violates conditions laid out in the consent form/information sheet or disregards instructions by the study personal

In all cases, the reason why subjects are withdrawn must be recorded in detail in the CRF and the subject's medical records. Should the study be discontinued prematurely, all study materials (complete, partially completed, and empty CRFs) will be retained.

Follow-up of patients withdrawn from the study:

In case of premature discontinuation after the start of vaccination, no further investigations concerning the study will be performed. Furthermore, participants may request that no more data will be recorded from the time point of withdrawal and that all biological samples collected in the course of the study will be destroyed.

**6.7.2	Premature termination of the study**

The sponsor has the right to close this study at any time. The IEC and the competent regulatory authority must be informed within 15 days of early termination.

The trial or single-dose steps will be terminated prematurely in the following cases:

- If the number of drop-outs is so high that proper completion of the trial cannot realistically be expected.
- Recruitment is not reaching the critical number

#### 7.8. **Adverse events and reporting**

##### j. Definition of adverse events

An adverse event is any untoward medical occurrence in a patient or clinical investigation subject administered a pharmaceutical product and which does not necessarily have a causal relationship with this treatment. An adverse event (AE) can therefore be any unfavourable and unintended sign (including an abnormal laboratory finding), symptom, or disease temporally associated with the use of a medicinal (investigational) product, whether or not related to the medicinal (investigational) product (see the ICH Guideline for Clinical Safety Data Management: Definitions and Standards for Expedited Reporting)

Adverse events include:

- Exacerbation of a pre-existing disease.
- Increase in frequency or intensity of a pre-existing episodic disease or medical condition.
- Disease or medical condition detected or diagnosed after study drug administration even though it may have been present prior to the start of the study.
- Continuous persistent disease or symptoms present at baseline that worsen following the start of the study.
- Lack of efficacy in the acute treatment of a life-threatening disease.
- Events considered by the Investigator to be related to study-mandated procedures.
- Abnormal assessments, e.g., ECG and physical examination findings, must be reported as AEs if they represent a clinically significant finding that was not present at baseline or worsened during the course of the study.
- Laboratory test abnormalities must be reported as AEs if they represent a clinically significant finding, symptomatic or not, which was not present at baseline or worsened during the course of the study or led to dose reduction, interruption or permanent discontinuation of study drug.

Adverse events do not include:

- Pre-planned interventions or occurrence of endpoints specified in the study protocol are not considered AE´s, if not defined otherwise (eg.as a result of overdose)
- Medical or surgical procedure, e.g., surgery, endoscopy, tooth extraction, transfusion. However, the event leading to the procedure is an AE. If this event is serious, the procedure must be described in the SAE narrative.
- Pre-existing disease or medical condition that does not worsen.
- Situations in which an adverse change did not occur, e.g., hospitalisations for cosmetic elective surgery or for social and/or convenience reasons.

Overdose of either study drug or concomitant medication without any signs or symptoms. However, overdose must be mentioned in the Study Drug Log.

##### k. Definition of serious adverse events (SAEs)

A Serious Adverse Event (SAE) is defined by the International Conference on Harmonization (ICH) guidelines as any AE fulfilling at least one of the following criteria:

- Fatal (including fetal death).
- Life-threatening – defined as an event in which the subject was, in the judgment of the investigator, at risk of death at the time of the event; it does not refer to an event that hypothetically might have caused death had it been more severe.
- Requiring subject's hospitalization or prolongation of existing hospitalization – inpatient hospitalization refers to any inpatient admission, regardless of length of stay.
- Resulting in persistent or significant disability or incapacity (i.e., a substantial disruption of a person’s ability to conduct normal life functions).
- Congenital anomaly or birth defect.
- Is medically significant or requires intervention to prevent at least one of the outcomes listed above.

In case of any SAE, the investigator has to use all means to ensure patient´s safety. Every SAE has to be reported as outlined in section 16.8.

##### l. Definition of suspected or unexpected serious adverse reactions (SUSARs)

SUSARs are all suspected adverse reactions related to the study drug that are both unexpected (not previously described in the SmPC or Investigator’s brochure) and serious.

##### m. Pregnancy

Maternal pregnancies must be reported to the principal investigator/sponsor. To ensure subject safety, each pregnancy must be reported to the PI/sponsor within 2 weeks of learning of its occurrence.

Pregnancies, should be followed up and reported to the PI/sponsor until the outcome of the pregnancy (including premature termination) and status of mother and child is known.

##### n. Severity of adverse events

The severity of clinical AEs is graded on a three-point scale: mild, moderate, severe, and reported on specific AE pages of the CRF.

If the severity of an AE worsens during study drug administration, only the worst intensity should be reported on the AE page. If the AE lessens in intensity, no change in the severity is required:

- *Mild* - Event may be noticeable to subject; does not influence daily activities; the AE resolves spontaneously or may require minimal therapeutic intervention;
- *Moderate* - Event may make subject uncomfortable; performance of daily activities may be influenced; intervention may be needed; the AE produces no sequelae.
- *Severe* - Event may cause noticeable discomfort; usually interferes with daily activities; subject may not be able to continue in the study; the AE produces sequelae, which require prolonged therapeutic intervention.

A mild, moderate or severe AE may or may not be serious.

##### o. Relationship of adverse events to study drug

For all AEs, the investigator will assess the causal relationship between the study drug and the AE using his/her clinical expertise and judgment according to the following algorithm that best fits the circumstances of the AE:

**Not related**

- May or may not follow a temporal sequence from administration of the study product
- Is biologically implausible and does not follow known response pattern to the suspect study drug (if response pattern is previously known).
- Can be explained by the known characteristics of the subject’s clinical state or other modes of therapy administered to the subject.

**Unlikely**

- There is a reasonable temporal relation between the AE and the intake of the study medication, but there is a plausible other explanation for the occurrence of the AE.

**Possibly**

- The AE has a reasonable temporal relationship with drug administration.
- The AE may equally be explained by the study subject’s clinically state, environmental or toxic factors, or concomitant therapy administered to the study subject.
- The relationship between study drug and AE may also be pharmacologically or clinically plausible.

**Probably**

- There is a reasonable temporal relation between the AE and the intake of the study medication, and plausible reasons point to a causal relation with the study medication.

**Related**

- Reasonable temporal relation between the AE and the intake of the study medication and
- There is no other explanation for the AE and
- Subsidence or disappearance of the AE on withdrawal of the study medication and
- Recurrence of the symptoms on restart at previous dose (only applies for re-institution of mediation).

**Not assessable**

The causal relationship between the study drug and the AE cannot be judged.

##### p. Adverse events reporting procedures

A special section is designated to adverse events in the case report form. For each subject, adverse events occurring after signing the informed consent must be recorded on the applicable adverse events page(s) in the case report form. Recording should be done in a concise manner using standard, acceptable medical terms. The adverse event recorded should not be a procedure or a clinical measurement (i.e., a laboratory value or vital sign) but should reflect the reason for the procedure or the diagnosis based on the abnormal measurement. If, in the investigator’s judgment, a clinically significant worsening from baseline is observed in laboratory or other parameters, physical exam finding, or vital sign, a corresponding clinical adverse event should be recorded on the adverse event page(s) of the CRF. If a specific medical diagnosis has been made that diagnosis should be recorded on the adverse event page(s) of the CRF.

SAEs and any newly identified pregnancy (maternal or paternal exposure), malignancy, overdose, opportunistic infection or case of active TB occurring after first administration of study agent in subjects participating in this clinical trial requires submission of a Safety Report Form to the Sponsor (only if other sites are reporting SAE) within 24 hours of notification or observation.

*The following details must thereby be entered:*

- Type of adverse event
- Start (date and time)
- End (date and time)
- Severity (mild, moderate, severe)
- Serious (no / yes)
- Unexpected (no / yes)
- Outcome (resolved, ongoing, ongoing – improved, ongoing – worsening)
- Relation to study drug (unrelated, possibly related, definitely related)

Adverse events are to be documented in the case report form in accordance with the above mentioned criteria.

##### q. Reporting procedures for SAEs

In the event of serious, the investigator has to use all supportive measures for best patient treatment. A serious adverse event must be reported if it occurs during a subject’s participation in the study (whether receiving study agent or not) or within six (6) months of receiving the last dose of study agent.

A Safety Report Form must be completed and faxed to the Sponsor (+43 (0)1 40400 43060) 24 hours of observation or notification of the event. The sponsor will be responsible for potential SUSAR assessment (see below) and reporting SAEs including potential SUSAR to the manufacturing company. A written report is also to be prepared and made available to the clinical investigator within five days.

The following details should at least be available:

- Patient initials and number
- Patient: date of birth, sex, ethical origin
- The suspected investigational medical product (IMP)
- The adverse event assessed as serious
- Short description of the event and outcome

Any serious adverse event that is ongoing when a subject completes his/her participation in the trial must be followed until any of the following occurs:

- The event resolves or stabilizes
- The event returns to baseline condition or value (if a baseline value is available)
- The event can be attributed to agent(s) other than the study agent, or to factors unrelated to study conduct

##### r. Reporting procedures for SUSARs

All SAEs will be evaluated regarding a possible classification as SUSAR by the sponsor, who will then perform all necessary reports to the manufacturing/distributing pharmaceutical company and forward the SUSAR to the CRO. The CRO will be responsible for reporting SUSARs to the regulatory authorities. In addition, due to their possible safety concern for the study participants, all SUSARs need to be reported to the Institutional Review Board / Independent Ethics Committee (IRB / IEC).

These reports are time critical and should be done within a maximum of 7 days (fatal or life threatening outcome) or 15 days (non fatal, not life threatening). The representative of the Sponsor Investigator shall inform all investigators concerned of relevant information about Serious Unexpected Suspected Adverse Reactions that could adversely affect the safety of subjects.

Such reports shall be made by the study management and the following details should be at least available:

- Patient inclusion number
- Patient: year of birth, sex, ethical origin
- Name of investigator and investigating site
- Period of administration
- The suspected investigational medical product (IMP)
- The adverse event assessed as serious and unexpected, and for which there is a reasonable suspected causal relationship to the IMP
- Concomitant disease and medication
- Short description of the event:
  - Description
  - Onset and if applicable, end
  - Therapeutic intervention
  - Causal relationship
  - Hospitalization of prolongation of hospitalization

8. **METHODOLOGY**

**7.1 Study design and blood samples**

A randomized controlled single-blind phase II trial will be performed. A total of 60 patients with current rituximab therapy who did not develop humoral immune response to SARS-CoV-2 after vaccination with mRNA SARS-CoV-2 vaccine (Biontech/Pfizer or Moderna). Four study visits per patient will be planned during the course of the trial: screening visit (-0-14 days), baseline, week 1, week 4. Patients will be unblinded at week 4. In addition, patients who did not show humoral response at week 4 will be invited for two additional study visits at week 8 and week 12 in an open label approach. Vaccination will take place at baseline visit.

Blood samples for the determination of SARS-CoV-2 antibodies will be obtained at screening visit, at week 1, week 4 and during the potential extension of the study at week 8 and week 12.

**7.2	Study procedures**

**7.2.1	General rules for trial procedures**

•	All study measures like blood sampling and measurements have to be documented with the date (dd:mm:yyyy).

•	In case several study procedures are scheduled simultaneously, there is no specific sequence that should be followed.

•	The dates of all procedures should be according to the protocol. The time margins mentioned in the study flow chart are admissible. If for any reason, a study procedure is not performed within scheduled margins, a protocol deviation should be noted, and the procedure should be performed as soon as possible or as adequate.

•	If it is necessary for organizational reasons, it is admissible to perform procedures scheduled for one visit at two different time points. Allowed time margins should thereby not be exceeded.

**7.2.2	Study visits**

**7.2.2.1 Screening visit (visit 1, -1-28 days, at least 4 weeks after the second immunization with mRNA SARS-CoV-2 vaccine)**

The investigator will inform the subject about the procedures, risks, and benefits of the study. Participants will be informed about future visits, which will be synchronized with the appointment for the third vaccination.

Fully informed, written consent must be obtained from each subject before any assessment is performed. The subject must be allowed sufficient time to consider his/her participation in the study.

The following assessments will be performed:

- Inclusion and exclusion criteria
- Demographic data, including sex, age, weight, and height
- Medical history and concomitant medication
- Assessment of adverse reactions after the mRNA SARS-CoV-2 vaccination
- Assessment of recent COVID-19 disease
- Blood will be drawn:
  - 16 ml of blood will be drawn for PBMC isolation and determination of cellular immunity
  - 8 ml of blood will be drawn for SARS-Cov-2 antibody level detection (prior vaccination).
  - 8 ml of blood will be drawn for determination of leukocyte subpopulations
  - 8 ml of blood will be drawn for biobanking of serum
  - 20 ml of blood will be drawn for routine laboratory testing: blood count, chemistry, coagulation factors
- Pregnancy test in women with childbearing potential
- Randomization to one of the three vaccines: mRNA SARS-CoV-2 (Biontech/Pfizer or Moderna) or vector SARS-CoV-2 vaccination (AstraZeneca)
- SARS-CoV-2 vaccine appointment

**7.2.2.2. Baseline visit (visit 2, week 0, max. 28 days after visit 1)**

The following activities will be performed:

- The investigator will ask all subjects about any adverse experiences occurring since Visit 1 (screening). All adverse experiences will be documented in the CRF.
- Patients will receive a patients diary, fever thermometer and spacer
- Application of mRNA SARS-CoV-2 (Biontech/Pfizer or Moderna) or vector SARS-CoV-2 vaccine (AstraZeneca)
- Patients health status will be observed for additional 30 minutes by a clinician and a nurse in case of any acute reactions to the vaccination

**7.2.2.3. Visit 3 (1 week after the third SARS-CoV-2 vaccination)**

The following activities will be performed:

- The investigator will ask all subjects about any adverse experiences occurring since Visit 2. All adverse experiences will be documented in the CRF.
- Patient´s diaries will be discussed
- Blood will be drawn:
  - 16 ml of blood will be drawn for PBMC isolation and determination of cellular immunity
  - 8 ml of blood will be drawn for SARS-Cov-2 antibody level detection
  - 6 ml of blood will be drawn for determination of leukocyte subpopulations
  - 8 ml of blood will be drawn for biobanking of serum
  - 20 ml of blood will be drawn for routine laboratory testing: blood account, chemistry as well as thrombocytes and coagulation factors
  - 8 ml blood will be drawn for detection of anti-PDE4D antibody

**7.2.2. 4. Visit 4 (4 weeks after the third SARS-CoV-2 vaccination)**

The following activities will be performed:

•	The investigator will ask all subjects about any adverse experiences occurring since Visit 3. All adverse experiences will be documented in the CRF.

- Patient´s diary will be returned to investigator

•	Blood draw:

- 8 ml of blood will be drawn for SARS-Cov-2 antibody level detection
- 6 ml of blood will be drawn for determination of leukocyte subpopulations
- 8 ml of blood will be drawn for biobanking of serum
- 20 ml of blood will be drawn for routine laboratory testing: blood account, chemistry, coagulation factors
- 8 ml blood will be drawn for detection of anti-PDE4D antibody

**7.2.2.5. Visits 5 (week 8) and 6 (week 12) (only for patients enrolling in open label extension)**

Fully informed, written consent to the extension period must be obtained from each subject before any assessment is performed. The subject must be allowed sufficient time to consider his/her participation in the open label extension.

The following activities will be performed:

•	The investigator will ask all subjects about any adverse experiences occurring since Visit 3. All adverse experiences will be documented in the CRF.

- Blood draw:
  - 8 ml of blood will be drawn for SARS-CoV-2 antibody level detection
  - 8 ml of blood will be drawn for biobanking of serum

**7.2.3	Determination of the humoral and cellular immunity**

**7.2.3.1 Determination of serum antibodies against vaccination antigens**

Analysis will be performed at the Department of Laboratory Medicine, Medical University of Vienna. All procedures will be carried out according to standard operating procedures in an ISO 9001:2015 certified environment (ref: 10.1089/bio.2018.0032).

Previous SARS-CoV-2 infection will be detected using nucleocapsid-based chemiluminescence assays (e.g., Roche SARS-CoV-2 NC total antibody ECLIA, Abbott SARS-CoV-2 NC IgG CMIA). Vaccination response will be assessed by spike-protein-based assays (e.g., Technozym RBD ELISA, Siemens RBD immunoassay, Roche SARS-CoV-2 RBD ECLIA).

All analyses will be carried out at the Department of Laboratory Medicine, Medical University of Vienna.

Neutralization assays will be performed as described earlier.^13^

**7.2.3.2 Determination of cellular immune response**

To investigate cellular immunity following SARS-CoV-2 vaccination Peripheral blood mononuclear cells (PBMCs) will be isolated from heparinized venous blood by density gradient centrifugation at 400 *g* of heparinized blood over LSM 1077 Lymphocyte Separation Medium, cryopreserved and stored in liquid nitrogen for later use.

**7.3	Study endpoints**

**7.3.1	Primary study endpoint**

Difference in SARS-CoV-2 antibody seroconversion rate by week 4 after vaccination boost at baseline between 3^rd^ mRNA SARS-CoV-2 (Biontech/Pfizer or Moderna) and vector SARS-CoV-2 vaccine (AstraZeneca).

**7.3.2	Secondary study endpoints**

The secondary endpoints of this study are:

- Overall SARS-Cov-2 antibody seroconversion rate by week 4 after vaccination boost at baseline
- Difference in overall SARS-Cov-2 antibody seroconversion rate by week 4 after vaccination boost at baseline between patient with and without B-cell repopulation.
- Antibody concentrations of SARS-CoV-2 ELISA 4 weeks after vaccination boost at baseline.
- Effect of immunosuppressive comedication on SARS-Cov-2 antibody seroconversion rate by week 4 after vaccination boost at baseline
- Evaluation of cellular immunity before and one week after the vaccination.
- Safety of vaccination boost.

#### 9. **ETHICAL CONSIDERATIONS**

##### a.  Ethical Review

All relevant documents must be reviewed and approved by the Ethics Committee and additionally submitted to the relevant authorities in accordance with the guidance of submission and conduct of clinical trials.

The clinical trial shall be performed in full compliance with the legal regulations according to the Drug Law (AMG - Arzneimittelgesetz) of the Republic of Austria.

An application must also be submitted to the Austrian Competent Authorities (Bundesamt für Sicherheit im Gesundheitswesen (BASG) represented by the Agency for Health and Food Safety (AGES PharmMed), and registered to the European Clinical Trial Database (EudraCT) using the required forms. The timelines for (silent) approval set by national law must be followed before starting the study.

##### b. Consent Procedures

After a detailed information about the study procedures and study medication, as well as the potentially related risks and benefits, the written informed consent will be obtained from each participant by the principal investigator or a designee. At each site, informed consent will be prepared according to the institutional requirements for informed consents. Consent will be collected by the investigator before patient inclusion in trial and before participating in any study procedure.

One copy of each consent form, signed by the participant and by the investigator, will be given to the patient and the original will be kept by the investigator.

##### c. Privacy of patients

All records will be kept confidential. The patient´s name will not be released at any time and data sets for each subject will be identified only by the patient enrollment number.

##### d. Amendments

Proposed amendments must be submitted to the appropriate Competent Authorities (CA) and Ethics Committee (EC) and approved before the change is implemented. These changes are usually presented in the form of an amendment. Amendments that are intended to eliminate an apparent immediate hazard to subjects may be implemented prior to receiving CA/EC approval. However, in this case, approval must be obtained as soon as possible after implementation.

##### e. Insurance

Patients will be covered by a clinical trial insurance. The patients will be insured as defined during their participation in the clinical trial by legal requirements. The investigator of the clinical trial will receive a copy of the insurance conditions of the ´patients insurance´. The sponsor is providing insurance in order to indemnify (legal and financial coverage) the investigator/center against claims arising from the study, except for claims that arise from malpractice and/or negligence. The compensation of the subject in the event of study-related injuries will comply with the applicable regulations. Details on the existing patients insurance are given in the patient information sheet.

Patients will be insured with a national insurance partner. The name and contact details for the insurer will be provided on the informed consent form.

##### f. Regulatory Requirements and GCP

The investigators at all sites are responsible for, and should warrant that the conduct of the study shall be compliant to ICH-GCP, local regulatory requirements with the EC-approved research protocol.

The investigators will ensure, that this study is conducted in full conformance with the principles of the “Declaration of Helsinki” (as amended at the 64th WMA General Assembly, Fortaleza, Brazil, October 2013) and with the national laws and regulations of the country in which the clinical trial is conducted.

g. Research Ethics

Patients who will participate on this clinical trial will receive the third SARS-CoV-2 vaccination in a controlled way and will be informed about their antibody results immediately. Knowledge gained from this study may be helpful to determine efficacy of additional SARS-CoV-2 vaccination. Additional risk equates to the risk of blood draw in general. Therefore pain, local irritation, small bruises, local damage of nerves but also dizziness and infections can occur. Assessed data will be stored from the principal investigator. Only authorized team members have access to the data. Processing of the data is only performed on password protected computers at the General Hospital Vienna.

#### Study Organization

##### h. Data Collection and Case Report From

For each subject enrolled, regardless of the study drug initiation, an CRF must be completed by the investigator or a designated sub-investigator. This also applies to those subjects, who fail to complete the study. If a subject withdraws from the study, the reason must be noted on the CRF. Case report forms are to be completed on an ongoing basis. Entries and corrections will only be performed by study site staff, who have been authorized by the investigator. Entry errors have to be corrected according the ICH-GCP Guidelines.

The entries will be checked by trained personnel (monitor) and any errors or inconsistencies will be checked and corrected immediately.

Data, collected at all visits, are entered into an Excel sheet. The CRFs constitute source documents established before study onset as detailed in the monitoring plan. The Maintenance of the study database will be performed by the Division of Rheumatology of the Medical University of Vienna.

The investigator shall maintain the records of drug disposition and the final CRF´s for a minimum of 15 years after the study closure.

##### i. Monitoring

The principal investigator will ensure, that the trial will be appropriately monitored by ensuring that all the rights of the subject are adequately protected, that the trial data are accurate, complete and verifiable from source documents and that the conduct of the trial is in compliance with the protocol and its subsequent amendments, with GCP and with applicable regulatory requirements. Monitoring will be performed by a contract research organization:

CW-Research & Management GmbH

A-1130 Wien, Auhofstraße 84/3/39

Web: [www.cw-rm.com](https://www.cw-rm.com)

The sponsor investigator will ensure, that monitoring activities occur following a pre-defined monitoring plan.

The investigators will verify, that for all patients a written informed consent is obtained before each subject´s participation in the trial. The investigators will also ensure, that all patients enrolled will be eligible according to the in- and exclusion criteria as defined in the protocol.

The on-site monitors will be responsible for verifying, that the appropriate assurances and certification for training in the protection of human subjects are in place at the site prior to the initiation of the protocol. They will provide education to all site staff regarding the conduct of the study according to good clinical practices (GCP´s). The sponsor will be responsible for ensuring, that informed consent has been obtained for each patient. These records will be verified during the monitoring/auditing visits that will be performed by the designated monitor.

Monitoring visits are planned at the beginning, during and at the end of the study according to the monitoring plan.

##### j. Audit and inspections

All investigators agree to accept audits and inspections by the competent authorities during and after completion of the study. All data and documents may be subject to audits and regulatory inspection.

##### k. Relevant protocol deviations

All protocol deviations will be listed in the study report and assessed as to their influence on the quality of the study analysis. No deviations from the protocol and of any type will be made without complying with all IRB/EC established procedures in accordance with applicable regulations.

##### l. Provision and handling for study medication

Vaccine for the study will be provided by the general Vaccination Board of the County of Vienna. Patients will be vaccinated utilizing the Vienna General Hospital Vaccination infrastructure (“Impfstrasse”, 4Süd). Study vaccine will be stored as recommended by the manufacturer’s instructions.

##### m. Accountability for study medication

The investigator or his/her representative will verify, that study drug supplies are received intact and in the correct amounts. This will be documented with sign and date, as well as the arrival of drugs. All sheets will be kept in the site files as a record of what was received. Additionally, an investigational product accountability log will be documented, including, but not limited to, date received, lot number, kit number, date dispensed, subject number and the identification of the person dispensing the drug.

**9. STATISTICAL ANALYSIS**

**9.1 Sample size considerations**

According to the available number of patients at our Department, including estimates of non-responders to a standard protocol of mRNA vaccination, and expected participation rates, we will attempt to include 60 patients into this trial. Based on a Chi² test comparing 3^rd^ mRNA versus vector vaccine, this number of patients will allow to achieve at least 80% power at a minimal detectable difference of 28% (5% of responders in the mRNA vaccine group versus 33% of responders in the vector vaccine group).

For differences smaller than the minimum detectable difference statistical significance will not be possible to claim.

**9.2 Relevant protocol deviations**

All protocol deviations will be listed in the study report. Major deviations regarding subjects' safety will lead to withdrawal.

**9.3 Endpoints analysis**

The primary endpoint (difference in SARS-CoV-2 antibody seroconversion rate by week 4 after vaccination boost at baseline between third mRNA SARS-CoV-2 (Biontech/Pfizer or Moderna) and vector SARS-CoV-2 vaccine (AstraZeneca)) will be analyzed using Chi-squared Test. For the seroconversion rates, 95% confidence intervals will be calculated. All subjects with available antibody responses fulfilling the eligibility criteria will be included in the analysis.

The secondary endpoints (see section 7.3.2) will be analyzed using descriptive statistics.

**9.4 Missing, unused and spurious data**

Only subjects for whom data are available will be included in the statistical analysis. Missing values will neither be replaced nor estimated.

**9.5 Interim analysis**

No interim analysis will be performed.

**9.6 Software program(s)**

Statistical analysis will be performed using R studios software or SPSS Statistics (Version 17.0 or higher).

#### 11. **DISCLOSURE OF DATA**

The investigator will publish the results of the study as an original scientific report in a peer reviewed scientific journal. In addition, the findings will be presented as oral and poster presentations at international scientific meetings. All obtained data and information will be regarded confidentially.

No data, results or any information of this study may be used for publication without prior agreement of the principal investigator, who is mentioned on the title page of this protocol. The authors of a publication, manuscript, article, abstract or oral presentation, are those, who contribute to the results of the study and/or the writing of the paper. The principal investigator will write the first draft of the paper and – if applicable – delivers the related presentations. He will also be the first author of the main efficacy manuscript. The correspondence address mentioned in each publication is the Sponsor-investigator´s address, as specified on the front-page. Additional co-authors will be determined based on their contributions during the course of the trial.

### SARS-CoV-2 VAC3 study protocol Version 3.0, June 1^st^ 2021

### Clinical Study Protocol

A Randomized, Parallel Group, Single-Blind, Phase 2 Study to Evaluate the immune response of two classes of SARS-CoV-2 Vaccines employed as Third Vaccination in Patients under current Rituximab Therapy and no humoral response after standard mRNA vaccination

**Daniela Sieghart^1^, Selma Tobudic^2^, Martina Durechova^1^, Daffodil Dioso^1^, Michael Zauner^1^, Daniel Mrak^1^, Peter Mandl^1^, Helga Lechner-Radner^1^, Michael Bonelli^1^, Barbara Kornek^3^, Daniel Aletaha^1^**

**1** Department of Internal Medicine III, Division of Rheumatology, Medical University of Vienna, Austria

**2** Department of Medicine I, Division of Infectious Diseases and Tropical Medicine, Medical University of Vienna, Austria

**3** Department of Neurology, Medical University of Vienna, Austria

**Confidentiality Statement**

The information contained in this document, especially unpublished data, is the property of the sponsor of this study. It is therefore provided to you in confidence as an Investigator, potential Investigator, or consultant, for review by you, your staff, and an Independent Ethics Committee or Institutional Review Board. It is understood that this information will not be disclosed to others without written authorization from the Sponsor except to the extent necessary to obtain informed consent from those persons to whom the study drug may be administered.

### 5. SPONSOR, INVESTIGATOR, MONITOR AND SIGNATURES

**Sponsor/or representative (OEL) (AMG §§ 2a, 31, 32)**

**Univ.-Prof. Dr.med.univ.** **Alexandra Kautzky-Willer,**

Department of Internal medicine III, Medical University of Vienna, Austria

Signature (OEL)	Date

**Coordinating Investigator (AMG §§ 2a, 35, 36**)

**Univ.-Prof. Dr.med.univ.** **Daniel Aletaha,**

Department of Internal Medicine III, Division of Rheumatology, Medical University of Vienna, Austria

Signature	Date

#### 6. **Study summary**

**Background.** Treatment with rituximab (RTX), a monoclonal antibody targeting CD20, constitutes an important therapeutic strategy for patients with several chronic inflammatory conditions such as rheumatoid arthritis, systemic lupus erythematosus, mixed connective tissue diseases and neurological disorders like multiple sclerosis and other demyelinating diseases of the central nervous system, immune-mediated muscular disorders or immune-mediated neuropathies. Some recent reports have already highlighted the risk of serious consequences upon severe acute respiratory syndrome coronavirus 2 (SARS-CoV-2) infection in patients treated with RTX. Besides the risk of a more severe disease course during B cell depleting therapy, a major concern is the immunogenicity of vaccination during immunomodulatory therapies, especially with RTX. Indeed, RTX has been shown to impair humoral responses to various vaccines including SARS-CoV-2 vaccination. Studies from other vaccines have shown that an additional vaccination or a change of the vaccine can lead to development of a humoral immune response.

**Objective.** To investigate if a third vaccination with a vector based vaccine is superior to a further dose of the vaccine used for basis immunization in previous non responders receiving rituximab.

**Methods.** We will perform a prospective single blind randomized controlled study. A total of 60 patients under rituximab treatment will be enrolled in this clinical trial who received two vaccinations with an mRNA vaccine. Four study visits per patient will be planned. During the screening visit antibodies to the receptor-binding domain will be determined. All patients without detectable humoral immunity against SARS-Cov2 will be invited for a second boost vaccination within 4 weeks after the screening visit. During the baseline visit patients will be randomized to receive a third vaccination with either an mRNA-SARS-CoV-2 vaccine (Biontech/Pfizer or Moderna – according to the vaccine used for their first two vaccinations) or a vector SARS-CoV-2 (AstraZeneca) vaccination as a second boost. Additional study visits are scheduled at weeks 1 and 4 for assessment of humoral and cellular immunity, as well as clinical signs of adverse effects of disease activity reactivation. Patients without humoral response at week 4 will be offered participation in an extended follow-up for additional 8 weeks (i.e. at week 8 and 12 after baseline)

**Expected Results.** We expect at least 30% of patients receiving a third SARS-CoV-2 vaccination to develop humoral response after 4 weeks. The change in mode of action of the vaccine might be beneficial to induce response in patients with rituximab.

##### **3. BACKGROUND**

The current pandemic caused by severe acute respiratory syndrome coronavirus‐2 (SARS‐CoV‐2) has led to exponentially rising morbidity and mortality worldwide. https://coronavirus.jhu.edu/map.html. Apart from aggressive quarantine and hygiene control measures, the most effective way to inhibit SARS‐CoV‐2 spread is a population‐wide vaccination campaign. The SARS-CoV-2 pandemic has required rapid action and the development of vaccines in an unprecedented timeframe. Data from the preclinical development of vaccine candidates for SARS-CoV and MERS-CoV enabled the initial step of experimental vaccine design to be essentially omitted, saving a considerable amount of time. In many cases, production processes were adapted from existing vaccines or vaccine candidates, and in some instances, preclinical and toxicology data from related vaccines could be used. As a result, the first clinical trial of a vaccine candidate for SARS-CoV-2 began in March 2020. Trials were designed such that clinical phases are overlapping and trial starts are staggered, with initial phase I/II trials followed by rapid progression to phase III trials after an interim analysis of the phase I/II data.

More than 180 vaccine candidates, based on several different platforms, are currently in development against SARS-CoV-2.^1^ The World Health Organization (WHO) maintains a working document that includes most of the vaccines in development and is available at https://www.who.int/publications/m/item/draft-landscape-of-covid-19-candidate-vaccines.

RNA vaccines are a relatively recent development. The genetic information for the antigen is delivered instead of the antigen itself. The antigen is then expressed in the vaccinated individual cells. ^2^ Either mRNA (with modifications) or a self-replicating RNA can be used. Higher doses are required for mRNA than for self-replicating RNA, and the RNA is usually delivered via lipid nanoparticles (LNPs).^2-5^

Two mRNA vaccines, BNT162b2, developed by BioNTech/Fosun Pharma/Pfizer, and mRNA-1273 developed by Moderna/NIAID, are currently in phase III trials. ^6^^,^ ^7^

In December 2020, BNT162b2 was under evaluation for emergency use authorization (EUA) for widespread use by several medical regulators globally. On 21 December, the European Medicines Agency (EMA) has given official approval for the BNT162b2 vaccine developed by BioNTech and Pfizer.

On 18 December 2020, mRNA-1273 developed by Moderna/NIAID was issued an emergency use authorization by the United States Food and Drug Administration. On 06 January, the EMA has given official approval for the mRNA-1273, developed by Moderna/NIAID.

Preliminary results from Phase I–II clinical trials on BNT162b2, published in October 2020, indicated the potential for its efficacy and safety. ^8^ ^9^ Findings from studies conducted in the United States and Germany among healthy men and women showed that two 30-μg doses of BNT162b2 elicited high SARS-CoV-2 neutralizing antibody titers and robust antigen-specific CD8+ and Th1-type CD4+ T-cell response. ^10^ Besides, the reactogenicity profile of BNT162b2 represented mainly short-term local (i.e., injection site) and systemic responses. These findings supported the progression of BNT162b2 into the next phase. In phase III, a total of 43548 participants underwent randomization, of whom 43448 received injections: 21720 with BNT162b2 and 21728 with placebo. There were 8 cases of COVID-19 with onset at least 7 days after the second dose among participants assigned to receive BNT162b2 and 162 cases among those assigned to placebo. BNT162b2 was 95% effective in preventing COVID-19 (95% credible interval, 90.3 to 97.6). Similar vaccine efficacy (generally 90 to 100%) was observed across subgroups defined by age, sex, race, ethnicity, baseline body-mass index, and the presence of coexisting conditions.

In July 2020, preliminary results of the Phase I dose-escalation clinical trial of mRNA-1273 were published, showing dose-dependent induction of neutralizing antibodies against SARS-CoV-2 as early as 15 days post-injection. 6 Mild to moderate adverse reactions, such as fever, fatigue, headache, myalgia, and pain at the injection site, were observed in all dose groups but were common with increased dosage. The vaccine in low doses was deemed safe and effective to advance a Phase III clinical trial using two 100-μg doses administered 29 days apart.

In a Phase III trial of mRNA-1273 conducted in the United States between July and October, enrolled and assigned 30 000 volunteers to two groups — one group receiving two 100-μg doses of mRNA-1273 vaccine and the other receiving a placebo of 0.9% sodium chloride. Preliminary data from Phase III clinical trial, published in November 2020, indicating 94% efficacy in preventing COVID-19 infection. Side effects included flu-like symptoms, such as pain at the injection site, fatigue, muscle pain, and headache. The Moderna results were not final – as the trial is not scheduled to conclude until late-2022– and were not peer-reviewed or published in a medical journal.

The efficacy of ChAdOx1 nCoV-19 vaccine was reported from phase III trials in the United Kingdom and Brazil.^11^ The interpretation of the results of these trials is complicated by a dosing error (in which some participants unintentionally received a half-dose for their first of two doses), a small number of participants, and differences in efficacy in the two countries. The overall efficacy in preventing symptomatic infection more than 14 days after the second dose was 70.4% (95%CI: 54.8%-80.6%), with efficacy of 62.1% (95%CI: 41.0%-75.7%) in those who received standard doses, and 90.0% (95%CI 67.4% to 97.0%) in those who received a half-dose followed by a standard dose. Notably, the lower efficacy was based on results from the study in Brazil, and higher efficacy was reported for the study in the United Kingdom. Hospitalizations and severe COVID-19 occurred rarely but exclusively in the placebo arm of these trials. Due to varying intervals between the first and second doses, vaccine efficacy after a single standard dose from day 22 to day 90 was modeled and estimated to be 76% (95%CI: 59%-86%), with maintenance of antibody levels up to day 90. Furthermore, vaccine efficacy appeared to be 82.4% (95%CI: 62.7%-91.7%) when the interval between doses was more than 12 weeks compared to 54.9% (95%CI: 32.7%-69.7%) when the interval was less than 6 weeks. Similarly, geometric mean antibody levels were higher with a longer prime-boost interval in those age 18-55 years.^12^ A larger phase III trial using two standard doses 28 days apart with a majority of the participants in the US recently completed enrollment. Preliminary results of this trial showed vaccine efficacy of 76% (95%CI: 68-82%) at preventing symptomatic infection and 100% efficacy at preventing severe or critical disease and hospitalization. Vaccine efficacy was consistent across ethnicity and age. TheChAdOx1nCoV-19 vaccine has received EUA from the United Kingdom and the European Union. In the United Kingdom, a single-blind multi-center randomized phase II/III trial of the ChAdOx1 nCOV-19 vaccine asked participants to provide a weekly self-administered nose and throat swab starting one week after administration of the first vaccine (or placebo). This study revealed that among those who were infected, vaccinated persons had lower peak viral load and shorter duration of RT-PCR-positive results for SARS-CoV-2 compared to controls, suggesting that the vaccine is effective in reducing transmission.^13^

Patients with immunosuppression with COVID-19 may be at higher risk of hospitalization and ICU admission than matched comparators.^14^ An mRNA vaccine vaccination should be performed as primary immunization with two vaccine-doses 21 or 28 days apart. There are no data about the efficacy and safety of mRNA vaccine against SARS-CoV-2 in patients with a chronic immune-mediated disease such as rheumatoid arthritis, certain kinds of vasculitis, lupus, multiple sclerosis or immune-mediated neurological conditions. However, after vaccination, immune responses might vary among immunosuppressed patients, dependent upon underlying clinical conditions and the level of immunosuppression. Moreover, it is not clear whether mRNA vaccination against SARS-CoV-2 in the immunosuppressed has a protective effect at all. Specifically, B-cell depleting therapies like CD20 antagonists rituximab have a high risk of vaccination failure.^15^ As for other vaccines, the efficacy of a COVID-19 vaccine may be reduced in patients treated with rituximab. For patients treated with rituximab, it is preferable to administer the vaccine at least 6 months after the last infusion^16^ to have the highest possible number of restored B-cells. The current treatment scheme for rituximab is 500mg or 1000mg intravenously every 6 months.

However, the vaccine is not contraindicated in this population and the treating physician should discuss with the patient the appropriate timing of vaccination taking into consideration the context of pandemic, the risk of poor COVID-19 disease prognosis, and a possibly reduced humoral response to vaccination under rituximab treatment.

**4. STUDY RATIONALE**

Patients who had received rituximab are at high risk of non-response to SARS-CoV-2 vaccination. It is unclear whether patients who did not develop humoral immunity after a standard protocol application with a mRNA vaccine would benefit from a second boost of mRNA SARS-CoV-2 or from a single additional shot of vector SARS-CoV-2 vaccine.

**5. STUDY OBJECTIVES**

The study aims to investigate the humoral and cellular immune responses after a second boost vaccination against SARS-CoV-2 in adult patients treated with rituximab (anti B cell therapy) who did not show response to the first two vaccinations with an mRNA vaccine.

**5.1. Primary Objective (Hypothesis)**

To assess the immunogenicity to a third vaccination mRNA-SARS-CoV-2 vaccine (Biontech/Pfizer or Moderna) compared to a vector SARS-CoV-2 (AstraZeneca) vaccination as a second boost in patients with rituximab by measuring quantitative antibody levels by enzyme-linked immunosorbent assay test (ELISA) and neutralization test (NT) or pseudo viral neutralization assay.

**Null and alternative hypotheses:**

H0: There is no statistical difference in the seroconversion rate between patients receiving a third mRNA vaccination and the patients receiving a second boost with AstraZeneca.

H1: There is statistical difference in the seroconversion rate between patients receiving a third mRNA vaccination and the patients receiving a second boost with AstraZeneca.

**Secondary Objectives**

- To compare cellular immunogenicity of the third mRNA SARS-Cov-2 vaccination will be compared to patients receiving AstraZeneca vaccination as second boost in immunosuppressed patients. T cell proliferation will be assessed, and T-cell cytokine expression will be measured using flow-cytometry following *in vitro* stimulation of peripheral blood mononuclear cells (PBMCs) with SARS-Cov-2 specific antigens.
- To assess safety of a second boost vaccination.
- To evaluate the influence of vaccination on underlying disease activity.

**6. STUDY DESIGN**

A prospective single blind randomized controlled study will be performed. A total of 60 patients with B-cell depleting therapy (rituximab) will be enrolled in this clinical trial. Four study visits per patient will be planned.

After inclusion of the patients (after receiving their written informed consent) serum samples for determination of antibody levels will be obtained at least 4 weeks after the second mRNA vaccination (screening visit). All patients without detectable humoral immunity against SARS-Cov2 will be invited for a second boost vaccination within 4 weeks after the screening visit. Additional study visits are scheduled at baseline (vaccination), and at weeks 1 and 4 for assessment of humoral and cellular immunity, as well as clinical signs of adverse effects of disease activity reactivation. Patients without humoral response at week 4 will be offered participation in an extended follow-up for additional 8 weeks (i.e. at week 8 and 12 after baseline)

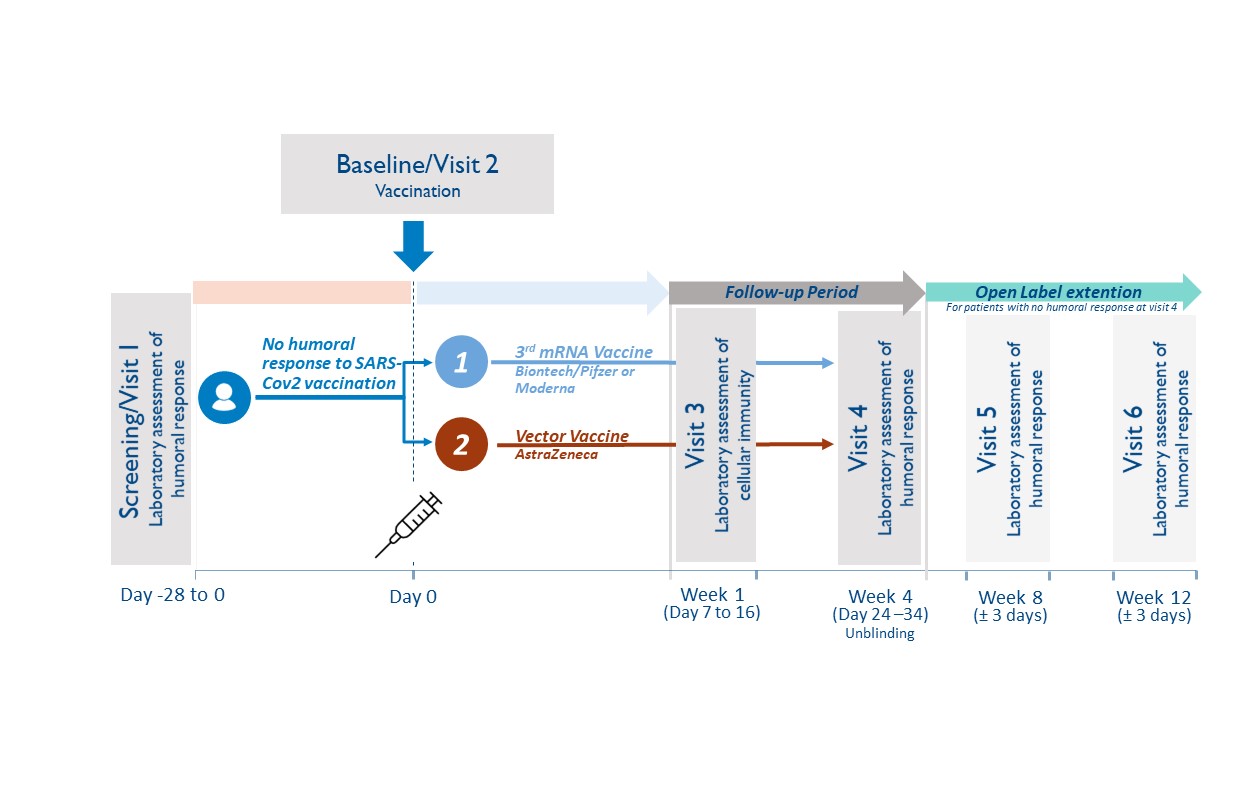

**6.1. Study population**

Adult Patient (≥ 18 years) under current rituximab therapy who did not develop humoral response against SARS-CoV-2 after their standard vaccination with Biontech/Pfizer or Moderna will be recruited at the Division of Rheumatology in collaboration with the Department of Neurology and the Division of Infectious Diseases and Tropical Medicine, Medical University of Vienna.

Study entry is defined as the date of signature of the study participant (subject) on the informed consent form. All subjects enrolled will be assigned a subject code, consisting of a consecutive subject number (2 digits).

**6.2. Inclusion criteria**

Male and female subjects will be eligible for participation in this study if they:

15. Are ≥18 years on the day of screening
16. Have a chronic condition and have been treated with a B-cell depleting therapy (rituximab) within the last 12 months
17. Received two doses of mRNA SARS-CoV-2 (Biontech/Pfizer or Moderna) vaccine according to recommendations in the label and/or national guidelines.
18. Did not develop humoral immunity 4 weeks after second mRNA vaccination to SARS-CoV-2 (analyzed during the study “Characterization of immune responsiveness after mRNA SARS-CoV-2 Vaccination in patients with immunodeficiency or immunosuppressive therapy”, EK-Nr. 1073/2021, EudraCT Nr. 2021-000291-11)
19. A maximum of 6 months after second vaccination
20. Have an understanding of the study, agree to its provisions, and give written informed consent before study entry
21. If female and capable of bearing children – have a negative urine pregnancy test result at study entry and agree to employ adequate birth control measures for the duration of the study

**6.3.	Exclusion criteria**

Subjects will be excluded from participation in this study if they:

13. Have shown humoral response to the SARS-CoV-2 vaccination
14. Had grade 3 adverse effects from the mRNA vaccination reported
15. Pregnancy and breast feeding
16. Signs of SARS-CoV-2 infection (including previous positive PCR testing)
17. Any other contraindication to any of the study compounds
18. Urgent need for next rituximab application

**6.4.	Study duration**

For the individual study participant, the active blinded study phase will be 4 weeks, with possibility to enroll in an open label extension for additional 8 weeks.

**6.5.	Randomization Procedure**

The web-based computerized randomization algorithm by randomlists.com will be used for randomization. Patients will be randomized in a 1:1 ratio between the third dose mRNA SARS-CoV-2 boost (Biontech/Pfizer or Moderna, respective of their initial vaccination compound) and a single dose boost vector SARS-CoV-2 vaccine (AstraZeneca). Randomization will be stratified by presence or absence of B-cell depletion (determined at screening visit).

**6.6.1.	Blinding**

All patients will be blinded. Blinding of initial treatment allocation will be maintained throughout the 4 week study period. Blinding is ensured during the routine vaccination protocol at the General Hospital Vienna, where patients will see the pre-arranged dose aliquots in syringes without reference to the type of vaccine. To facilitate the process of vaccination of full vials, patients randomized to the same vaccine will be scheduled on the same day. Blinding will be mainly performed to prevent selective drop-outs due to knowledge of treatment allocation.

**6.6.2.	Unblinding Process**

The blinding can be lifted at the request of an investigator, when knowledge of the treatment is essential for appropriate patient management (e.g. after trial termination). In case of an emergency, the principal investigator has the sole responsibility for determining if unblinding of a patient´s treatment assignment is warranted.

If any serious adverse event arises during the study and unblinding appears necessary, the subinvestigator will notify the principal Investigator within two days in maximum. In the following the matter will be discussed and may lead to unblinding.

**6.7.1	Withdrawal and replacement of subjects**

Criteria for withdrawal:

Subjects may prematurely discontinue from the study at any time. The study's premature discontinuation is to be understood when the subject did not undergo complete the last visit (study visit 4) or all pivotal assessments during the study.

Subjects must be withdrawn under the following circumstances:

•	at their own request

•	if the investigator feels it would not be in the best interest of the subject to continue

•	if the subject violates conditions laid out in the consent form/information sheet or disregards instructions by the study personal

In all cases, the reason why subjects are withdrawn must be recorded in detail in the CRF and the subject's medical records. Should the study be discontinued prematurely, all study materials (complete, partially completed, and empty CRFs) will be retained.

Follow-up of patients withdrawn from the study:

In case of premature discontinuation after the start of vaccination, no further investigations concerning the study will be performed. Furthermore, participants may request that no more data will be recorded from the time point of withdrawal and that all biological samples collected in the course of the study will be destroyed.

**6.7.2	Premature termination of the study**

The sponsor has the right to close this study at any time. The IEC and the competent regulatory authority must be informed within 15 days of early termination.

The trial or single-dose steps will be terminated prematurely in the following cases:

- If the number of drop-outs is so high that proper completion of the trial cannot realistically be expected.
- Recruitment is not reaching the critical number

#### 8.8. **Adverse events and reporting**

##### s. Definition of adverse events

An adverse event is any untoward medical occurrence in a patient or clinical investigation subject administered a pharmaceutical product and which does not necessarily have a causal relationship with this treatment. An adverse event (AE) can therefore be any unfavourable and unintended sign (including an abnormal laboratory finding), symptom, or disease temporally associated with the use of a medicinal (investigational) product, whether or not related to the medicinal (investigational) product (see the ICH Guideline for Clinical Safety Data Management: Definitions and Standards for Expedited Reporting)

Adverse events include:

- Exacerbation of a pre-existing disease.
- Increase in frequency or intensity of a pre-existing episodic disease or medical condition.
- Disease or medical condition detected or diagnosed after study drug administration even though it may have been present prior to the start of the study.
- Continuous persistent disease or symptoms present at baseline that worsen following the start of the study.
- Lack of efficacy in the acute treatment of a life-threatening disease.
- Events considered by the Investigator to be related to study-mandated procedures.
- Abnormal assessments, e.g., ECG and physical examination findings, must be reported as AEs if they represent a clinically significant finding that was not present at baseline or worsened during the course of the study.
- Laboratory test abnormalities must be reported as AEs if they represent a clinically significant finding, symptomatic or not, which was not present at baseline or worsened during the course of the study or led to dose reduction, interruption or permanent discontinuation of study drug.

Adverse events do not include:

- Pre-planned interventions or occurrence of endpoints specified in the study protocol are not considered AE´s, if not defined otherwise (eg.as a result of overdose)
- Medical or surgical procedure, e.g., surgery, endoscopy, tooth extraction, transfusion. However, the event leading to the procedure is an AE. If this event is serious, the procedure must be described in the SAE narrative.
- Pre-existing disease or medical condition that does not worsen.
- Situations in which an adverse change did not occur, e.g., hospitalisations for cosmetic elective surgery or for social and/or convenience reasons.

Overdose of either study drug or concomitant medication without any signs or symptoms. However, overdose must be mentioned in the Study Drug Log.

##### t. Definition of serious adverse events (SAEs)

A Serious Adverse Event (SAE) is defined by the International Conference on Harmonization (ICH) guidelines as any AE fulfilling at least one of the following criteria:

- Fatal (including fetal death).
- Life-threatening – defined as an event in which the subject was, in the judgment of the investigator, at risk of death at the time of the event; it does not refer to an event that hypothetically might have caused death had it been more severe.
- Requiring subject's hospitalization or prolongation of existing hospitalization – inpatient hospitalization refers to any inpatient admission, regardless of length of stay.
- Resulting in persistent or significant disability or incapacity (i.e., a substantial disruption of a person’s ability to conduct normal life functions).
- Congenital anomaly or birth defect.
- Is medically significant or requires intervention to prevent at least one of the outcomes listed above.

In case of any SAE, the investigator has to use all means to ensure patient´s safety. Every SAE has to be reported as outlined in section 16.8.

##### u. Definition of suspected or unexpected serious adverse reactions (SUSARs)

SUSARs are all suspected adverse reactions related to the study drug that are both unexpected (not previously described in the SmPC or Investigator’s brochure) and serious.

##### v. Pregnancy

Maternal pregnancies must be reported to the principal investigator/sponsor. To ensure subject safety, each pregnancy must be reported to the PI/sponsor within 2 weeks of learning of its occurrence.

Pregnancies, should be followed up and reported to the PI/sponsor until the outcome of the pregnancy (including premature termination) and status of mother and child is known.

##### w. Severity of adverse events

The severity of clinical AEs is graded on a three-point scale: mild, moderate, severe, and reported on specific AE pages of the CRF.

If the severity of an AE worsens during study drug administration, only the worst intensity should be reported on the AE page. If the AE lessens in intensity, no change in the severity is required:

- *Mild* - Event may be noticeable to subject; does not influence daily activities; the AE resolves spontaneously or may require minimal therapeutic intervention;
- *Moderate* - Event may make subject uncomfortable; performance of daily activities may be influenced; intervention may be needed; the AE produces no sequelae.
- *Severe* - Event may cause noticeable discomfort; usually interferes with daily activities; subject may not be able to continue in the study; the AE produces sequelae, which require prolonged therapeutic intervention.

A mild, moderate or severe AE may or may not be serious.

##### x. Relationship of adverse events to study drug

For all AEs, the investigator will assess the causal relationship between the study drug and the AE using his/her clinical expertise and judgment according to the following algorithm that best fits the circumstances of the AE:

**Not related**

- May or may not follow a temporal sequence from administration of the study product
- Is biologically implausible and does not follow known response pattern to the suspect study drug (if response pattern is previously known).
- Can be explained by the known characteristics of the subject’s clinical state or other modes of therapy administered to the subject.

**Unlikely**

- There is a reasonable temporal relation between the AE and the intake of the study medication, but there is a plausible other explanation for the occurrence of the AE.

**Possibly**

- The AE has a reasonable temporal relationship with drug administration.
- The AE may equally be explained by the study subject’s clinically state, environmental or toxic factors, or concomitant therapy administered to the study subject.
- The relationship between study drug and AE may also be pharmacologically or clinically plausible.

**Probably**

- There is a reasonable temporal relation between the AE and the intake of the study medication, and plausible reasons point to a causal relation with the study medication.

**Related**

- Reasonable temporal relation between the AE and the intake of the study medication and
- There is no other explanation for the AE and
- Subsidence or disappearance of the AE on withdrawal of the study medication and
- Recurrence of the symptoms on restart at previous dose (only applies for re-institution of mediation).

**Not assessable**

The causal relationship between the study drug and the AE cannot be judged.

##### y. Adverse events reporting procedures

A special section is designated to adverse events in the case report form. For each subject, adverse events occurring after signing the informed consent must be recorded on the applicable adverse events page(s) in the case report form. Recording should be done in a concise manner using standard, acceptable medical terms. The adverse event recorded should not be a procedure or a clinical measurement (i.e., a laboratory value or vital sign) but should reflect the reason for the procedure or the diagnosis based on the abnormal measurement. If, in the investigator’s judgment, a clinically significant worsening from baseline is observed in laboratory or other parameters, physical exam finding, or vital sign, a corresponding clinical adverse event should be recorded on the adverse event page(s) of the CRF. If a specific medical diagnosis has been made that diagnosis should be recorded on the adverse event page(s) of the CRF.

SAEs and any newly identified pregnancy (maternal or paternal exposure), malignancy, overdose, opportunistic infection or case of active TB occurring after first administration of study agent in subjects participating in this clinical trial requires submission of a Safety Report Form to the Sponsor (only if other sites are reporting SAE) within 24 hours of notification or observation.

*The following details must thereby be entered:*

- Type of adverse event
- Start (date and time)
- End (date and time)
- Severity (mild, moderate, severe)
- Serious (no / yes)
- Unexpected (no / yes)
- Outcome (resolved, ongoing, ongoing – improved, ongoing – worsening)
- Relation to study drug (unrelated, possibly related, definitely related)

Adverse events are to be documented in the case report form in accordance with the above mentioned criteria.

##### z. Reporting procedures for SAEs

In the event of serious, the investigator has to use all supportive measures for best patient treatment. A serious adverse event must be reported if it occurs during a subject’s participation in the study (whether receiving study agent or not) or within six (6) months of receiving the last dose of study agent.

A Safety Report Form must be completed and faxed to the Sponsor (+43 (0)1 40400 43060) 24 hours of observation or notification of the event. The sponsor will be responsible for potential SUSAR assessment (see below) and reporting SAEs including potential SUSAR to the manufacturing company. A written report is also to be prepared and made available to the clinical investigator within five days.

The following details should at least be available:

- Patient initials and number
- Patient: date of birth, sex, ethical origin
- The suspected investigational medical product (IMP)
- The adverse event assessed as serious
- Short description of the event and outcome

Any serious adverse event that is ongoing when a subject completes his/her participation in the trial must be followed until any of the following occurs:

- The event resolves or stabilizes
- The event returns to baseline condition or value (if a baseline value is available)
- The event can be attributed to agent(s) other than the study agent, or to factors unrelated to study conduct

##### aa. Reporting procedures for SUSARs

All SAEs will be evaluated regarding a possible classification as SUSAR by the sponsor, who will then perform all necessary reports to the manufacturing/distributing pharmaceutical company and forward the SUSAR to the CRO. The CRO will be responsible for reporting SUSARs to the regulatory authorities. In addition, due to their possible safety concern for the study participants, all SUSARs need to be reported to the Institutional Review Board / Independent Ethics Committee (IRB / IEC).

These reports are time critical and should be done within a maximum of 7 days (fatal or life threatening outcome) or 15 days (non fatal, not life threatening). The representative of the Sponsor Investigator shall inform all investigators concerned of relevant information about Serious Unexpected Suspected Adverse Reactions that could adversely affect the safety of subjects.

Such reports shall be made by the study management and the following details should be at least available:

- Patient inclusion number
- Patient: year of birth, sex, ethical origin
- Name of investigator and investigating site
- Period of administration
- The suspected investigational medical product (IMP)
- The adverse event assessed as serious and unexpected, and for which there is a reasonable suspected causal relationship to the IMP
- Concomitant disease and medication
- Short description of the event:
  - Description
  - Onset and if applicable, end
  - Therapeutic intervention
  - Causal relationship
  - Hospitalization of prolongation of hospitalization

9. **METHODOLOGY**

**7.1 Study design and blood samples**

A randomized controlled single-blind phase II trial will be performed. A total of 60 patients with current rituximab therapy who did not develop humoral immune response to SARS-CoV-2 after vaccination with mRNA SARS-CoV-2 vaccine (Biontech/Pfizer or Moderna). Four study visits per patient will be planned during the course of the trial: screening visit (-0-14 days), baseline, week 1, week 4. Patients will be unblinded at week 4. In addition, patients who did not show humoral response at week 4 will be invited for two additional study visits at week 8 and week 12 in an open label approach. Vaccination will take place at baseline visit.

Blood samples for the determination of SARS-CoV-2 antibodies will be obtained at screening visit, at week 1, week 4 and during the potential extension of the study at week 8 and week 12.

**7.2	Study procedures**

**7.2.1	General rules for trial procedures**

•	All study measures like blood sampling and measurements have to be documented with the date (dd:mm:yyyy).

•	In case several study procedures are scheduled simultaneously, there is no specific sequence that should be followed.

•	The dates of all procedures should be according to the protocol. The time margins mentioned in the study flow chart are admissible. If for any reason, a study procedure is not performed within scheduled margins, a protocol deviation should be noted, and the procedure should be performed as soon as possible or as adequate.

•	If it is necessary for organizational reasons, it is admissible to perform procedures scheduled for one visit at two different time points. Allowed time margins should thereby not be exceeded.

**7.2.2	Study visits**

**7.2.2.1 Screening visit (visit 1, -1-28 days, at least 4 weeks after the second immunization with mRNA SARS-CoV-2 vaccine)**

The investigator will inform the subject about the procedures, risks, and benefits of the study. Participants will be informed about future visits, which will be synchronized with the appointment for the third vaccination.

Fully informed, written consent must be obtained from each subject before any assessment is performed. The subject must be allowed sufficient time to consider his/her participation in the study.

The following assessments will be performed:

- Inclusion and exclusion criteria
- Demographic data, including sex, age, weight, and height
- Medical history and concomitant medication
- Assessment of adverse reactions after the mRNA SARS-CoV-2 vaccination
- Assessment of recent COVID-19 disease
- Blood will be drawn:
  - 16 ml of blood will be drawn for PBMC isolation and determination of cellular immunity
  - 8 ml of blood will be drawn for SARS-Cov-2 antibody level detection (prior vaccination).
  - 8 ml of blood will be drawn for determination of leukocyte subpopulations
  - 8 ml of blood will be drawn for biobanking of serum
  - 20 ml of blood will be drawn for routine laboratory testing: blood count, chemistry, coagulation factors
- Pregnancy test in women with childbearing potential
- Randomization to one of the three vaccines: mRNA SARS-CoV-2 (Biontech/Pfizer or Moderna) or vector SARS-CoV-2 vaccination (AstraZeneca)
- SARS-CoV-2 vaccine appointment

**7.2.2.2. Baseline visit (visit 2, week 0, max. 28 days after visit 1)**

The following activities will be performed:

- The investigator will ask all subjects about any adverse experiences occurring since Visit 1 (screening). All adverse experiences will be documented in the CRF.
- Patients will receive a patients diary, fever thermometer and spacer
- Application of mRNA SARS-CoV-2 (Biontech/Pfizer or Moderna) or vector SARS-CoV-2 vaccine (AstraZeneca)
- Patients health status will be observed for additional 30 minutes by a clinician and a nurse in case of any acute reactions to the vaccination

**7.2.2.3. Visit 3 (1 week after the third SARS-CoV-2 vaccination)**

The following activities will be performed:

- The investigator will ask all subjects about any adverse experiences occurring since Visit 2. All adverse experiences will be documented in the CRF.
- Patient´s diaries will be discussed
- Blood will be drawn:
  - 16 ml of blood will be drawn for PBMC isolation and determination of cellular immunity
  - 8 ml of blood will be drawn for SARS-Cov-2 antibody level detection
  - 6 ml of blood will be drawn for determination of leukocyte subpopulations
  - 8 ml of blood will be drawn for biobanking of serum
  - 20 ml of blood will be drawn for routine laboratory testing: blood account, chemistry as well as thrombocytes and coagulation factors
  - 8 ml blood will be drawn for detection of anti-PDE4D antibody

**7.2.2. 4. Visit 4 (4 weeks after the third SARS-CoV-2 vaccination)**

The following activities will be performed:

•	The investigator will ask all subjects about any adverse experiences occurring since Visit 3. All adverse experiences will be documented in the CRF.

- Patient´s diary will be returned to investigator

•	Blood draw:

- 8 ml of blood will be drawn for SARS-Cov-2 antibody level detection
- 6 ml of blood will be drawn for determination of leukocyte subpopulations
- 8 ml of blood will be drawn for biobanking of serum
- 20 ml of blood will be drawn for routine laboratory testing: blood account, chemistry, coagulation factors
- 8 ml blood will be drawn for detection of anti-PDE4D antibody

**7.2.2.5. Visits 5 (week 8) and 6 (week 12) (only for patients enrolling in open label extension)**

Fully informed, written consent to the extension period must be obtained from each subject before any assessment is performed. The subject must be allowed sufficient time to consider his/her participation in the open label extension.

The following activities will be performed:

•	The investigator will ask all subjects about any adverse experiences occurring since Visit 3. All adverse experiences will be documented in the CRF.

- Blood draw:
  - 8 ml of blood will be drawn for SARS-CoV-2 antibody level detection
  - 8 ml of blood will be drawn for biobanking of serum

**7.2.3	Determination of the humoral and cellular immunity**

**7.2.3.1 Determination of serum antibodies against vaccination antigens**

Analysis will be performed at the Department of Laboratory Medicine, Medical University of Vienna. All procedures will be carried out according to standard operating procedures in an ISO 9001:2015 certified environment (ref: 10.1089/bio.2018.0032).

Previous SARS-CoV-2 infection will be detected using nucleocapsid-based chemiluminescence assays (e.g., Roche SARS-CoV-2 NC total antibody ECLIA, Abbott SARS-CoV-2 NC IgG CMIA). Vaccination response will be assessed by spike-protein-based assays (e.g., Technozym RBD ELISA, Siemens RBD immunoassay, Roche SARS-CoV-2 RBD ECLIA).

All analyses will be carried out at the Department of Laboratory Medicine, Medical University of Vienna.

Neutralization assays will be performed as described earlier.^13^

**7.2.3.2 Determination of cellular immune response**

To investigate cellular immunity following SARS-CoV-2 vaccination Peripheral blood mononuclear cells (PBMCs) will be isolated from heparinized venous blood by density gradient centrifugation at 400 *g* of heparinized blood over LSM 1077 Lymphocyte Separation Medium, cryopreserved and stored in liquid nitrogen for later use.

**7.3	Study endpoints**

**7.3.1	Primary study endpoint**

Difference in SARS-CoV-2 antibody seroconversion rate by week 4 after vaccination boost at baseline between 3^rd^ mRNA SARS-CoV-2 (Biontech/Pfizer or Moderna) and vector SARS-CoV-2 vaccine (AstraZeneca).

**7.3.2	Secondary study endpoints**

The secondary endpoints of this study are:

- Overall SARS-Cov-2 antibody seroconversion rate by week 4 after vaccination boost at baseline
- Difference in overall SARS-Cov-2 antibody seroconversion rate by week 4 after vaccination boost at baseline between patient with and without B-cell repopulation.
- Antibody concentrations of SARS-CoV-2 ELISA 4 weeks after vaccination boost at baseline.
- Effect of immunosuppressive comedication on SARS-Cov-2 antibody seroconversion rate by week 4 after vaccination boost at baseline
- Evaluation of cellular immunity before and one week after the vaccination.
- Safety of vaccination boost.

#### 10. **ETHICAL CONSIDERATIONS**

##### a.  Ethical Review

All relevant documents must be reviewed and approved by the Ethics Committee and additionally submitted to the relevant authorities in accordance with the guidance of submission and conduct of clinical trials.

The clinical trial shall be performed in full compliance with the legal regulations according to the Drug Law (AMG - Arzneimittelgesetz) of the Republic of Austria.

An application must also be submitted to the Austrian Competent Authorities (Bundesamt für Sicherheit im Gesundheitswesen (BASG) represented by the Agency for Health and Food Safety (AGES PharmMed), and registered to the European Clinical Trial Database (EudraCT) using the required forms. The timelines for (silent) approval set by national law must be followed before starting the study.

##### b. Consent Procedures

After a detailed information about the study procedures and study medication, as well as the potentially related risks and benefits, the written informed consent will be obtained from each participant by the principal investigator or a designee. At each site, informed consent will be prepared according to the institutional requirements for informed consents. Consent will be collected by the investigator before patient inclusion in trial and before participating in any study procedure.

One copy of each consent form, signed by the participant and by the investigator, will be given to the patient and the original will be kept by the investigator.

##### c. Privacy of patients

All records will be kept confidential. The patient´s name will not be released at any time and data sets for each subject will be identified only by the patient enrollment number.

##### d. Amendments

Proposed amendments must be submitted to the appropriate Competent Authorities (CA) and Ethics Committee (EC) and approved before the change is implemented. These changes are usually presented in the form of an amendment. Amendments that are intended to eliminate an apparent immediate hazard to subjects may be implemented prior to receiving CA/EC approval. However, in this case, approval must be obtained as soon as possible after implementation.

##### e. Insurance

Patients will be covered by a clinical trial insurance. The patients will be insured as defined during their participation in the clinical trial by legal requirements. The investigator of the clinical trial will receive a copy of the insurance conditions of the ´patients insurance´. The sponsor is providing insurance in order to indemnify (legal and financial coverage) the investigator/center against claims arising from the study, except for claims that arise from malpractice and/or negligence. The compensation of the subject in the event of study-related injuries will comply with the applicable regulations. Details on the existing patients insurance are given in the patient information sheet.

Patients will be insured with a national insurance partner. The name and contact details for the insurer will be provided on the informed consent form.

##### f. Regulatory Requirements and GCP

The investigators at all sites are responsible for, and should warrant that the conduct of the study shall be compliant to ICH-GCP, local regulatory requirements with the EC-approved research protocol.

The investigators will ensure, that this study is conducted in full conformance with the principles of the “Declaration of Helsinki” (as amended at the 64th WMA General Assembly, Fortaleza, Brazil, October 2013) and with the national laws and regulations of the country in which the clinical trial is conducted.

g. Research Ethics

Patients who will participate on this clinical trial will receive the third SARS-CoV-2 vaccination in a controlled way and will be informed about their antibody results immediately. Knowledge gained from this study may be helpful to determine efficacy of additional SARS-CoV-2 vaccination. Additional risk equates to the risk of blood draw in general. Therefore pain, local irritation, small bruises, local damage of nerves but also dizziness and infections can occur. Assessed data will be stored from the principal investigator. Only authorized team members have access to the data. Processing of the data is only performed on password protected computers at the General Hospital Vienna.

#### Study Organization

##### h. Data Collection and Case Report From

For each subject enrolled, regardless of the study drug initiation, an CRF must be completed by the investigator or a designated sub-investigator. This also applies to those subjects, who fail to complete the study. If a subject withdraws from the study, the reason must be noted on the CRF. Case report forms are to be completed on an ongoing basis. Entries and corrections will only be performed by study site staff, who have been authorized by the investigator. Entry errors have to be corrected according the ICH-GCP Guidelines.

The entries will be checked by trained personnel (monitor) and any errors or inconsistencies will be checked and corrected immediately.

Data, collected at all visits, are entered into an Excel sheet. The CRFs constitute source documents established before study onset as detailed in the monitoring plan. The Maintenance of the study database will be performed by the Division of Rheumatology of the Medical University of Vienna.

The investigator shall maintain the records of drug disposition and the final CRF´s for a minimum of 15 years after the study closure.

##### i. Monitoring

The principal investigator will ensure, that the trial will be appropriately monitored by ensuring that all the rights of the subject are adequately protected, that the trial data are accurate, complete and verifiable from source documents and that the conduct of the trial is in compliance with the protocol and its subsequent amendments, with GCP and with applicable regulatory requirements. Monitoring will be performed by a contract research organization:

CW-Research & Management GmbH

A-1130 Wien, Auhofstraße 84/3/39

Web: [www.cw-rm.com](https://www.cw-rm.com)

The sponsor investigator will ensure, that monitoring activities occur following a pre-defined monitoring plan.

The investigators will verify, that for all patients a written informed consent is obtained before each subject´s participation in the trial. The investigators will also ensure, that all patients enrolled will be eligible according to the in- and exclusion criteria as defined in the protocol.

The on-site monitors will be responsible for verifying, that the appropriate assurances and certification for training in the protection of human subjects are in place at the site prior to the initiation of the protocol. They will provide education to all site staff regarding the conduct of the study according to good clinical practices (GCP´s). The sponsor will be responsible for ensuring, that informed consent has been obtained for each patient. These records will be verified during the monitoring/auditing visits that will be performed by the designated monitor.

Monitoring visits are planned at the beginning, during and at the end of the study according to the monitoring plan.

##### j. Audit and inspections

All investigators agree to accept audits and inspections by the competent authorities during and after completion of the study. All data and documents may be subject to audits and regulatory inspection.

##### k. Relevant protocol deviations

All protocol deviations will be listed in the study report and assessed as to their influence on the quality of the study analysis. No deviations from the protocol and of any type will be made without complying with all IRB/EC established procedures in accordance with applicable regulations.

##### l. Provision and handling for study medication

Vaccine for the study will be provided by the general Vaccination Board of the County of Vienna. Patients will be vaccinated utilizing the Vienna General Hospital Vaccination infrastructure (“Impfstrasse”, 4Süd). Study vaccine will be stored as recommended by the manufacturer’s instructions.

##### m. Accountability for study medication

The investigator or his/her representative will verify, that study drug supplies are received intact and in the correct amounts. This will be documented with sign and date, as well as the arrival of drugs. All sheets will be kept in the site files as a record of what was received. Additionally, an investigational product accountability log will be documented, including, but not limited to, date received, lot number, kit number, date dispensed, subject number and the identification of the person dispensing the drug.

**9. STATISTICAL ANALYSIS**

**9.1 Sample size considerations**

According to the available number of patients at our Department, including estimates of non-responders to a standard protocol of mRNA vaccination, and expected participation rates, we will attempt to include 60 patients into this trial. Based on a Chi² test comparing 3^rd^ mRNA versus vector vaccine, this number of patients will allow to achieve at least 80% power at a minimal detectable difference of 28% (5% of responders in the mRNA vaccine group versus 33% of responders in the vector vaccine group).

For differences smaller than the minimum detectable difference statistical significance will not be possible to claim.

**9.2 Relevant protocol deviations**

All protocol deviations will be listed in the study report. Major deviations regarding subjects' safety will lead to withdrawal.

**9.3 Endpoints analysis**

The primary endpoint (difference in SARS-CoV-2 antibody seroconversion rate by week 4 after vaccination boost at baseline between third mRNA SARS-CoV-2 (Biontech/Pfizer or Moderna) and vector SARS-CoV-2 vaccine (AstraZeneca)) will be analyzed using Chi-squared Test. For the seroconversion rates, 95% confidence intervals will be calculated. All subjects with available antibody responses fulfilling the eligibility criteria will be included in the analysis.

The secondary endpoints (see section 7.3.2) will be analyzed using descriptive statistics.

**9.4 Missing, unused and spurious data**

Only subjects for whom data are available will be included in the statistical analysis. Missing values will neither be replaced nor estimated.

**9.5 Interim analysis**

No interim analysis will be performed.

**9.6 Software program(s)**

Statistical analysis will be performed using R studios software or SPSS Statistics (Version 17.0 or higher).

#### 12. **DISCLOSURE OF DATA**

The investigator will publish the results of the study as an original scientific report in a peer reviewed scientific journal. In addition, the findings will be presented as oral and poster presentations at international scientific meetings. All obtained data and information will be regarded confidentially.

No data, results or any information of this study may be used for publication without prior agreement of the principal investigator, who is mentioned on the title page of this protocol. The authors of a publication, manuscript, article, abstract or oral presentation, are those, who contribute to the results of the study and/or the writing of the paper. The principal investigator will write the first draft of the paper and – if applicable – delivers the related presentations. He will also be the first author of the main efficacy manuscript. The correspondence address mentioned in each publication is the Sponsor-investigator´s address, as specified on the front-page. Additional co-authors will be determined based on their contributions during the course of the trial.

### SARS-CoV-2 VAC3 study protocol Version 4.0, August 10^th^ 2021

### Clinical Study Protocol

A Randomized, Parallel Group, Single-Blind, Phase 2 Study to Evaluate the immune response of two classes of SARS-CoV-2 Vaccines employed as Third Vaccination in Patients under current Rituximab Therapy and no humoral response after standard mRNA vaccination

**Daniela Sieghart^1^, Selma Tobudic^2^, Martina Durechova^1^, Daffodil Dioso^1^, Michael Zauner^1^, Daniel Mrak^1^, Peter Mandl^1^, Helga Lechner-Radner^1^, Michael Bonelli^1^, Barbara Kornek^3^, Daniel Aletaha^1^**

**1** Department of Internal Medicine III, Division of Rheumatology, Medical University of Vienna, Austria

**2** Department of Medicine I, Division of Infectious Diseases and Tropical Medicine, Medical University of Vienna, Austria

**3 Department of Neurology, Medical University of Vienna, Austria**

**Confidentiality Statement**

The information contained in this document, especially unpublished data, is the property of the sponsor of this study. It is therefore provided to you in confidence as an Investigator, potential Investigator, or consultant, for review by you, your staff, and an Independent Ethics Committee or Institutional Review Board. It is understood that this information will not be disclosed to others without written authorization from the Sponsor except to the extent necessary to obtain informed consent from those persons to whom the study drug may be administered.

### 7. SPONSOR, INVESTIGATOR, MONITOR AND SIGNATURES

**Sponsor/or representative (OEL) (AMG §§ 2a, 31, 32)**

**Univ.-Prof. Dr.med.univ.** **Alexandra Kautzky-Willer,**

Department of Internal medicine III, Medical University of Vienna, Austria

Signature (OEL)	Date

**Coordinating Investigator (AMG §§ 2a, 35, 36**)

**Univ.-Prof. Dr.med.univ.** **Daniel Aletaha,**

Department of Internal Medicine III, Division of Rheumatology, Medical University of Vienna, Austria

Signature	Date

#### 8. **Study summary**

**Background.** Treatment with rituximab (RTX), a monoclonal antibody targeting CD20, constitutes an important therapeutic strategy for patients with several chronic inflammatory conditions such as rheumatoid arthritis, systemic lupus erythematosus, mixed connective tissue diseases and neurological disorders like multiple sclerosis or immune-mediated peripheral neuropathies. Some recent reports have already highlighted the risk of serious consequences upon severe acute respiratory syndrome coronavirus 2 (SARS-CoV-2) infection in patients treated with RTX. Besides the risk of a more severe disease course during B cell depleting therapy, a major concern is the immunogenicity of vaccination during immunomodulatory therapies, especially with RTX. Indeed, RTX has been shown to impair humoral responses to various vaccines including SARS-CoV-2 vaccination. Studies from other vaccines have shown that an additional vaccination or a change of the vaccine can lead to development of a humoral immune response.

**Objective.** To investigate if a third vaccination with a vector based vaccine is superior to a further dose of the vaccine used for basis immunization in previous non responders receiving rituximab.

**Methods.** We will perform a prospective single blind randomized controlled study. A total of 68 patients under rituximab treatment will be enrolled in this clinical trial who received two vaccinations with an mRNA vaccine. Four study visits per patient will be planned. During the screening visit antibodies to the receptor-binding domain will be determined. All patients without detectable humoral immunity against SARS-Cov2 will be invited for a second boost vaccination within 4 weeks after the screening visit. During the baseline visit patients will be randomized to receive a third vaccination with either an mRNA-SARS-CoV-2 vaccine (Biontech/Pfizer or Moderna – according to the vaccine used for their first two vaccinations) or a vector SARS-CoV-2 (AstraZeneca) vaccination as a second boost. Additional study visits are scheduled at weeks 1 and 4 for assessment of humoral and cellular immunity, as well as clinical signs of adverse effects of disease activity reactivation. Patients without humoral response at week 4 will be offered participation in an extended follow-up for additional 8 weeks (i.e. at week 8 and 12 after baseline)

**Expected Results.** We expect at least 30% of patients receiving a third SARS-CoV-2 vaccination to develop humoral response after 4 weeks. The change in mode of action of the vaccine might be beneficial to induce response in patients with rituximab.

##### **3. BACKGROUND**

The current pandemic caused by severe acute respiratory syndrome coronavirus‐2 (SARS‐CoV‐2) has led to exponentially rising morbidity and mortality worldwide. https://coronavirus.jhu.edu/map.html. Apart from aggressive quarantine and hygiene control measures, the most effective way to inhibit SARS‐CoV‐2 spread is a population‐wide vaccination campaign. The SARS-CoV-2 pandemic has required rapid action and the development of vaccines in an unprecedented timeframe. Data from the preclinical development of vaccine candidates for SARS-CoV and MERS-CoV enabled the initial step of experimental vaccine design to be essentially omitted, saving a considerable amount of time. In many cases, production processes were adapted from existing vaccines or vaccine candidates, and in some instances, preclinical and toxicology data from related vaccines could be used. As a result, the first clinical trial of a vaccine candidate for SARS-CoV-2 began in March 2020. Trials were designed such that clinical phases are overlapping and trial starts are staggered, with initial phase I/II trials followed by rapid progression to phase III trials after an interim analysis of the phase I/II data.

More than 180 vaccine candidates, based on several different platforms, are currently in development against SARS-CoV-2.^1^ The World Health Organization (WHO) maintains a working document that includes most of the vaccines in development and is available at https://www.who.int/publications/m/item/draft-landscape-of-covid-19-candidate-vaccines.

RNA vaccines are a relatively recent development. The genetic information for the antigen is delivered instead of the antigen itself. The antigen is then expressed in the vaccinated individual cells. ^2^ Either mRNA (with modifications) or a self-replicating RNA can be used. Higher doses are required for mRNA than for self-replicating RNA, and the RNA is usually delivered via lipid nanoparticles (LNPs).^2-5^

Two mRNA vaccines, BNT162b2, developed by BioNTech/Fosun Pharma/Pfizer, and mRNA-1273 developed by Moderna/NIAID, are currently in phase III trials. ^6^^,^ ^7^

In December 2020, BNT162b2 was under evaluation for emergency use authorization (EUA) for widespread use by several medical regulators globally. On 21 December, the European Medicines Agency (EMA) has given official approval for the BNT162b2 vaccine developed by BioNTech and Pfizer.

On 18 December 2020, mRNA-1273 developed by Moderna/NIAID was issued an emergency use authorization by the United States Food and Drug Administration. On 06 January, the EMA has given official approval for the mRNA-1273, developed by Moderna/NIAID.

Preliminary results from Phase I–II clinical trials on BNT162b2, published in October 2020, indicated the potential for its efficacy and safety. ^8^ ^9^ Findings from studies conducted in the United States and Germany among healthy men and women showed that two 30-μg doses of BNT162b2 elicited high SARS-CoV-2 neutralizing antibody titers and robust antigen-specific CD8+ and Th1-type CD4+ T-cell response. ^10^ Besides, the reactogenicity profile of BNT162b2 represented mainly short-term local (i.e., injection site) and systemic responses. These findings supported the progression of BNT162b2 into the next phase. In phase III, a total of 43548 participants underwent randomization, of whom 43448 received injections: 21720 with BNT162b2 and 21728 with placebo. There were 8 cases of COVID-19 with onset at least 7 days after the second dose among participants assigned to receive BNT162b2 and 162 cases among those assigned to placebo. BNT162b2 was 95% effective in preventing COVID-19 (95% credible interval, 90.3 to 97.6). Similar vaccine efficacy (generally 90 to 100%) was observed across subgroups defined by age, sex, race, ethnicity, baseline body-mass index, and the presence of coexisting conditions.

In July 2020, preliminary results of the Phase I dose-escalation clinical trial of mRNA-1273 were published, showing dose-dependent induction of neutralizing antibodies against SARS-CoV-2 as early as 15 days post-injection. 6 Mild to moderate adverse reactions, such as fever, fatigue, headache, myalgia, and pain at the injection site, were observed in all dose groups but were common with increased dosage. The vaccine in low doses was deemed safe and effective to advance a Phase III clinical trial using two 100-μg doses administered 29 days apart.

In a Phase III trial of mRNA-1273 conducted in the United States between July and October, enrolled and assigned 30 000 volunteers to two groups — one group receiving two 100-μg doses of mRNA-1273 vaccine and the other receiving a placebo of 0.9% sodium chloride. Preliminary data from Phase III clinical trial, published in November 2020, indicating 94% efficacy in preventing COVID-19 infection. Side effects included flu-like symptoms, such as pain at the injection site, fatigue, muscle pain, and headache. The Moderna results were not final – as the trial is not scheduled to conclude until late-2022– and were not peer-reviewed or published in a medical journal.

The efficacy of ChAdOx1 nCoV-19 vaccine was reported from phase III trials in the United Kingdom and Brazil.^11^ The interpretation of the results of these trials is complicated by a dosing error (in which some participants unintentionally received a half-dose for their first of two doses), a small number of participants, and differences in efficacy in the two countries. The overall efficacy in preventing symptomatic infection more than 14 days after the second dose was 70.4% (95%CI: 54.8%-80.6%), with efficacy of 62.1% (95%CI: 41.0%-75.7%) in those who received standard doses, and 90.0% (95%CI 67.4% to 97.0%) in those who received a half-dose followed by a standard dose. Notably, the lower efficacy was based on results from the study in Brazil, and higher efficacy was reported for the study in the United Kingdom. Hospitalizations and severe COVID-19 occurred rarely but exclusively in the placebo arm of these trials. Due to varying intervals between the first and second doses, vaccine efficacy after a single standard dose from day 22 to day 90 was modeled and estimated to be 76% (95%CI: 59%-86%), with maintenance of antibody levels up to day 90. Furthermore, vaccine efficacy appeared to be 82.4% (95%CI: 62.7%-91.7%) when the interval between doses was more than 12 weeks compared to 54.9% (95%CI: 32.7%-69.7%) when the interval was less than 6 weeks. Similarly, geometric mean antibody levels were higher with a longer prime-boost interval in those age 18-55 years.^12^ A larger phase III trial using two standard doses 28 days apart with a majority of the participants in the US recently completed enrollment. Preliminary results of this trial showed vaccine efficacy of 76% (95%CI: 68-82%) at preventing symptomatic infection and 100% efficacy at preventing severe or critical disease and hospitalization. Vaccine efficacy was consistent across ethnicity and age. TheChAdOx1nCoV-19 vaccine has received EUA from the United Kingdom and the European Union. In the United Kingdom, a single-blind multi-center randomized phase II/III trial of the ChAdOx1 nCOV-19 vaccine asked participants to provide a weekly self-administered nose and throat swab starting one week after administration of the first vaccine (or placebo). This study revealed that among those who were infected, vaccinated persons had lower peak viral load and shorter duration of RT-PCR-positive results for SARS-CoV-2 compared to controls, suggesting that the vaccine is effective in reducing transmission.^13^

Patients with immunosuppression with COVID-19 may be at higher risk of hospitalization and ICU admission than matched comparators.^14^ An mRNA vaccine vaccination should be performed as primary immunization with two vaccine-doses 21 or 28 days apart. There are no data about the efficacy and safety of mRNA vaccine against SARS-CoV-2 in patients with a chronic immune-mediated disease such as rheumatoid arthritis, certain kinds of vasculitis, lupus multiple sclerosis. However, after vaccination, immune responses might vary among immunosuppressed patients, dependent upon underlying clinical conditions and the level of immunosuppression. Moreover, it is not clear whether mRNA vaccination against SARS-CoV-2 in the immunosuppressed has a protective effect at all. Specifically, B-cell depleting therapies like CD20 antagonists rituximab have a high risk of vaccination failure.^15^ As for other vaccines, the efficacy of a COVID-19 vaccine may be reduced in patients treated with rituximab. For patients treated with rituximab, it is preferable to administer the vaccine at least 6 months after the last infusion^16^ to have the highest possible number of restored B-cells. The current treatment scheme for rituximab is 500mg or 1000mg intravenously every 6 months.

However, the vaccine is not contraindicated in this population and the treating physician should discuss with the patient the appropriate timing of vaccination taking into consideration the context of pandemic, the risk of poor COVID-19 disease prognosis, and a possibly reduced humoral response to vaccination under rituximab treatment.

**4. STUDY RATIONALE**

Patients who had received rituximab are at high risk of non-response to SARS-CoV-2 vaccination. It is unclear whether patients who did not develop humoral immunity after a standard protocol application with a mRNA vaccine would benefit from a second boost of mRNA SARS-CoV-2 or from a single additional shot of vector SARS-CoV-2 vaccine.

**5. STUDY OBJECTIVES**

The study aims to investigate the humoral and cellular immune responses after a second boost vaccination against SARS-CoV-2 in adult patients treated with rituximab (anti B cell therapy) who did not show response to the first two vaccinations with an mRNA vaccine.

**5.1. Primary Objective (Hypothesis)**

To assess the immunogenicity to a third vaccination mRNA-SARS-CoV-2 vaccine (Biontech/Pfizer or Moderna) compared to a vector SARS-CoV-2 (AstraZeneca) vaccination as a second boost in patients with rituximab by measuring quantitative antibody levels by enzyme-linked immunosorbent assay test (ELISA) and neutralization test (NT) or pseudo viral neutralization assay.

**Null and alternative hypotheses:**

H0: There is no statistical difference in the seroconversion rate between patients receiving a third mRNA vaccination and the patients receiving a second boost with AstraZeneca.

H1: There is statistical difference in the seroconversion rate between patients receiving a third mRNA vaccination and the patients receiving a second boost with AstraZeneca.

**Secondary Objectives**

- To compare cellular immunogenicity of the third mRNA SARS-Cov-2 vaccination will be compared to patients receiving AstraZeneca vaccination as second boost in immunosuppressed patients. T cell proliferation will be assessed, and T-cell cytokine expression will be measured using flow-cytometry following *in vitro* stimulation of peripheral blood mononuclear cells (PBMCs) with SARS-Cov-2 specific antigens.
- To assess safety of a second boost vaccination.
- To evaluate the influence of vaccination on underlying disease activity.

**6. STUDY DESIGN**

A prospective single blind randomized controlled study will be performed. A total of 68 patients with B-cell depleting therapy (rituximab) will be enrolled in this clinical trial. Four study visits per patient will be planned.

After inclusion of the patients (after receiving their written informed consent) serum samples for determination of antibody levels will be obtained at least 4 weeks after the second mRNA vaccination (screening visit). All patients without detectable humoral immunity against SARS-Cov2 will be invited for a second boost vaccination within 4 weeks after the screening visit. Additional study visits are scheduled at baseline (vaccination), and at weeks 1 and 4 for assessment of humoral and cellular immunity, as well as clinical signs of adverse effects of disease activity reactivation. Patients without humoral response at week 4 will be offered participation in an extended follow-up for additional 8 weeks (i.e. at week 8 and 12 after baseline)

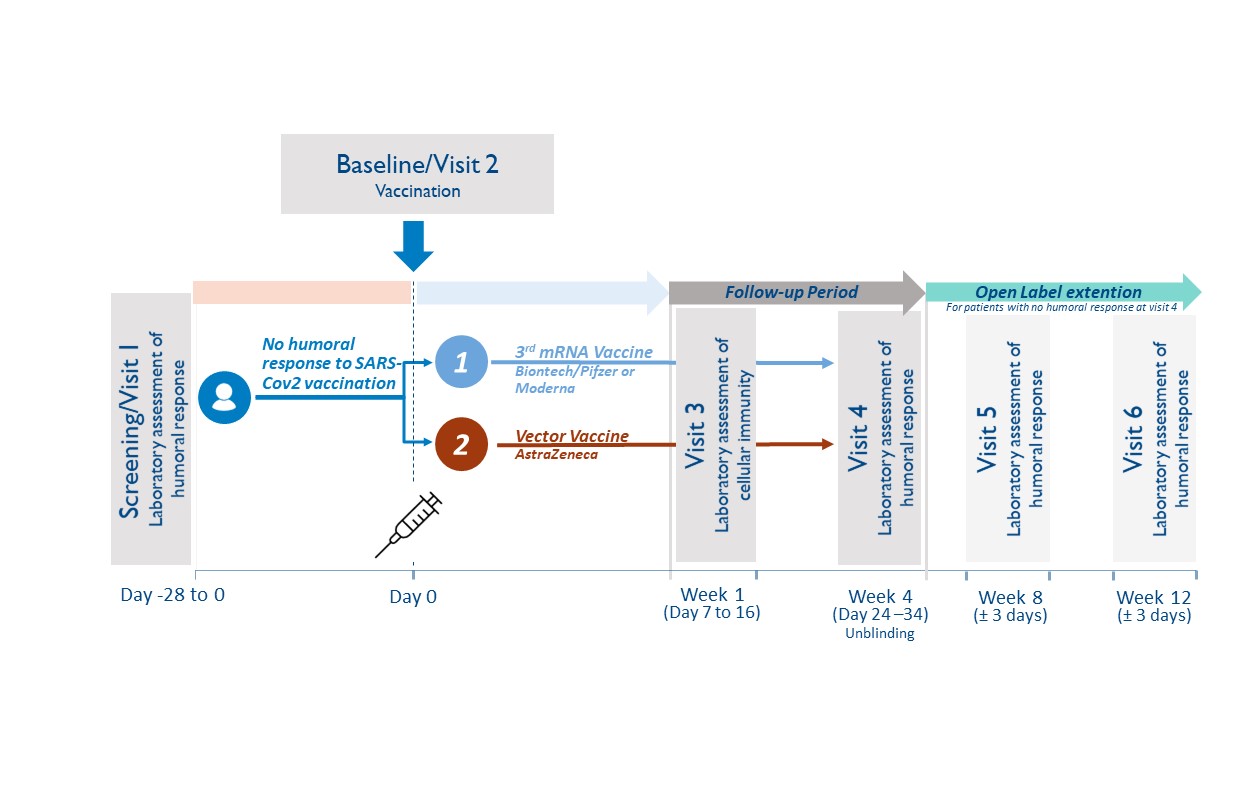

**6.1. Study population**

Adult Patient (≥ 18 years) under current rituximab therapy who did not develop humoral response against SARS-CoV-2 after their standard vaccination with Biontech/Pfizer or Moderna will be recruited at the Division of Rheumatology, Medical University of Vienna.

Study entry is defined as the date of signature of the study participant (subject) on the informed consent form. All subjects enrolled will be assigned a subject code, consisting of a consecutive subject number (2 digits).

**6.2. Inclusion criteria**

Male and female subjects will be eligible for participation in this study if they:

22. Are ≥18 years on the day of screening
23. Have a chronic condition and have been treated with a B-cell depleting therapy (rituximab) within the last 12 months
24. Received two doses of mRNA SARS-CoV-2 (Biontech/Pfizer or Moderna) vaccine according to recommendations in the label and/or national guidelines.
25. Did not develop humoral immunity 4 weeks after second mRNA vaccination to SARS-CoV-2 (analyzed during the study “Characterization of immune responsiveness after mRNA SARS-CoV-2 Vaccination in patients with immunodeficiency or immunosuppressive therapy”, EK-Nr. 1073/2021, EudraCT Nr. 2021-000291-11)
26. A maximum of 6 months after second vaccination
27. Have an understanding of the study, agree to its provisions, and give written informed consent before study entry
28. If female and capable of bearing children – have a negative urine pregnancy test result at study entry and agree to employ adequate birth control measures for the duration of the study

**6.3.	Exclusion criteria**

Subjects will be excluded from participation in this study if they:

19. Have shown humoral response to the SARS-CoV-2 vaccination
20. Had grade 3 adverse effects from the mRNA vaccination reported
21. Pregnancy and breast feeding
22. Signs of SARS-CoV-2 infection (including previous positive PCR testing)
23. Any other contraindication to any of the study compounds
24. Urgent need for next rituximab application

**6.4.	Study duration**

For the individual study participant, the active blinded study phase will be 4 weeks, with possibility to enroll in an open label extension for additional 8 weeks.

**6.5.	Randomization Procedure**

The web-based computerized randomization algorithm “Randomizer for Clinical Trials” by the Medical University of Vienna will be used for randomization. Patients will be randomized in a 1:1 ratio between the third dose mRNA SARS-CoV-2 boost (Biontech/Pfizer or Moderna, respective of their initial vaccination compound) and a single dose boost vector SARS-CoV-2 vaccine (AstraZeneca). Randomization will be stratified by presence or absence of B-cell depletion (determined at screening visit).

**6.6.1.	Blinding**

All patients will be blinded. Blinding of initial treatment allocation will be maintained throughout the 4 week study period. Blinding is ensured during the routine vaccination protocol at the General Hospital Vienna, where patients will see the pre-arranged dose aliquots in syringes without reference to the type of vaccine. To facilitate the process of vaccination of full vials, patients randomized to the same vaccine will be scheduled on the same day. Blinding will be mainly performed to prevent selective drop-outs due to knowledge of treatment allocation.

**6.6.2.	Unblinding Process**

The blinding can be lifted at the request of an investigator, when knowledge of the treatment is essential for appropriate patient management (e.g. after trial termination). In case of an emergency, the principal investigator has the sole responsibility for determining if unblinding of a patient´s treatment assignment is warranted.

If any serious adverse event arises during the study and unblinding appears necessary, the subinvestigator will notify the principal Investigator within two days in maximum. In the following the matter will be discussed and may lead to unblinding.

**6.7.1	Withdrawal and replacement of subjects**

Criteria for withdrawal:

Subjects may prematurely discontinue from the study at any time. The study's premature discontinuation is to be understood when the subject did not undergo complete the last visit (study visit 4) or all pivotal assessments during the study.

Subjects must be withdrawn under the following circumstances:

•	at their own request

•	if the investigator feels it would not be in the best interest of the subject to continue

•	if the subject violates conditions laid out in the consent form/information sheet or disregards instructions by the study personal

In all cases, the reason why subjects are withdrawn must be recorded in detail in the CRF and the subject's medical records. Should the study be discontinued prematurely, all study materials (complete, partially completed, and empty CRFs) will be retained.

Follow-up of patients withdrawn from the study:

In case of premature discontinuation after the start of vaccination, no further investigations concerning the study will be performed. Furthermore, participants may request that no more data will be recorded from the time point of withdrawal and that all biological samples collected in the course of the study will be destroyed.

**6.7.2	Premature termination of the study**

The sponsor has the right to close this study at any time. The IEC and the competent regulatory authority must be informed within 15 days of early termination.

The trial or single-dose steps will be terminated prematurely in the following cases:

- If the number of drop-outs is so high that proper completion of the trial cannot realistically be expected.
- Recruitment is not reaching the critical number

#### 9.8. **Adverse events and reporting**

##### bb. Definition of adverse events

An adverse event is any untoward medical occurrence in a patient or clinical investigation subject administered a pharmaceutical product and which does not necessarily have a causal relationship with this treatment. An adverse event (AE) can therefore be any unfavourable and unintended sign (including an abnormal laboratory finding), symptom, or disease temporally associated with the use of a medicinal (investigational) product, whether or not related to the medicinal (investigational) product (see the ICH Guideline for Clinical Safety Data Management: Definitions and Standards for Expedited Reporting)

Adverse events include:

- Exacerbation of a pre-existing disease.
- Increase in frequency or intensity of a pre-existing episodic disease or medical condition.
- Disease or medical condition detected or diagnosed after study drug administration even though it may have been present prior to the start of the study.
- Continuous persistent disease or symptoms present at baseline that worsen following the start of the study.
- Lack of efficacy in the acute treatment of a life-threatening disease.
- Events considered by the Investigator to be related to study-mandated procedures.
- Abnormal assessments, e.g., ECG and physical examination findings, must be reported as AEs if they represent a clinically significant finding that was not present at baseline or worsened during the course of the study.
- Laboratory test abnormalities must be reported as AEs if they represent a clinically significant finding, symptomatic or not, which was not present at baseline or worsened during the course of the study or led to dose reduction, interruption or permanent discontinuation of study drug.

Adverse events do not include:

- Pre-planned interventions or occurrence of endpoints specified in the study protocol are not considered AE´s, if not defined otherwise (eg.as a result of overdose)
- Medical or surgical procedure, e.g., surgery, endoscopy, tooth extraction, transfusion. However, the event leading to the procedure is an AE. If this event is serious, the procedure must be described in the SAE narrative.
- Pre-existing disease or medical condition that does not worsen.
- Situations in which an adverse change did not occur, e.g., hospitalisations for cosmetic elective surgery or for social and/or convenience reasons.

Overdose of either study drug or concomitant medication without any signs or symptoms. However, overdose must be mentioned in the Study Drug Log.

##### cc. Definition of serious adverse events (SAEs)

A Serious Adverse Event (SAE) is defined by the International Conference on Harmonization (ICH) guidelines as any AE fulfilling at least one of the following criteria:

- Fatal (including fetal death).
- Life-threatening – defined as an event in which the subject was, in the judgment of the investigator, at risk of death at the time of the event; it does not refer to an event that hypothetically might have caused death had it been more severe.
- Requiring subject's hospitalization or prolongation of existing hospitalization – inpatient hospitalization refers to any inpatient admission, regardless of length of stay.
- Resulting in persistent or significant disability or incapacity (i.e., a substantial disruption of a person’s ability to conduct normal life functions).
- Congenital anomaly or birth defect.
- Is medically significant or requires intervention to prevent at least one of the outcomes listed above.

In case of any SAE, the investigator has to use all means to ensure patient´s safety. Every SAE has to be reported as outlined in section 16.8.

##### dd. Definition of suspected or unexpected serious adverse reactions (SUSARs)

SUSARs are all suspected adverse reactions related to the study drug that are both unexpected (not previously described in the SmPC or Investigator’s brochure) and serious.

##### ee. Pregnancy

Maternal pregnancies must be reported to the principal investigator/sponsor. To ensure subject safety, each pregnancy must be reported to the PI/sponsor within 2 weeks of learning of its occurrence.

Pregnancies, should be followed up and reported to the PI/sponsor until the outcome of the pregnancy (including premature termination) and status of mother and child is known.

##### ff. Severity of adverse events

The severity of clinical AEs is graded on a three-point scale: mild, moderate, severe, and reported on specific AE pages of the CRF.

If the severity of an AE worsens during study drug administration, only the worst intensity should be reported on the AE page. If the AE lessens in intensity, no change in the severity is required:

- *Mild* - Event may be noticeable to subject; does not influence daily activities; the AE resolves spontaneously or may require minimal therapeutic intervention;
- *Moderate* - Event may make subject uncomfortable; performance of daily activities may be influenced; intervention may be needed; the AE produces no sequelae.
- *Severe* - Event may cause noticeable discomfort; usually interferes with daily activities; subject may not be able to continue in the study; the AE produces sequelae, which require prolonged therapeutic intervention.

A mild, moderate or severe AE may or may not be serious.

##### gg. Relationship of adverse events to study drug

For all AEs, the investigator will assess the causal relationship between the study drug and the AE using his/her clinical expertise and judgment according to the following algorithm that best fits the circumstances of the AE:

**Not related**

- May or may not follow a temporal sequence from administration of the study product
- Is biologically implausible and does not follow known response pattern to the suspect study drug (if response pattern is previously known).
- Can be explained by the known characteristics of the subject’s clinical state or other modes of therapy administered to the subject.

**Unlikely**

- There is a reasonable temporal relation between the AE and the intake of the study medication, but there is a plausible other explanation for the occurrence of the AE.

**Possibly**

- The AE has a reasonable temporal relationship with drug administration.
- The AE may equally be explained by the study subject’s clinically state, environmental or toxic factors, or concomitant therapy administered to the study subject.
- The relationship between study drug and AE may also be pharmacologically or clinically plausible.

**Probably**

- There is a reasonable temporal relation between the AE and the intake of the study medication, and plausible reasons point to a causal relation with the study medication.

**Related**

- Reasonable temporal relation between the AE and the intake of the study medication and
- There is no other explanation for the AE and
- Subsidence or disappearance of the AE on withdrawal of the study medication and
- Recurrence of the symptoms on restart at previous dose (only applies for re-institution of mediation).

**Not assessable**

The causal relationship between the study drug and the AE cannot be judged.

##### hh. Adverse events reporting procedures

A special section is designated to adverse events in the case report form. For each subject, adverse events occurring after signing the informed consent must be recorded on the applicable adverse events page(s) in the case report form. Recording should be done in a concise manner using standard, acceptable medical terms. The adverse event recorded should not be a procedure or a clinical measurement (i.e., a laboratory value or vital sign) but should reflect the reason for the procedure or the diagnosis based on the abnormal measurement. If, in the investigator’s judgment, a clinically significant worsening from baseline is observed in laboratory or other parameters, physical exam finding, or vital sign, a corresponding clinical adverse event should be recorded on the adverse event page(s) of the CRF. If a specific medical diagnosis has been made that diagnosis should be recorded on the adverse event page(s) of the CRF.

SAEs and any newly identified pregnancy (maternal or paternal exposure), malignancy, overdose, opportunistic infection or case of active TB occurring after first administration of study agent in subjects participating in this clinical trial requires submission of a Safety Report Form to the Sponsor (only if other sites are reporting SAE) within 24 hours of notification or observation.

*The following details must thereby be entered:*

- Type of adverse event
- Start (date and time)
- End (date and time)
- Severity (mild, moderate, severe)
- Serious (no / yes)
- Unexpected (no / yes)
- Outcome (resolved, ongoing, ongoing – improved, ongoing – worsening)
- Relation to study drug (unrelated, possibly related, definitely related)

Adverse events are to be documented in the case report form in accordance with the above mentioned criteria.

##### ii. Reporting procedures for SAEs

In the event of serious, the investigator has to use all supportive measures for best patient treatment. A serious adverse event must be reported if it occurs during a subject’s participation in the study (whether receiving study agent or not) or within six (6) months of receiving the last dose of study agent.

A Safety Report Form must be completed and faxed to the Sponsor (+43 (0)1 40400 43060) 24 hours of observation or notification of the event. The sponsor will be responsible for potential SUSAR assessment (see below) and reporting SAEs including potential SUSAR to the manufacturing company. A written report is also to be prepared and made available to the clinical investigator within five days.

The following details should at least be available:

- Patient initials and number
- Patient: date of birth, sex, ethical origin
- The suspected investigational medical product (IMP)
- The adverse event assessed as serious
- Short description of the event and outcome

Any serious adverse event that is ongoing when a subject completes his/her participation in the trial must be followed until any of the following occurs:

- The event resolves or stabilizes
- The event returns to baseline condition or value (if a baseline value is available)
- The event can be attributed to agent(s) other than the study agent, or to factors unrelated to study conduct

##### jj. Reporting procedures for SUSARs

All SAEs will be evaluated regarding a possible classification as SUSAR by the sponsor, who will then perform all necessary reports to the manufacturing/distributing pharmaceutical company and forward the SUSAR to the CRO. The CRO will be responsible for reporting SUSARs to the regulatory authorities. In addition, due to their possible safety concern for the study participants, all SUSARs need to be reported to the Institutional Review Board / Independent Ethics Committee (IRB / IEC).

These reports are time critical and should be done within a maximum of 7 days (fatal or life threatening outcome) or 15 days (non fatal, not life threatening). The representative of the Sponsor Investigator shall inform all investigators concerned of relevant information about Serious Unexpected Suspected Adverse Reactions that could adversely affect the safety of subjects.

Such reports shall be made by the study management and the following details should be at least available:

- Patient inclusion number
- Patient: year of birth, sex, ethical origin
- Name of investigator and investigating site
- Period of administration
- The suspected investigational medical product (IMP)
- The adverse event assessed as serious and unexpected, and for which there is a reasonable suspected causal relationship to the IMP
- Concomitant disease and medication
- Short description of the event:
  - Description
  - Onset and if applicable, end
  - Therapeutic intervention
  - Causal relationship
  - Hospitalization of prolongation of hospitalization

10. **METHODOLOGY**

**7.1 Study design and blood samples**

A randomized controlled single-blind phase II trial will be performed. A total of 68 patients with current rituximab therapy who did not develop humoral immune response to SARS-CoV-2 after vaccination with mRNA SARS-CoV-2 vaccine (Biontech/Pfizer or Moderna). Four study visits per patient will be planned during the course of the trial: screening visit (-0-14 days), baseline, week 1, week 4. Patients will be unblinded at week 4. In addition, patients who did not show humoral response at week 4 will be invited for two additional study visits at week 8 and week 12 in an open label approach. Vaccination will take place at baseline visit.

Blood samples for the determination of SARS-CoV-2 antibodies will be obtained at screening visit, at week 1, week 4 and during the potential extension of the study at week 8 and week 12.

**7.2	Study procedures**

**7.2.1	General rules for trial procedures**

•	All study measures like blood sampling and measurements have to be documented with the date (dd:mm:yyyy).

•	In case several study procedures are scheduled simultaneously, there is no specific sequence that should be followed.

•	The dates of all procedures should be according to the protocol. The time margins mentioned in the study flow chart are admissible. If for any reason, a study procedure is not performed within scheduled margins, a protocol deviation should be noted, and the procedure should be performed as soon as possible or as adequate.

•	If it is necessary for organizational reasons, it is admissible to perform procedures scheduled for one visit at two different time points. Allowed time margins should thereby not be exceeded.

**7.2.2	Study visits**

**7.2.2.1 Screening visit (visit 1, -1-28 days, at least 4 weeks after the second immunization with mRNA SARS-CoV-2 vaccine)**

The investigator will inform the subject about the procedures, risks, and benefits of the study. Participants will be informed about future visits, which will be synchronized with the appointment for the third vaccination.

Fully informed, written consent must be obtained from each subject before any assessment is performed. The subject must be allowed sufficient time to consider his/her participation in the study.

The following assessments will be performed:

- Inclusion and exclusion criteria
- Demographic data, including sex, age, weight, and height
- Medical history and concomitant medication
- Assessment of adverse reactions after the mRNA SARS-CoV-2 vaccination
- Assessment of recent COVID-19 disease
- Blood will be drawn:
  - 16 ml of blood will be drawn for PBMC isolation and determination of cellular immunity
  - 8 ml of blood will be drawn for SARS-Cov-2 antibody level detection (prior vaccination).
  - 8 ml of blood will be drawn for determination of leukocyte subpopulations
  - 8 ml of blood will be drawn for biobanking of serum
  - 20 ml of blood will be drawn for routine laboratory testing: blood count, chemistry, coagulation factors
- Pregnancy test in women with childbearing potential
- Randomization to one of the three vaccines: mRNA SARS-CoV-2 (Biontech/Pfizer or Moderna) or vector SARS-CoV-2 vaccination (AstraZeneca)
- SARS-CoV-2 vaccine appointment

**7.2.2.2. Baseline visit (visit 2, week 0, max. 28 days after visit 1)**

The following activities will be performed:

- The investigator will ask all subjects about any adverse experiences occurring since Visit 1 (screening). All adverse experiences will be documented in the CRF.
- Patients will receive a patients diary, fever thermometer and spacer
- Application of mRNA SARS-CoV-2 (Biontech/Pfizer or Moderna) or vector SARS-CoV-2 vaccine (AstraZeneca)
- Patients health status will be observed for additional 30 minutes by a clinician and a nurse in case of any acute reactions to the vaccination

**7.2.2.3. Visit 3 (1 week after the third SARS-CoV-2 vaccination)**

The following activities will be performed:

- The investigator will ask all subjects about any adverse experiences occurring since Visit 2. All adverse experiences will be documented in the CRF.
- Patient´s diaries will be discussed
- Blood will be drawn:
  - 16 ml of blood will be drawn for PBMC isolation and determination of cellular immunity
  - 8 ml of blood will be drawn for SARS-Cov-2 antibody level detection
  - 6 ml of blood will be drawn for determination of leukocyte subpopulations
  - 8 ml of blood will be drawn for biobanking of serum
  - 20 ml of blood will be drawn for routine laboratory testing: blood account, chemistry as well as thrombocytes and coagulation factors
  - 8 ml blood will be drawn for detection of anti-PDE4D antibody

**7.2.2. 4. Visit 4 (4 weeks after the third SARS-CoV-2 vaccination)**

The following activities will be performed:

•	The investigator will ask all subjects about any adverse experiences occurring since Visit 3. All adverse experiences will be documented in the CRF.

- Patient´s diary will be returned to investigator

•	Blood draw:

- 8 ml of blood will be drawn for SARS-Cov-2 antibody level detection
- 6 ml of blood will be drawn for determination of leukocyte subpopulations
- 8 ml of blood will be drawn for biobanking of serum
- 20 ml of blood will be drawn for routine laboratory testing: blood account, chemistry, coagulation factors
- 8 ml blood will be drawn for detection of anti-PDE4D antibody

**7.2.2.5. Visits 5 (week 8) and 6 (week 12) (only for patients enrolling in open label extension)**

Fully informed, written consent to the extension period must be obtained from each subject before any assessment is performed. The subject must be allowed sufficient time to consider his/her participation in the open label extension.

The following activities will be performed:

•	The investigator will ask all subjects about any adverse experiences occurring since Visit 3. All adverse experiences will be documented in the CRF.

- Blood draw:
  - 8 ml of blood will be drawn for SARS-CoV-2 antibody level detection
  - 8 ml of blood will be drawn for biobanking of serum

**7.2.3	Determination of the humoral and cellular immunity**

**7.2.3.1 Determination of serum antibodies against vaccination antigens**

Analysis will be performed at the Department of Laboratory Medicine, Medical University of Vienna. All procedures will be carried out according to standard operating procedures in an ISO 9001:2015 certified environment (ref: 10.1089/bio.2018.0032).

Previous SARS-CoV-2 infection will be detected using nucleocapsid-based chemiluminescence assays (e.g., Roche SARS-CoV-2 NC total antibody ECLIA, Abbott SARS-CoV-2 NC IgG CMIA). Vaccination response will be assessed by spike-protein-based assays (e.g., Technozym RBD ELISA, Siemens RBD immunoassay, Roche SARS-CoV-2 RBD ECLIA).

All analyses will be carried out at the Department of Laboratory Medicine, Medical University of Vienna.

Neutralization assays will be performed as described earlier.^13^

**7.2.3.2 Determination of cellular immune response**

To investigate cellular immunity following SARS-CoV-2 vaccination Peripheral blood mononuclear cells (PBMCs) will be isolated from heparinized venous blood by density gradient centrifugation at 400 *g* of heparinized blood over LSM 1077 Lymphocyte Separation Medium, cryopreserved and stored in liquid nitrogen for later use.

**7.3	Study endpoints**

**7.3.1	Primary study endpoint**

Difference in SARS-CoV-2 antibody seroconversion rate by week 4 after vaccination boost at baseline between 3^rd^ mRNA SARS-CoV-2 (Biontech/Pfizer or Moderna) and vector SARS-CoV-2 vaccine (AstraZeneca).

**7.3.2	Secondary study endpoints**

The secondary endpoints of this study are:

- Overall SARS-Cov-2 antibody seroconversion rate by week 4 after vaccination boost at baseline
- Difference in overall SARS-Cov-2 antibody seroconversion rate by week 4 after vaccination boost at baseline between patient with and without B-cell repopulation.
- Antibody concentrations of SARS-CoV-2 ELISA 4 weeks after vaccination boost at baseline.
- Effect of immunosuppressive comedication on SARS-Cov-2 antibody seroconversion rate by week 4 after vaccination boost at baseline
- Evaluation of cellular immunity before and one week after the vaccination.
- Safety of vaccination boost.

#### 11. **ETHICAL CONSIDERATIONS**

##### a.  Ethical Review

All relevant documents must be reviewed and approved by the Ethics Committee and additionally submitted to the relevant authorities in accordance with the guidance of submission and conduct of clinical trials.

The clinical trial shall be performed in full compliance with the legal regulations according to the Drug Law (AMG - Arzneimittelgesetz) of the Republic of Austria.

An application must also be submitted to the Austrian Competent Authorities (Bundesamt für Sicherheit im Gesundheitswesen (BASG) represented by the Agency for Health and Food Safety (AGES PharmMed), and registered to the European Clinical Trial Database (EudraCT) using the required forms. The timelines for (silent) approval set by national law must be followed before starting the study.

##### b. Consent Procedures

After a detailed information about the study procedures and study medication, as well as the potentially related risks and benefits, the written informed consent will be obtained from each participant by the principal investigator or a designee. At each site, informed consent will be prepared according to the institutional requirements for informed consents. Consent will be collected by the investigator before patient inclusion in trial and before participating in any study procedure.

One copy of each consent form, signed by the participant and by the investigator, will be given to the patient and the original will be kept by the investigator.

##### c. Privacy of patients

All records will be kept confidential. The patient´s name will not be released at any time and data sets for each subject will be identified only by the patient enrollment number.

##### d. Amendments

Proposed amendments must be submitted to the appropriate Competent Authorities (CA) and Ethics Committee (EC) and approved before the change is implemented. These changes are usually presented in the form of an amendment. Amendments that are intended to eliminate an apparent immediate hazard to subjects may be implemented prior to receiving CA/EC approval. However, in this case, approval must be obtained as soon as possible after implementation.

##### e. Insurance

Patients will be covered by a clinical trial insurance. The patients will be insured as defined during their participation in the clinical trial by legal requirements. The investigator of the clinical trial will receive a copy of the insurance conditions of the ´patients insurance´. The sponsor is providing insurance in order to indemnify (legal and financial coverage) the investigator/center against claims arising from the study, except for claims that arise from malpractice and/or negligence. The compensation of the subject in the event of study-related injuries will comply with the applicable regulations. Details on the existing patients insurance are given in the patient information sheet.

Patients will be insured with a national insurance partner. The name and contact details for the insurer will be provided on the informed consent form.

##### f. Regulatory Requirements and GCP

The investigators at all sites are responsible for, and should warrant that the conduct of the study shall be compliant to ICH-GCP, local regulatory requirements with the EC-approved research protocol.

The investigators will ensure, that this study is conducted in full conformance with the principles of the “Declaration of Helsinki” (as amended at the 64th WMA General Assembly, Fortaleza, Brazil, October 2013) and with the national laws and regulations of the country in which the clinical trial is conducted.

g. Research Ethics

Patients who will participate on this clinical trial will receive the third SARS-CoV-2 vaccination in a controlled way and will be informed about their antibody results immediately. Knowledge gained from this study may be helpful to determine efficacy of additional SARS-CoV-2 vaccination. Additional risk equates to the risk of blood draw in general. Therefore pain, local irritation, small bruises, local damage of nerves but also dizziness and infections can occur. Assessed data will be stored from the principal investigator. Only authorized team members have access to the data. Processing of the data is only performed on password protected computers at the General Hospital Vienna.

#### Study Organization

##### h. Data Collection and Case Report From

For each subject enrolled, regardless of the study drug initiation, an CRF must be completed by the investigator or a designated sub-investigator. This also applies to those subjects, who fail to complete the study. If a subject withdraws from the study, the reason must be noted on the CRF. Case report forms are to be completed on an ongoing basis. Entries and corrections will only be performed by study site staff, who have been authorized by the investigator. Entry errors have to be corrected according the ICH-GCP Guidelines.

The entries will be checked by trained personnel (monitor) and any errors or inconsistencies will be checked and corrected immediately.

Data, collected at all visits, are entered into an Excel sheet. The CRFs constitute source documents established before study onset as detailed in the monitoring plan. The Maintenance of the study database will be performed by the Division of Rheumatology of the Medical University of Vienna.

The investigator shall maintain the records of drug disposition and the final CRF´s for a minimum of 15 years after the study closure.

##### i. Monitoring

The principal investigator will ensure, that the trial will be appropriately monitored by ensuring that all the rights of the subject are adequately protected, that the trial data are accurate, complete and verifiable from source documents and that the conduct of the trial is in compliance with the protocol and its subsequent amendments, with GCP and with applicable regulatory requirements. Monitoring will be performed by a contract research organization:

CW-Research & Management GmbH

A-1130 Wien, Auhofstraße 84/3/39

Web: [www.cw-rm.com](https://www.cw-rm.com)

The sponsor investigator will ensure, that monitoring activities occur following a pre-defined monitoring plan.

The investigators will verify, that for all patients a written informed consent is obtained before each subject´s participation in the trial. The investigators will also ensure, that all patients enrolled will be eligible according to the in- and exclusion criteria as defined in the protocol.

The on-site monitors will be responsible for verifying, that the appropriate assurances and certification for training in the protection of human subjects are in place at the site prior to the initiation of the protocol. They will provide education to all site staff regarding the conduct of the study according to good clinical practices (GCP´s). The sponsor will be responsible for ensuring, that informed consent has been obtained for each patient. These records will be verified during the monitoring/auditing visits that will be performed by the designated monitor.

Monitoring visits are planned at the beginning, during and at the end of the study according to the monitoring plan.

##### j. Audit and inspections

All investigators agree to accept audits and inspections by the competent authorities during and after completion of the study. All data and documents may be subject to audits and regulatory inspection.

##### k. Relevant protocol deviations

All protocol deviations will be listed in the study report and assessed as to their influence on the quality of the study analysis. No deviations from the protocol and of any type will be made without complying with all IRB/EC established procedures in accordance with applicable regulations.

##### l. Provision and handling for study medication

Vaccine for the study will be provided by the general Vaccination Board of the County of Vienna. Patients will be vaccinated utilizing the Vienna General Hospital Vaccination infrastructure (“Impfstrasse”, 4Süd). Study vaccine will be stored as recommended by the manufacturer’s instructions.

##### m. Accountability for study medication

The investigator or his/her representative will verify, that study drug supplies are received intact and in the correct amounts. This will be documented with sign and date, as well as the arrival of drugs. All sheets will be kept in the site files as a record of what was received. Additionally, an investigational product accountability log will be documented, including, but not limited to, date received, lot number, kit number, date dispensed, subject number and the identification of the person dispensing the drug.

**9. STATISTICAL ANALYSIS**

**9.1 Sample size considerations**

According to the available number of patients at our Department, including estimates of non-responders to a standard protocol of mRNA vaccination, and expected participation rates, we will attempt to include 68 patients into this trial. Based on a Chi² test comparing 3^rd^ mRNA versus vector vaccine, this number of patients will allow to achieve at least 80% power at a minimal detectable difference of 28% (5% of responders in the mRNA vaccine group versus 33% of responders in the vector vaccine group).

For differences smaller than the minimum detectable difference statistical significance will not be possible to claim.

**9.2 Relevant protocol deviations**

All protocol deviations will be listed in the study report. Major deviations regarding subjects' safety will lead to withdrawal.

**9.3 Endpoints analysis**

The primary endpoint (difference in SARS-CoV-2 antibody seroconversion rate by week 4 after vaccination boost at baseline between third mRNA SARS-CoV-2 (Biontech/Pfizer or Moderna) and vector SARS-CoV-2 vaccine (AstraZeneca)) will be analyzed using Chi-squared Test. For the seroconversion rates, 95% confidence intervals will be calculated. All subjects with available antibody responses fulfilling the eligibility criteria will be included in the analysis.

The secondary endpoints (see section 7.3.2) will be analyzed using descriptive statistics.

**9.4 Missing, unused and spurious data**

Only subjects for whom data are available will be included in the statistical analysis. Missing values will neither be replaced nor estimated.

**9.5 Interim analysis**

No interim analysis will be performed.

**9.6 Software program(s)**

Statistical analysis will be performed using R studios software or SPSS Statistics (Version 17.0 or higher).

#### 13. **DISCLOSURE OF DATA**

The investigator will publish the results of the study as an original scientific report in a peer reviewed scientific journal. In addition, the findings will be presented as oral and poster presentations at international scientific meetings. All obtained data and information will be regarded confidentially.

No data, results or any information of this study may be used for publication without prior agreement of the principal investigator, who is mentioned on the title page of this protocol. The authors of a publication, manuscript, article, abstract or oral presentation, are those, who contribute to the results of the study and/or the writing of the paper. The principal investigator will write the first draft of the paper and – if applicable – delivers the related presentations. He will also be the first author of the main efficacy manuscript. The correspondence address mentioned in each publication is the Sponsor-investigator´s address, as specified on the front-page. Additional co-authors will be determined based on their contributions during the course of the trial.
